# Likelihood based Mendelian randomization analysis with automated instrument selection and horizontal pleiotropic modeling

**DOI:** 10.1101/2021.11.03.21265848

**Authors:** Zhongshang Yuan, Lu Liu, Ping Guo, Ran Yan, Fuzhong Xue, Xiang Zhou

## Abstract

Mendelian randomization (MR) is a common tool for identifying causal risk factors underlying diseases. Here, we present a method, MRAID, for effective MR analysis. MRAID borrows ideas from fine mapping analysis to model an initial set of candidate SNPs that are in potentially high linkage disequilibrium with each other and automatically selects among them the suitable instruments for causal inference. MRAID also explicitly models both uncorrelated and correlated horizontal pleiotropic effects that are widespread for complex trait analysis. MRAID achieves both tasks through a joint likelihood framework and relies on a scalable sampling-based algorithm to compute calibrated *p*-values. Comprehensive and realistic simulations show MRAID can provide calibrated type I error control, reduce false positives, while being more powerful than existing approaches. We illustrate the benefits of MRAID for an MR screening analysis across 645 trait pairs in UK Biobank, identifying multiple lifestyle causal risk factors of cardiovascular disease-related traits.

## Introduction

Investigating causal relationship among complex traits and identifying causal risk factors are an important first step towards understanding the biology of diseases. A common statistical tool for performing such causal inference in observational studies is Mendelian randomization (MR). MR is a form of instrumental variable analysis that uses SNPs to serve as instruments for inferring the causal effect of an exposure variable on an outcome variable(*1*). MR requires only summary statistics from genome-wide association studies (GWASs) and is often performed in a two-sample study setting where the exposure variable and the outcome variable are measured in two separate studies(*2*). With the abundant availability of GWAS summary statistics, numerous MR analyses are being carried out, identifying important causal risk factors for various common diseases. These MR studies are facilitated by many recently developed MR methods that include the inverse variance weighted (IVW) method, MR-Egger(*3*), median-based regression(*4*), BWMR(*5*), RAPS(*6*), MRMix(*7*), CAUSE(*8*), to name a few. Different MR methods differ in their modeling assumptions and inference algorithms, but the majority of them encounter two important modeling and algorithmic challenges that have so far limited the effectiveness of MR analysis.

First, almost all existing MR methods rely on a pre-selected set of independent SNPs to serve as instruments for MR analysis. The instruments are selected to be independent from each other to ensure the validity of the statistical inference framework used in many common MR methods such as IVW. The independent SNPs are often selected through linkage disequilibrium (LD) clumping, a procedure that first ranks SNPs based on their marginal association evidence with the exposure variable and then retains SNPs that are not in high LD with the SNPs on top of the ranking list. Using LD clumping to select SNPs may be suboptimal, however, as the selected SNPs may only represent tagging SNPs that are in LD with the causal SNPs rather than the causal ones themselves. Using tagging SNPs instead of the causal ones to serve as instruments can reduce the power of MR analysis. In addition, perhaps more importantly, selecting independent SNPs for MR analysis may not be ideal either, as complex traits can be influenced by multiple causal SNPs residing in the same local region that are in potential LD with each other. Consequently, selecting independent SNPs may only capture a small proportion of the phenotypic variance in the exposure variable, again leading to a loss of power in the subsequent MR analysis (*1, 2, 9, 10*). Indeed, in the parallel research field of transcriptome-wide association studies, it has been well documented that incorporating correlated SNPs can substantially improve analysis power than using independent SNPs only(*11-14*). Therefore, incorporating correlated SNPs and developing effective approaches to select instruments among them are important to fully captivate the potential of MR.

Second, only a limited number of MR methods model horizontal pleiotropy and even fewer can effectively control for it during MR analysis(*15*). Horizontal pleiotropy occurs when the SNP instruments exhibit effects on the outcome through pathways other than the exposure. Horizontal pleiotropy has been widely observed in complex trait analysis(*13, 15*) and often comes in two distinct types. The first type of horizontal pleiotropy arises through paths independent of the exposure, with the resulting horizontal pleotropic effects being independent of the SNP effects on the exposure. The second type of horizontal pleiotropy arises through unobserved exposure-outcome confounders and induces correlation between the horizontal pleotropic effects and the SNP effects on the exposure. The presence of either type of horizontal pleiotropy violates standard MR modeling assumptions and can lead to biased causal effect estimates and increased false discoveries. Early MR analyses control for horizontal pleiotropy by simply removing instrumental SNPs that are potentially associated with the outcome variable(*15-18*). Removing SNPs associated with the outcome would result in a conservative set of selected instruments and lead to a loss of power in the subsequent MR analysis. Recent MR methods explicitly model horizontal pleiotropy by specifying modeling assumptions on the horizontal pleiotropic effects. For example, the Egger assumption assumes the same horizontal pleiotropic effect across SNP instruments(*3, 13*), while PMR-VC(*13*) and BWMR(*5*) assume the horizontal pleiotropic effects to follow a normal distribution; all these methods model the first type of horizontal pleiotropy. MRMix(*7*) and CAUSE(*8*), by contrast, employ a normal-mixture model to control for both types of horizontal pleiotropy. Unfortunately, modeling both types of horizontal pleiotropy has been technically challenging, as the resulting likelihood function of the MR model often consists of an integration that cannot be solved analytically. Consequently, both MRMix and CAUSE rely on non-likelihood based approaches to perform MR inference. Specifically, MRMix searches on a grid of causal effect candidates to identify the one that maximizes the proportion of GWAS summary statistics residing in the expected sub-model without horizontal pleiotropy. CAUSE contrasts the out-of-sample prediction accuracy between two different models, one with the causal effect and the other without, by computing the expected log pointwise posterior density between the two, for causal inference. Non-likelihood based causal inference, however, can lead to a loss of power and/or uncalibrated test statistics that are essential for large-scale screening of causal risk factors underlying diseases. Indeed, as we will show here, MRMix is not robust to modeling misspecifications on the instrumental effect sizes and is prone to estimation bias, while CAUSE yields overly conservative *p*-values.

Here, we present a likelihood-based two-sample MR method for causal inference that overcomes the above two challenges. Specifically, our method models an initial set of candidate SNP instruments that are in high LD with each other and automatically selects among them the suitable instruments for MR analysis. In addition, our method accounts for both types of horizontal pleiotropy in a likelihood framework and relies on a scalable sampling-based algorithm for calibrated *p*-values computation. We refer to our method as the two-sample Mendelian Randomization with Automated Instrument Determination (MRAID). We demonstrate the effectiveness of MRAID through comprehensive and realistic simulations. We also apply MRAID for an MR screening analysis across 645 trait pairs in the UK Biobank(*19*), identifying lifestyle risk factors that may causally influence cardiovascular disease-related traits.

## Results

### Method overview

MRAID is described in the Materials and Methods, with its technical details provided in the Supplementary Text and a method schematic shown in Fig. 1. Briefly, MRAID is a two-sample MR method that aims to infer the causal effect of an exposure variable on an outcome variable using GWAS summary statistics. MRAID models jointly all genome-wide significant SNPs that are in potential LD with each other and performs automated instrument selection among them to identify suitable instruments for MR analysis. In addition, MRAID explicitly accounts for two types of horizontal pleiotropic effects through a maximum likelihood-based inference framework and is scalable to biobank datasets (Table 1).

**Fig. 1.**
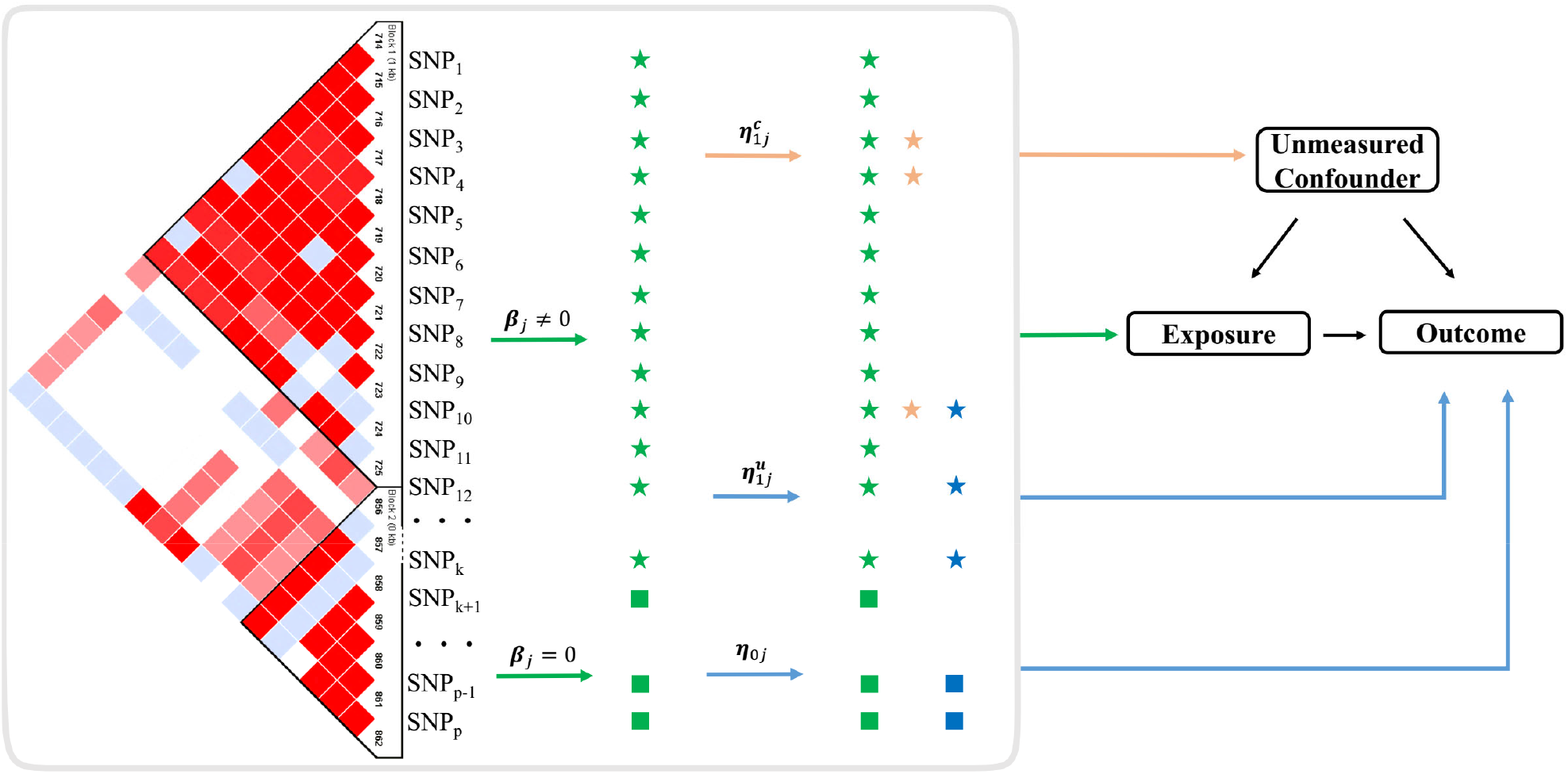
Schematic of MRAID. MRAID is a Mendelian randomization method that infers the causal effect of an exposure on an outcome in the presence of unmeasured confounder by using SNPs as instrumental variables. MRAID first obtains an initial set of candidate SNP instruments that are marginally associated with the exposure (SNP_1_, …, SNP_p_) and that are in potential LD with each other (LD plot on left). MRAID imposes a sparsity assumption on the instrumental effects of the candidate SNPs to select instruments with non-zero effects on the exposure (green stars, selected by a green arrow). Among the selected instruments, MRAID assumes that a proportion of them display horizontal pleiotropic effects that are uncorrelated with instrumental effects (blue stars, selected by a blue arrow) and that another proportion of them display horizontal pleiotropic effects that are correlated with instrumental effects (orange stars, selected by an orange arrow). Among the non-selected instrument candidates (green squares, selected by a green arrow), MRAID also assumes that a proportion of them display horizontal pleiotropic effects that are uncorrelated with instrumental effects (blue squares, selected by a blue arrow). Overall, MRAID models jointly all genome-wide significant SNPs that are in potential LD with each other and performs automated instrument selection among them to identify suitable instruments. MRAID explicitly accounts for both correlated and uncorrelated horizontal pleiotropy and relies on a likelihood framework for effective and scalable inference.

**Table 1.**
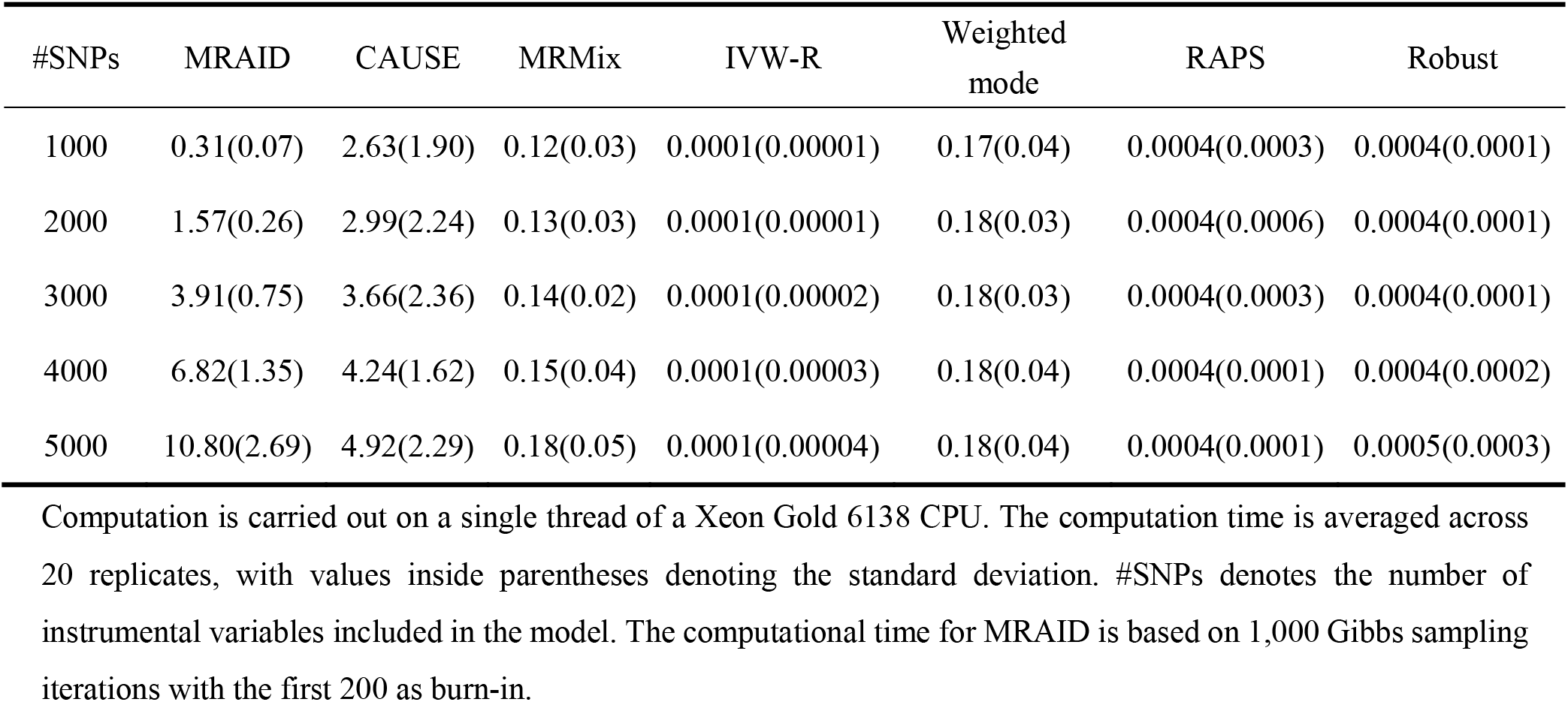
Mean computational time (in minutes) of various two-sample MR methods.

| #SNPs | MRAID | CAUSE | MRMix | IVW-R | Weighted<br>mode | RAPS | Robust |
| --- | --- | --- | --- | --- | --- | --- | --- |
| 1000 | 0.31(0.07) | 2.63(1.90) | 0.12(0.03) | 0.0001(0.00001) | 0.17(0.04) | 0.0004(0.0003) | 0.0004(0.0001) |
| 2000 | 1.57(0.26) | 2.99(2.24) | 0.13(0.03) | 0.0001(0.00001) | 0.18(0.03) | 0.0004(0.0006) | 0.0004(0.0001) |
| 3000 | 3.91(0.75) | 3.66(2.36) | 0.14(0.02) | 0.0001(0.00002) | 0.18(0.03) | 0.0004(0.0003) | 0.0004(0.0001) |
| 4000 | 6.82(1.35) | 4.24(1.62) | 0.15(0.04) | 0.0001(0.00003) | 0.18(0.04) | 0.0004(0.0001) | 0.0004(0.0002) |
| 5000 | 10.80(2.69) | 4.92(2.29) | 0.18(0.05) | 0.0001(0.00004) | 0.18(0.04) | 0.0004(0.0001) | 0.0005(0.0003) |

Computation is carried out on a single thread of a Xeon Gold 6138 CPU. The computation time is averaged across 20 replicates, with values inside parentheses denoting the standard deviation. #SNPs denotes the number of instrumental variables included in the model. The computational time for MRAID is based on 1,000 Gibbs sampling iterations with the first 200 as burn-in.

### Simulations: Type I error control

We evaluated the performance of MRAID and compared it with six existing MR methods in simulations (details in Materials and Methods). We first examined type I error control of different methods in different scenarios. In the absence of both correlated and uncorrelated horizontal pleiotropic effects, most methods, including MRAID, IVW-R, Robust, RAPS and MRMix, all yield reasonably calibrated type I error control (Fig. 2A). Weighted mode and CAUSE, on the other hand, display overly conservative type I error control, which is consistent with the original studies(*7, 8, 20*). The null *p*-value distributions from different methods remain largely similar regardless of the number of SNPs that affect the exposure (Fig. S1A) and their total effects on the exposure (Fig. S1B). We further examined the robustness of different methods in settings where the SNP effects on the exposure do not follow a simple normal distribution but with some SNPs displaying larger effects than the others. In these settings, MRAID, IVW-R and RAPS remain calibrated, while both MRMix and Robust method show inflated type I errors, presumably due to their restricted normality assumptions on the SNP effect sizes (Fig. 2B). Note that we directly used correlated SNPs for MRAID but performed clumping to select independent SNPs for the other methods. Without clumping, all other MR methods produce overly inflated type I errors (Fig. S2).

**Fig. 2.**
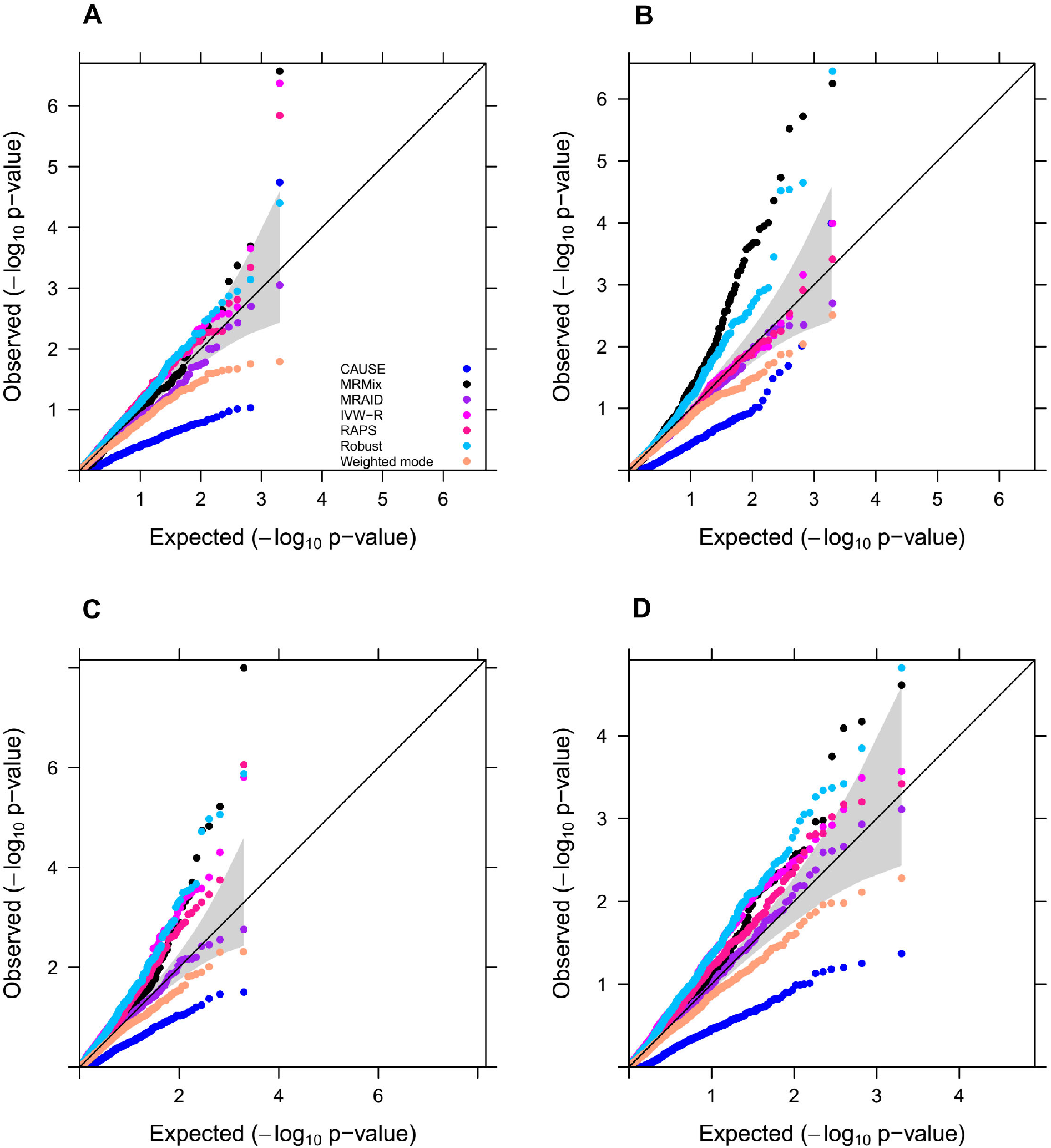
Type I error control of different MR methods in simulations. Type I error is evaluated by quantile-quantile plots of -log10 p-values from different MR methods on testing the causal effect under the null simulations. Compared methods include CAUSE (blue), MRMix (black), MRAID (purple), IVW-R (magenta), RAPS (deep pink), Robust (deep sky blue), Weighted mode (light salmon). Four null simulation scenarios are examined. (**A**) Null simulations in the absence of both correlated and uncorrelated horizontal pleiotropic effects. We simulated 100 instrumental SNPs with their effect sizes drawing from a normal distribution. (**B**) Null simulations in the absence of both correlated and uncorrelated horizontal pleiotropic effects. We simulated 1,000 instrumental SNPs with their effect sizes drawing from a BSLMM distribution with 1% SNPs having large effects and 99% SNPs having small effects. **(C)** Null simulations in the absence of correlated horizontal pleiotropic effect but in the presence of uncorrelated horizontal pleiotropic effect (*PVE*_u_ = 5%). We simulated 100 instrumental SNPs and set the proportion of instrumental SNPs having uncorrelated horizontal pleiotropy to be 20%. **(D)** Null simulations in the presence of both correlated (*π*_c_ = 5%, 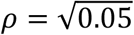) and uncorrelated horizontal pleiotropic effects (*PVE*_u_ = 5%). We simulated 100 instrumental SNPs and set the proportion of instrumental SNPs having the uncorrelated horizontal pleiotropy effect to be 20%.

We examined the effects of horizontal pleiotropy on type I error control for different methods. When horizontal pleiotropic effects are present but are uncorrelated with the instrumental effects, MRAID maintains type I error control (Fig. 2C). In contrast, both Weighted mode and CAUSE remain overly conservative, while MRMix, Robust, IVW-R and RAPS yield inflated *p*-values (Fig. 2C). Similar conclusion holds regardless of the effect size for the uncorrelated horizontal pleiotropy or the proportion of SNPs that display uncorrelated pleiotropic effects (Fig. S3). The *p*-value inflation problem of MRMix relieves when the proportion of SNPs that display uncorrelated horizontal pleiotropic effects decreases. When correlated horizontal pleiotropic effects are also present in addition to the uncorrelated horizontal pleiotropic effects, MRAID maintains effective type I error control (Fig. 2D). In contrast, both Weighted mode and CAUSE remain overly conservative, while MRMix, Robust, IVW-R and RAPS produce inflated *p*-values. Similar conclusion holds regardless of the effect size of the correlated horizontal pleiotropy (Fig. S4A *vs* Fig. S4C), the proportion of SNPs that display uncorrelated horizontal pleiotropic effects (Fig. S4A *vs* Fig. S4B), the proportion of SNPs that display correlated horizontal pleiotropic effects (Fig. S4A *vs* Fig. S4D), or how the correlated horizontal pleiotropic effects are created (Fig. S5).

### Simulations: Power comparison

We examined the power of different MR methods to detect non-zero causal effect. Because the same *p*-value from different methods may correspond to different type I errors, we computed power based on an false discovery rate (FDR) of 0.05 instead of a nominal *p*-value threshold to allow for fair comparison among methods. In the absence of both uncorrelated and correlated horizontal pleiotropic effects, MRAID, IVW-R and RAPS all have higher power than the other methods across different scenarios. Among these three methods, MRAID is slightly more powerful than the other two when the instrumental effects are small or when the causal effect is small (Fig. 3A, 3B), presumably due to the automated instrument selection procedure employed in MRAID. MRAID is slightly less powerful than the other two when the instrumental effects are large and the causal effect is large (Fig. S6), as the simple instrumental selection approaches used in the other methods can be effective in these lesser challenging settings. The performance of these three methods is generally followed by Robust. While Weighted mode, MRMix, and, to a lesser extent, CAUSE, have low power.

**Fig. 3.**
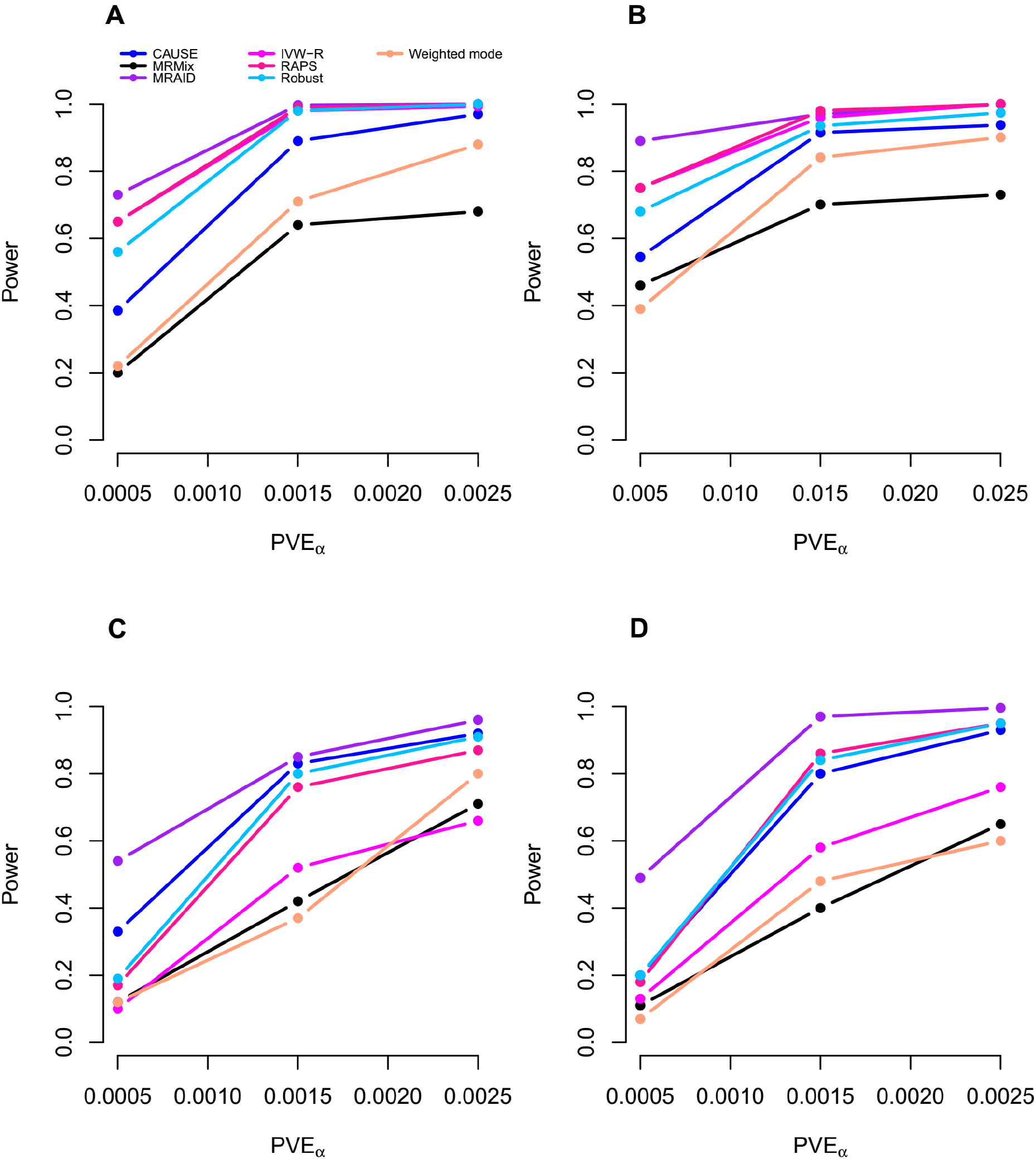
Power of different MR methods in simulations. Power (y-axis) at a false discovery rate of 0.05 to detect the causal effect is plotted against different causal effect size characterized by *PVE*_α_ (x-axis). Compared methods include CAUSE (blue), MRMix (black), MRAID (purple), IVW-R (magenta), RAPS (deep pink), Robust (deep sky blue), Weighted mode (light salmon). Four alternative simulation scenarios are examined. (**A**) Simulations in the absence of both correlated and uncorrelated horizontal pleiotropic effects. We simulated 100 instrumental SNPs with their effects size drawing from a normal distribution. (**B**) Simulations in the absence of both correlated and uncorrelated horizontal pleiotropic effects. We simulated 1,000 instrumental SNPs with their effects size drawing from a BSLMM distribution with 1% SNPs having large effects and 99% SNPs having small effects. **(C)** Simulations in the absence of correlated horizontal pleiotropic effect but in the presence of uncorrelated horizontal pleiotropic effect (*PVE*_u_ = 5%). We simulated 100 instrumental SNPs and set the proportion of instrumental SNPs having the uncorrelated horizontal pleiotropy effect to be 30%. **(D)** Simulations in the presence of both correlated (*π*_c_ = 5%, 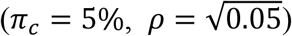) and uncorrelated horizontal pleiotropic effects (*PVE*_u_ = 5%). We simulated 100 causal instrumental SNPs and set the proportion of instrumental SNPs having the uncorrelated horizontal pleiotropy effect to be 20%.

We examined the influence of horizontal pleiotropy on the power of different methods. When horizontal pleiotropic effects are present but are uncorrelated with the instrumental effects, MRAID is more powerful than the other MR methods (Fig. 3C). The power gain brought by MRAID becomes more apparent with increasing horizontal pleiotropy, which is characterized by increased horizontal pleiotropic effect sizes and/or increased proportion of SNPs that display horizontal pleiotropic effects (Fig. S7). The performance of MRAID is often followed by RAPS, Robust, CAUSE, and IVW-R, while MRMix and Weighted mode generally have low power (Fig. S7). Among these methods, the performance of IVW-R is particularly sensitive to the horizontal pleiotropic effect sizes or the proportion of SNPs that display horizontal pleiotropic effects. When correlated horizontal pleiotropic effects are also present in addition to the uncorrelated horizontal pleiotropic effects, the power of MRAID remains higher than the other methods. The higher power of MRAID maintains regardless of the correlated horizontal pleiotropic effect sizes, the proportion of instrumental SNPs that display correlated horizontal pleiotropic effects (Fig. 3D, Fig. S7D-F), or how the correlated horizontal pleiotropic effects are created (Fig. S8). The power gain brought by MRAID is particularly apparent with increased proportion of instrumental SNPs that display uncorrelated horizontal pleiotropic effects (Fig. 3D *vs* Fig. S7F, Fig. S7E *vs* Fig. S7D). Importantly, the power of MRAID is close to an oracle MR approach that uses the actual set of instrumental SNPs for MR inference, especially when the casual effect size is large, supporting the effectiveness of the automatic instrument selection procedure in MRAID (Fig. S9).

Next, we examined the ability of different MR methods in distinguishing the causal effect direction through reverse causality analysis. In particular, we tested the causal effect of the outcome on the exposure in the alternative simulations where the exposure had casual effect on the outcome but not vice versa. In the presence of horizontal pleiotropy, the SNP instruments obtained for the outcome in the reverse MR analysis would contain two sets of SNPs: a set of exposure SNP instruments that are indirectly associated with the outcome through the exposure and a set of SNPs that are directly associated with the outcome thought their horizontal pleiotropic effects on the outcome. Because the two sets of SNPs displayed heterogeneous effects on the exposure, we would fail to detect a non-zero causal effect of the outcome on the exposure. Therefore, the reverse causality analysis in the presence of horizontal pleiotropy effectively served as analysis on null simulations. Indeed, we found that MRAID provides effective type I error control and calibrated *p*-values in the reverse causality analysis across a range of simulation scenarios (Fig. S10 and S11). In contrast, the type I error control of the other methods is highly dependent on the extent of the horizontal pleiotropy. Specifically, when a small proportion of exposure instrumental SNPs display horizontal pleiotropy on the outcome, the majority of the candidate instrumental SNPs for the outcome in the reverse causality analysis would not display horizontal pleiotropic effects on the exposure. In this case, both CAUSE and Weighted mode remain overly conservative, while IVW-R, MRMix, RAPS and Robust yield slightly inflated *p*-values (Fig. S10A, S10C, S11A, and S11C). By contrast, when a large proportion of instrumental SNPs for the exposure display horizontal pleiotropic effects on the outcome, the majority of the candidate instrumental SNPs for the outcome in the reverse causality analysis would display horizontal pleiotropic effects on the exposure. In this case, MRMix, Robust, IVW-R and RAPS all start to produce inflated *p*-values (Fig. S10B and S11B) as we have shown in the corresponding null scenarios. The *p*-value information of these methods becomes more prominent with smaller horizontal pleiotropic effect sizes, where it becomes increasingly hard to select the second set of SNPs to serve as outcome instruments (Fig. S10D and S11D).

Finally, MRAID produces reasonably unbiased causal effect estimates under the null (Fig. S12A) and under various alternatives (Fig. S12B-D).

### Real data applications

We applied MRAID and the other MR methods to analyze 38 lifestyle risk factors and 11 CVD-related traits in the UK Biobank (details in Materials and Methods). Specifically, we divided the UK Biobank data into two separate, equal-sized subsets, representing an exposure GWAS and an outcome GWAS. We performed two sets of analysis. First, we focused on the eight CVD-related traits and examined the causal effect of each trait in the exposure GWAS on the same trait in the outcome GWAS, effectively examining the causal effect of the trait on itself. The true causal effect in such analysis is non-zero and equals exactly one. In the analyses, we found that all methods were able to detect a non-zero causal effect for the trait on itself across all eight CVD-related traits (Fig. 4). However, only MRAID and CAUSE were able to produce 95% confidence intervals that cover the true causal effects for all eight trait pairs, with CAUSE producing confidence intervals that are 2.39-5.69 times larger than MRAID (Fig. 4). For example, in the HDL-HDL analysis, MRAID (estimate = 0.98; 95% CI: 0.96-1.01), CAUSE (0.95; 0.82-1.09) and MRMix (0.96; 0.90-1.02) correctly inferred the causal effect, with MRAID providing the smallest confidence interval (Fig. 4H). In contrast, the confidence intervals from the other four methods did not cover the true causal effect of one. In the LDL-LDL analysis, MRAID (0.97; 0.94-1.01) and CAUSE (0.96; 0.84-1.08) correctly inferred the causal effect, with MRAID providing a smaller confidence interval (Fig. 4F). While the confidence intervals from the other five methods also did not cover the true causal effect of one. The results suggest that both MRAID and CAUSE can produce accurate causal effect estimates and calibrated confidence intervals for trait on itself analysis, with MRAID being more powerful than CAUSE.

**Fig. 4.**
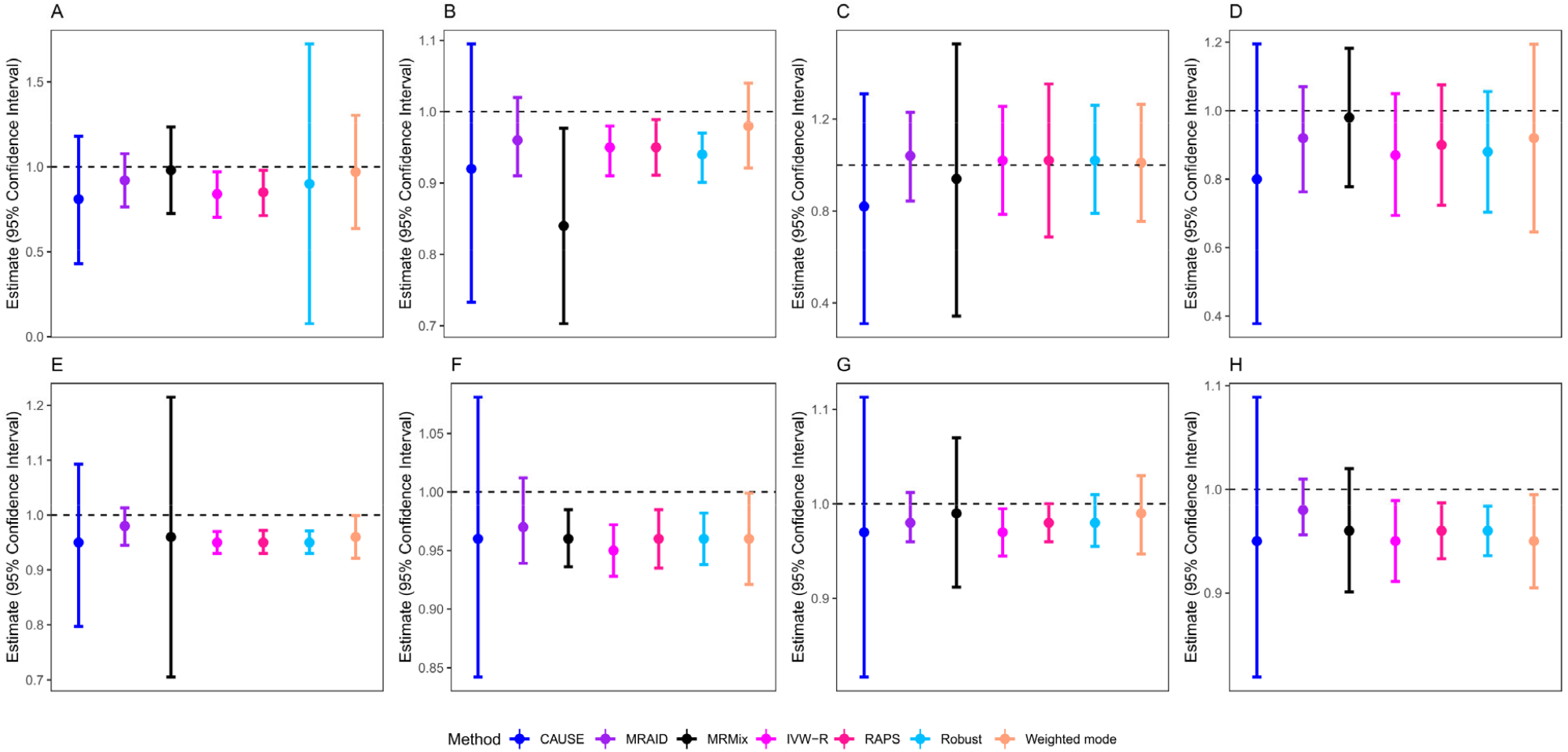
Point estimates and 95% confidence intervals from different MR methods in the trait on itself analysis in the real data. Compared methods include CAUSE (blue), MRMix (black), MRAID (purple), IVW-R (magenta), RAPS (deep pink), Robust (deep sky blue), Weighted mode (light salmon). Analyzed trait pairs include SBP-SBP (**A**), BMI-BMI (**B**), DBP-DBP (**C**), Pulse rate-Pulse rate (**D**), TC-TC (**E**), LDL-LDL (**F**), TG-TG (**G**), and HDL-HDL (**H**). The horizontal black dashed line in each panel represents the true causal effect size of *α*=1. Both MRAID and CAUSE can produce 95% confidence intervals that cover the true causal effects of all trait pairs, with CAUSE producing much larger confidence intervals than MRAID.

Next, we investigated the causal relationship between 38 lifestyle risk factors and 11 CVD-related traits. The association of lifestyle risk factors on CVD-related traits has been extensively documented(*21, 22*). However, it remains controversial on whether the detected associations are causal as some of the association effects were estimated to have different signs in different studies(*23, 24*). We performed both forward causality analysis examining the causal effects of lifestyle factors on CVD-related traits and reverse causality analysis examining the causal effects of CVD-related traits on lifestyle factors. The distribution of *p*-values for the analyzed trait pairs from different methods are shown in Fig. 5A. Consistent with the simulations, we found that the *p*-values from MRAID (genomic inflation factor, GIF =0.90), and to a lesser extent MRMix (GIF = 0.78), are generally well behaved and slightly conservative across analyzed trait pairs, more so than the other methods (Fig. 5A). Also consistent with the simulations, we found that the *p*-values from CAUSE are overly conservative (GIF = 0.12), while the *p*-values from RAPS (GIF = 1.96), Weighted mode (GIF = 1.70), IVW-R (GIF = 2.12), and Robust (GIF = 2.00) all show appreciable inflation (Fig. 5A). Indeed, only MRAID produces calibrated *p*-values in the permutation analysis where we permuted the outcome trait (Fig. 5B).

**Fig. 5.**
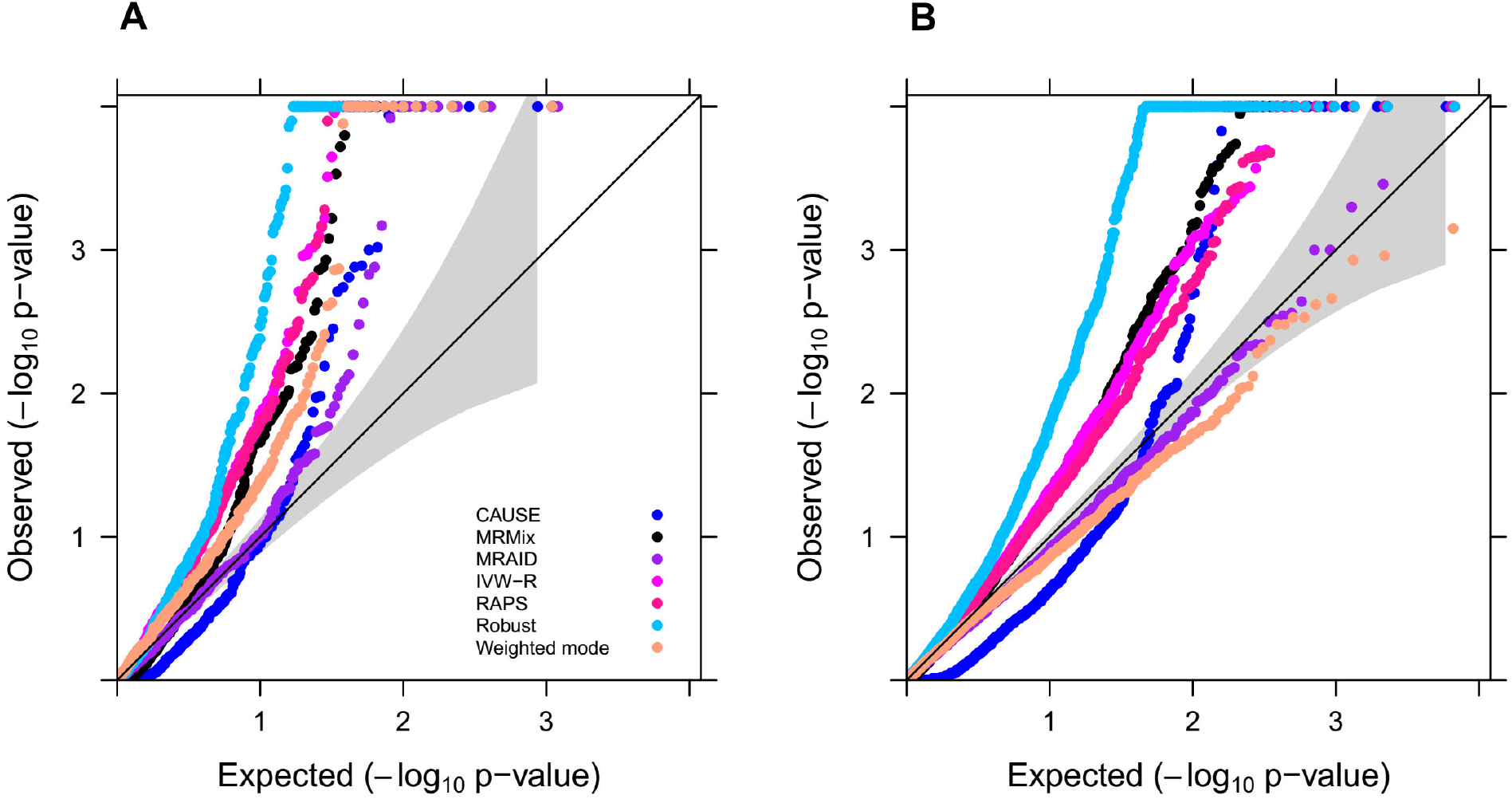
Quantile-quantile plot of -log10 p-values from different MR methods on testing the causal relationship between lifestyle risk factors and CVD-related traits in UK Biobank. Compared methods include CAUSE (blue), MRMix (black), MRAID (purple), IVW-R (magenta), RAPS (deep pink), Robust (deep sky blue), and Weighted mode (light salmon). The results are shown for all 645 analyzed trait pairs **(A)** and the empirical null where we permuted the outcome ten times in the MR analysis of lifestyle traits on CVD-related traits **(B)**.

Based on a Bonferroni corrected *p*-value threshold (8 × 10^−5^), MRAID detected eight causal associations (Table S1), all of which have strong biological support. For example, MRAID detected a negative causal effect of smoking on BMI. The negative association between smoking and obesity has been well documented in observational studies(*25, 26*) and MR studies(*27*). Specifically, nicotine intake during smoking decreases resting metabolic rate(*28, 29*) and inhibits lipoprotein lipase activity and other kinase pathways to reduce lipolysis(*26*), all of which lead to a reduction in the net energy storage in adipose tissues and subsequent weight loss(*30*). Nicotine also activates acetylcholine receptors in the hypothalamus and subsequently anorexigenic neurons(*31, 32*), which leads to suppressed appetite and food intake. As another example, MRAID detected an effect of age started smoking in the former smokers on HDL, suggesting a negative effect of smoking behavior on HDL. Smoking behavior in general is well known to be causally associated with HDL(*33*). In particular, smoking can modify the activity of critical enzymes for lipid transport, lower lecithin-cholesterol acyltransferase (LCAT) activity, and alter cholesterol ester transfer protein (CETP) and hepatic lipase activity, all of which can reduce HDL metabolism. In addition, smoking induces oxidative modifications that render HDL dysfunctional and deprive its atheroprotective properties(*34, 35*).

Importantly, MRAID did not mistakenly detect many false causal associations that were detected by the other methods. A well-known example of a potential false causal association is the effect of smoking on blood pressure. A negative association between smoking and blood pressure has been observed in observational studies(*22*). However, multiple subsequent MR studies on large datasets did not support a causal relationship between the two traits(*33, 36*). Indeed, the association between smoking and blood pressure in observational studies is likely confounded by factors that include, but not limited to age, BMI, social class, salt intake, drinking habits, as well as unmeasured confounders(*37*). Consistent with these previous MR studies, MRAID did not detect a significant causal effect from any of the eight smoking related traits on either SBP or DBP. In contrast, almost all other methods falsely detected causal effects of some of the smoking related traits on blood pressure. For example, the causal effect of the number of unsuccessful stop-smoking attempts on SBP is not detected by MRAID (*p* = 0.44), CAUSE (*p* = 0.01) nor Weighted mode (*p* = 1.3 × 10^−4^), but falsely identified by IVW-R (*p* = 1.4 × 10^−^), Robust (*p* = 4.1 × 10^− 4^), RAPS (*p* = 5.5 × 10^−^), and MRMix (*p* = 2.8 × 10^−^). Similarly, the causal effect of age started smoking in former smokers on SBP is not detected by MRAID (*p* = 0.06) nor CAUSE (*p* = 1.3 × 10^−^), but falsely detected by IVW-R (*p* = 7.8 × 10^−5^), Robust (*p* = 1.5 × 10^−^), RAPS (*p* = 8.9 × 10^−^), Weighted mode (*p* = 1.7 × 10^−^), and MRMix (*p* = 4.1 × 10^−^). As another false example, BMI is unlikely to causally influence the time spent driving, at least not positively. Indeed, MRAID (*p* = 0.01), along with MRMix (*p* = 0.12), CAUSE (*p* = 0.02) and Weighted mode (*p* = 0.04), did not detect any causal effect of BMI on time spent driving. However, both IVW-R (*p* = 2.5 × 10^−^) and RAPS (*p* = 3.1 × 10^−5^) detected a false positive effect of BMI on time spent driving.

Finally, we note that an important feature of MRAID is its ability to effectively decompose the SNP effects on the outcome into three distinct paths: one directly acts from SNPs to the outcome, one mediated through the exposure, and the other acts through a hidden confounding factor that influences both exposure and outcome. Consequently, MRAID can be used to estimate the proportion of SNPs in different categories, including the proportion of SNPs that are associated with the exposure among the genome-wide significant ones (*π*_*β*_), the proportion of SNPs that exhibit correlated horizontal pleiotropy (*π*_c_), the proportion of SNPs that exhibit uncorrelated horizontal pleiotropy among the selected instruments (*π*_l_), and the proportion of SNPs that exhibit uncorrelated horizontal pleiotropy among the remaining candidate instruments (*π*). In the real data applications, we estimated the mean of *π*_*β*_, *π*_c_, *π*_l_ and *π* across the 645 analyzed trait pairs to be 14.6%, 6.4%, 16.4%, and 5%, respectively (Fig. S13). In addition, we estimated their means in the eight significant trait pairs to be 6.2%, 5.7%, 11.4%, and 0.1%, respectively. The proportion of SNPs displaying correlated pleiotropy is also highly correlated with the proportion of SNPs displaying uncorrelated pleiotropy, with the latter generally being larger than the former (Fig. S14). These proportion estimates support the wide-spread horizontal pleiotropy previously identified in complex trait analysis(*15*) and provide detailed quantifications on the extent to which the two types of horizontal pleiotropy influence MR analysis.

## Discussion

We have presented MRAID, a two-sample MR method that can automatically select suitable instruments from a candidate set of correlated SNPs and that can control for both correlated and uncorrelated horizontal pleiotropy in a likelihood-based inference framework. Overall, by automatically selecting instrumental SNPs and performing inference under a likelihood-based framework, MRAID yields calibrated *p*-values across a wide range of scenarios and improves power of MR analysis over existing approaches. We have illustrated the benefits of MRAID through simulations and applications to complex trait analysis.

We have primarily focused on modeling quantitative traits with MRAID in the present study. For binary exposures and outcomes, one could treat them as continuous variables and directly applied MRAID for MR analysis. Treating binary exposures and outcomes as continuous variables can be justified by recognizing the linear model as a first-order Taylor approximation to a generalized linear model such as the logistic regression(*38*). However, such approximation is accurate only when the SNP effects on the exposure and outcome are relatively small. While similar approaches have been applied in many previous MR studies(*39-41*), we caution that the interpretation of the causal effect estimate can be challenging when the linear models are used to fit binary exposures and outcomes, especially when a two-stage inference procedure is used for MR analysis(*42, 43*). For example, when a binary exposure is a dichotomization of a continuous risk factor, the causal effect estimation through modeling the binary exposure without the underlying continuous risk factor may require additional modeling assumptions, even when the main MR assumptions are satisfied. In addition, modeling binary exposure without the underlying continuous risk factor can lead to violation of the exclusion restriction assumption, as the instruments can influence the outcome via the continuous risk factor even if the binary exposure does not change. Therefore, extending MRAID to explicitly model data types beyond quantitative traits is important to ensure its wide applicability. Because MRAID builds upon a data generative model and performs inference on the SNP-exposure model and the SNP-outcome model jointly through a maximum likelihood-based framework, it can be naturally extended towards modeling other types of exposure or outcome data, through, for example, a generalized linear model framework. To the best of our knowledge, the only likelihood-based MR method that accommodates both binary risk factors and outcome is IV-MVB(*44*). IV-MVB, however, requires individual-level data, applies to the one-sample analysis setting, and cannot easily handle multiple instruments in a computationally efficient fashion especially for those that are correlated. Therefore, exploring the benefits of MRAID extensions towards modeling generalized data types while keeping computation in check will be an important direction for future research.

MRAID is not without limitations. First, while MRAID performs automated selection on SNP instruments, such selection builds upon a sparsity inducing modeling assumption specified on the SNP effect sizes. The sparse modeling assumption contains multiple hyper-parameters that rely on a sampling-based algorithm for inference. Accurate and robust inference of the hyper-parameters will likely require at least a moderate number of candidate instruments. While the significance of the trait pairs evaluated by MRAID in our real data application does not appear to be dependent on the number of candidate instruments selected for the trait pair (Fig. S15), we caution that MRAID may incur low power when the instrumental effect size is small and the number of candidate instruments is low, which can happen in GWAS with small sample sizes and for exposure traits with a non-polygenic architecture. Second, MRAID primarily follows the approach of CAUSE to model correlated horizontal pleiotropy by introducing a single latent variable to serve as the confounder for both the exposure and the outcome. Because of its limitation in modeling only a single unobserved confounding factor, MRAID may not be fully effective in settings where multiple or other types of shared genetic components are present between the exposure and the outcome. Finally, the summary statistics version of MRAID requires as input two LD matrices, one from the exposure GWAS and another from the outcome GWAS. In the present study, we have estimated both LD matrices using individual level data. In the absence of individual level data, both LD matrices may be estimated from a reference panel with the same genetic ancestry (e.g. from the 1,000 Genomes Project). However, care needs to be taken when the exposure and outcome GWASs are carried out on two populations with distinct genetic ancestries.

## Materials and Methods

### MRAID for individual level data

We provide an overview of our method here, with its inference and technical details provided in the Supplementary Text and an illustrative diagram displayed in Fig. 1. Our goal is to estimate and test the causal effect of an exposure variable on an outcome variable in the two-sample MR setting where the exposure and outcome variables are measured in two separate GWASs with no sample overlap. We refer to the two separate GWASs as the exposure GWAS and the outcome GWAS, respectively. To set up the notations, we denote **x** as an *n*_l_ -vector of the exposure variable measured on *n*_l_ individuals in the exposure GWAS. We denote **y** as an *n*_2_-vector of the outcome variable measured on *n*_2_ individuals in the outcome GWAS. We scale both **x** and **y** to have zero mean and unit standard deviation. In the exposure GWAS, we perform an initial screening to select SNPs that are associated with the exposure variable with a marginal *p*-value below the genome-wide significance threshold of 5 × 10^−8^. These SNPs are likely in LD with each other and are selected to serve as the initial set of candidate instruments. We denote **Z**_x_ as the resulting *n*_l_ by *p* genotype matrix for the *p* selected candidate instrumental SNPs in the exposure GWAS. We also denote **Z** as an *n*_2_ by *p* genotype matrix for the same *p* candidate instrumental SNPs in the outcome GWAS. We scale each column of the two genotype matrices to have mean zero and standard deviation of one. We model the relationship among the exposure, outcome and genotypes through the following three linear regressions:

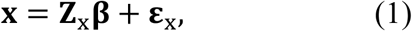

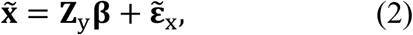

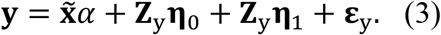

Above, equation (1) describes the relationship between the genotypes **Z**_x_ and the exposure variable **x** in the exposure GWAS; equation (2) describes the relationship between the genotypes **Z**_y_ and the unobserved exposure 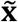 in the outcome GWAS; equation (3) describes the relationship among the genotypes **Z**_y_, the outcome **y**, and the unobserved exposure 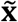 in the outcome GWAS; **β** is a *p*-vector of SNP effects on the exposure; both **η**_0_ and **η**_l_ are *p*-vectors of horizontal pleiotropy effects on the outcome; *α* is a scalar that represents the causal effect of the exposure on the outcome; **ε**_x_ is an *n*_l_-vector of residual error with each element independently and identically distributed from a normal distribution 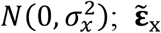 is an *n*_2_ -vector of residual error with each element distributed from the same normal distribution 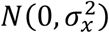; and **ε**_y_ is an *n*_2_-vector of residual error with each element distributed from a normal distribution 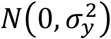. We note that while the above three equations are specified based on two separate GWASs, they are connected to each other by the common parameter **β**. We carefully consider the modeling assumptions on the SNP effects on the exposure variable **β** as well as the horizontal pleiotropic effects **η**_0_ and **η**_l_ as follows.

The *p* SNPs included in the above model represent an initial set of candidate instruments. While all the candidate instruments are marginally associated with the exposure, the majority of them are unlikely the causal SNPs for the exposure variable. Instead, most candidate instruments likely represent tagging SNPs that are associated with the exposure variable due to LD with the truly causal ones underlying the exposure. Therefore, it would be beneficial to perform additional selections on the candidate instruments to identify SNPs that are causal for the exposure and treat them as the instruments in order to maximize the power of MR analysis. To do so, we borrow ideas from fine-mapping approaches developed in the research field of GWAS and specify a sparsity inducing modeling assumption on the SNP effects on the exposure (**β**) to perform automated instrument selection. In particular, we assume that 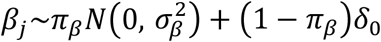, where *δ*_0_ is the Dirac function that represents a point mass at zero. That is, with probability 1 − *π*_*β*_, the *j*-th SNP has zero effect on the exposure; while with probability *π*_*β*_, the *j*-th SNP has a non-zero effect on the exposure and its effect size follows a normal distribution with mean zero and variance 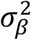, where the variance parameter 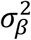 determines the magnitude of the effect sizes. The sparse assumption on **β** allows us to select SNPs with non-zero effects on the exposure to serve as the instruments in the MR model.

In addition, the *p* SNPs included in the above model can also exhibit horizontal pleiotropic effects and influence the outcome variable through pathways other than the exposure. To control for the potential horizontal pleiotropic effects and improve causal effect inference, we introduce two sets of parameters, **η**_0_ and **η**_l_, to model horizontal pleiotropic effects. The two sets of parameters are placed separately for the two SNP groups – the group of selected instrumental SNPs and the group of unselected non-instrumental SNPs – that are categorized by the sparse modeling assumption on **β**. In particular, **η**_l_ represents the horizontal pleiotropic effects exhibited by the selected SNPs instruments with non-zero **β** while **η**_0_ represents the horizontal pleiotropic effects exhibited by the unselected non-instrumental SNPs with zero **β**. Controlling for **η**_l_ can help mitigate the bias in causal effect estimation induced by horizontal pleiotropic effects from the instrumental SNPs. While controlling for **η**_0_ can reduce residual error variance in equation (3) and thus help improve the statistical efficiency of causal effect estimation.

To effectively control for the horizontal pleiotropic effects exhibited from both SNP groups, we specify separate modeling assumptions on **η**_0_ and **η**_l_. Specifically, for the selected SNP instruments, we assume that they can exhibit horizontal pleiotropic effects in two different ways: they can affect the outcome through a common confounder that is associated with both the exposure and outcome, and they can affect the outcome through paths independent of the exposure. For the first type of horizontal pleiotropy, we assume that each selected SNP instrument has a probability of *π*_c_ to induce pleiotropy through the confounder. Following(*8*), we assume that the confounder effect on the outcome is *ρ* times its effect on the exposure. Consequently, the effect of the selected SNP instrument acted through the confounder on the outcome becomes *ρβ*_j_, if the SNP effect on the exposure is *β*_j_. Thus, our assumption on 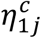, which represents the first type of horizontal pleiotropy as a part of **η**_l_ for the *j*-th SNP, is 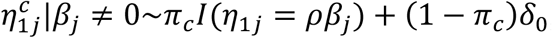, where *I*(⋅) is an indicator function that sets the horizontal pleiotropic effect to be *ρβ*_j_. For the second type of horizontal pleiotropy, we assume that each selected SNP instrument has a probability of *π*_l_ to exhibit a horizontal pleiotropic effect on the outcome directly, bypassing the exposure. We use 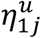 to represent the second type of horizontal pleiotropy as a part of **η**_l_ for the *j*-th SNP. Our assumption on 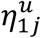 is thus 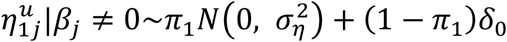, where the variance 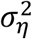 determines the strength of the horizontal pleiotropic effect. Note that the first type of horizontal pleiotropic effects are correlated with the instrumental effects on the exposure due to the confounder, while the second type of horizontal pleiotropic effects are uncorrelated with the instrumental effects on the exposure. The total horizontal pleiotropy is the summation of the two, with 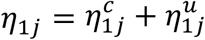. Certainly, because **η**_l_ are the horizontal pleiotropic effects for the selected SNP instruments, we have *η*_lj_ = 0 if *β*_j_ = 0. For the unselected non-instrumental SNPs with a zero *β*_j_, we assume that 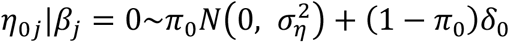. That is, with probability *π*_0_, the non-instrumental SNPs display horizontal pleiotropic effects characterized by the same variance parameter 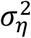. We use the same variance parameter 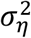 for modeling the uncorrelated horizontal pleiotropic effects from both instrumental and non-instrumental SNPs because we often do not have enough number of SNPs to estimate two separate parameters accurately. Since **η**_0_ are the horizontal pleiotropic effects for the non-instruments, we also have *η*_0j_ = 0 if *β*_j_ ≠ 0.

The above parameterization of the horizontal pleiotropic effects is based on the selection of SNP instruments. An equivalent and alternative parametrization of the horizontal pleiotropic effects is to partition them into a correlated horizontal pleiotropic component **η**_C_ and an uncorrelated horizontal pleiotropic component **η**_u_. Specifically, the correlated horizontal pleiotropy occurs only for the selected SNP instruments with *η*_cj_|*β*_j_ ≠ 0∼*π*_c_*I*(*η*_lj_ = *ρβ*_j_) (1 − *π*_c_)*δ*_0_ and *η*_cj_ = 0 if *β*_j_ = 0. The uncorrelated horizontal pleiotropy, on the other hand, occurs for both instrumental and non-instrumental SNPs with 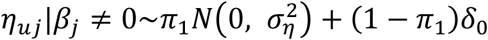 and 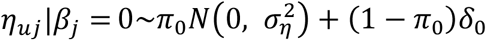. In other words, 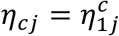 and 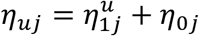.

Overall, the SNP effects on the outcome in our model are exhibited through three different paths: via the exposure on outcome causal effect *α* ; via the correlated horizontal pleiotropic effects mediated by an unobserved confounder; and via the uncorrelated horizontal pleiotropic effects. SNPs in the model can exhibit none, one, or multiple types of these effects. Note that the SNP effects on the outcome through the causal effect and through the correlated horizontal pleiotropy are not distinguishable from each other unless we make further modeling assumptions. Here, following(*8*), we assume *π*_c_ to be small. Thus, among the selected SNP instruments with non-zero effects on the exposure, only a fraction of them exhibit correlated horizontal pleiotropic effects on the outcome (details in Supplementary Text).

Our key parameter of interest is the causal effect *α*. The causal interpretation of *α* in a standard MR model requires the selected SNP instruments to satisfy three conditions: (i) instruments are associated with the exposure (relevance condition); (ii) instruments are not associated with any other confounder that may be associated with both exposure and outcome (independence condition); (iii) instruments only influence the outcome through the path of exposure (exclusion restriction condition). Our modeling assumption on **β** allows us to select SNPs to satisfy the relevance condition. Our modeling assumptions on **η**_0_ and **η**_l_ allow us to explicitly model the violation of the independence and exclusion restriction conditions. Therefore, our model effectively replaces the general conditions (ii) and (iii) with specific modeling assumptions on **β**, **η**_0_ and **η**_l_. In addition, through explicit modeling of the correlation between the instrument-exposure effects and instrument-outcome effects through *ρ*, our model no longer requires the InSIDE assumption, which is sometimes referred to as the weak exclusion restriction condition(*3*). Consequently, the causal effect interpretation of *α* in our model only depends on the explicit assumptions made in the model.

We are interested in estimating the causal effect *α* and testing the null hypothesis *H*_0_ : *α* = 0. Performing inference on *α*, however, is computationally challenging, as the likelihood defined based on the above modeling assumptions is in a complicated form and involves integrations that cannot be obtained analytically. Here, we develop an approximate inference algorithm under the maximum likelihood framework to perform numerical integration of the likelihood and obtain an approximate *p-*value for testing *α*. Our algorithm is based on the observation that the likelihood function of *α* can be expressed as a ratio between the posterior and the prior. Because the posterior of *α* is asymptotically normally distributed(*45, 46*), we can use Gibbs sampling to obtai posterior samples of *α* and use the sample mean and sample standard deviation to summarize this posterior distribution. In addition, we can also specify a normal prior on *α* and obtain the prior mean and standard deviation. Because the likelihood of *α* is expressed as the posterior divided by the prior and is itself asymptotically normally distributed(*45, 46*), we can rely on the method of moments to obtain the approximate maximum likelihood estimate 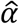 and its standard error 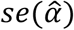 based on the mean and standard deviation from both the posterior and the prior. Afterwards, we can construct an approximate Wald test statistic and obtain a *p*-value for hypothesis testing. Details of the algorithm is provided in the Supplementary Text. Note that, while our algorithm relies on Gibbs sampling, we do not perform a Bayesian analysis; rather, we treat the Gibbs sampling as a convenient and accurate numerical approximation tool to obtain the marginal likelihood of *α*, which is otherwise infeasible or inaccurate to obtain under various frequentist approaches.

We refer to our model and algorithm together as the two-sample Mendelian Randomization with Automated Instrument Determination (MRAID). The automated instrument determination part highlights the desirable feature of our model in automatically selecting instrumental variables from a set of candidate ones that are in potentially high LD with each other. Compared with existing two-sample MR approaches, MRAID relies on a likelihood inference framework, is capable of modeling correlated instruments, performs automated instrument selection, controls for both correlated and uncorrelated horizontal pleiotropy, and is computationally scalable. MRAID is implemented in an R package, freely available at www.xzlab.org/software.html.

### MRAID for summary statistics

While we have described MRAID using individual-level data, MRAID can be extended to make use of only summary statistics. Details for the summary statistics version of MRAID are provided in the Supplementary Text. Briefly, the summary statistics version of MRAID requires two types of input: the SNP marginal effect size estimates on the exposure and outcome; and the SNP correlation matrices in the exposure and outcome GWASs. Both input types are obtained based on standardized genotype data where the genotypes for each SNP have been standardized to have zero mean and unit standard deviation. Here, we denote the *p*-vector of the SNP marginal effect size estimates on the exposure as 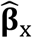 and the corresponding vector of marginal effect size estimates on the outcome as 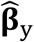. We denote the *p* by *p* SNP correlation matrix in the exposure GWAS as Σ_l_ and the corresponding matrix in the outcome GWAS as Σ_2_. Both Σ_l_ and Σ_2_ are positive semi-definite and can be estimated from the same LD reference panel (e.g. individuals with the same ancestry in the 1,000 Genomes Project). The MRAID model for summary statistics can be constructed based on the following two equations

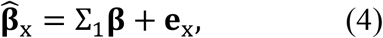

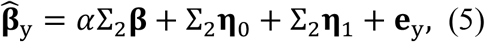

where **e**_**x**_ is a *p*-vector of residual error that follows a multivariate normal distribution 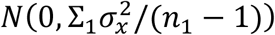; and **e**_y_ is a *p*-vector of residual error that follows another a multivariate normal distribution 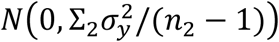. A similar approximate inference algorithm under the maximum likelihood framework is developed for the summary version of MRAID.

#### Simulations

We performed realistic simulations to evaluate the performance of MRAID and compared it with six existing MR methods. For simulations, we randomly selected 60,000 individuals from UK Biobank(*19*). We split these individuals randomly into two equal-sized sets: one set with 30,000 individuals to serve as the exposure GWAS and another set with the remaining 30,000 individuals to serve as the outcome GWAS. For these individuals, we obtained their genotypes from 649,695 SNPs on chromosome 1 that are overlapped with the GERA study we used before(*13*), standardized each SNP to have mean zero and unit standard deviation, and used the standardized genotypes to simulate the exposure and outcome. Specifically, in the exposure GWAS, we randomly selected *K* SNPs (*K* =100 or 1,000) to have non-zero effects on the exposure. We denoted the genotype matrix of the *K* SNPs as 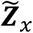. We simulated the *K* SNP effect sizes on the exposure (**β**) from a normal distribution 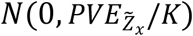, where the scalar 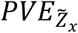 represents the proportion of variance in the exposure variable explained by these genetic effects. We summed the genetic effects across all *K* SNPs as 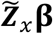. In addition, we simulated the residual errors **ε**_x_ from a normal distribution 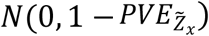. We then summed the genetic effects and the residual errors to yield the simulated exposure variable **x**. In the outcome GWAS, we obtained the genotypes for the same *K* SNPs as 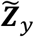 and used the same **β** from the exposure GWAS to compute the genetic component underlying the outcome as 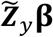. We set the causal effect *α* to be 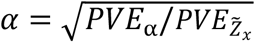, so that the proportion of variance in the outcome variable explained by the causal effect term 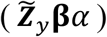 is *PVE*_α_. We randomly obtained *π*_c_*K* SNPs (rounded to an integer) from the *K* SNPs to exhibit correlated pleiotropy.

We simulated the correlated pleiotropic effect sizes to be *ρ***β** and set *ρ* so that the proportion of variance in the outcome variable explained by correlated pleiotropy is *PVE*_C_. In addition, we randomly obtained *π*_l_*K* SNPs (again rounded to an integer) from the *K* SNPs and randomly obtained 100 − *π*_l_*K* SNPs from the remaining non-causal SNPs to exhibit uncorrelated pleiotropy, so that a total of 100 SNPs displayed uncorrelated pleiotropy. We simulated the uncorrelated horizontal pleiotropic effects for these 100 SNPs from a normal distribution and scaled them so that the proportion of phenotypic variance in the outcome explained by uncorrelated pleiotropy is *PVE*_u_. We simulated the residual errors **ε** from a normal distribution *N*(0, 1 − *PVE*_α_ − *PVE*_C_ − *PVE*_u_). We summed the causal effect term, correlated and uncorrelated horizontal pleiotropic effects, and the residual errors to yield the simulated outcome **y**. We treated the causal SNPs as unknown and followed standard MR procedure to select SNPs to serve as the instrumental variables. To do so, we used the linear regression model implemented in GEMMA(*47*) to perform association analysis in the exposure GWAS and selected SNPs with a *p*-value below 5 × 10^−8^ as the candidate instrumental variables for analysis. For the selected SNPs, we obtained their effect size estimates, standard errors, and Z scores to serve as the summary statistics input. We also denoted the standardized genotype matrices for the selected SNPs in the exposure and outcome GWASs as ***Z***_X_ and ***Z***_y_, respectively. Based on the genotype matrices, we obtained the SNP correlation matrices as 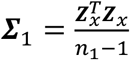 and 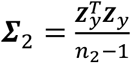 to serve as input for MR model fitting.

In the simulations, we first examined a baseline simulation setting where we set 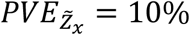, *PVE*_α_ = 0, *K* = 100, *π*_c_ = 0, *PVE*_u_ = 0, *PVE*_c_ = 0. On top of the baseline setting, we varied one parameter at a time to examine the influence of various parameters on method performance. For 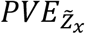, we set it to be either 5% or 10%. For **β**, in addition to simulating it from a normal distribution, we also simulated them from the Bayesian sparse linear mixed model (BSLMM) distribution(*38*). Specifically, we randomly selected either 1% or 10% of the *K* SNPs to have large effects and these large-effect SNPs explain 20% of 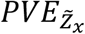. We set the remaining SNPs to have small effects to explain the remaining 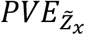. For *K*, we set it to be either 100 or 1,000. For *PVE*_α_, we set it to be zero in the null simulations and examined different values in the alternative simulations. In the alternative simulations, we set *PVE*_α_ to be 0.05%, 0.15% or 0.25% when *K*=100 and set it to be 0.5%, 1.5% and 2.5% when *K*=1,000 to ensure sufficient power. For the uncorrelated horizontal pleiotropic effects, we set *PVE*_u_ to be either 0, 2.5% or 5%. Under the null (*PVE*_α_ = 0) in the absence of uncorrelated horizontal pleiotropy (*PVE*_u_ = 0), we set *K* to be 100 or 1,000. In the presence of uncorrelated horizontal pleiotropy, we set *K* to be 100 and set *π*_l_ to be either 0, 10%, 20%, or 30%. We also simulated the correlated pleiotropy effects and set *π*_c_ to be either 5% or 10%, with *ρ* being 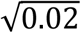 or 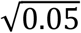 following the previous literature(*8*).

For null simulations, we performed 1,000 simulation replicates in each scenario to examine type I error control. For power evaluation, we performed 100 alternative simulations along with 900 null simulations, with which we computed power based on an FDR of 0.05. We then repeated such analysis five times and report the average power across these replicates. Note that we computed power based on FDR instead of a nominal *p*-value threshold to allow for fair comparison across methods, as the same *p*-value from different methods may correspond to different type I errors.

### Real Data Applications

We applied MRAID and other MR methods to detect causal associations between 38 lifestyle risk factors and 11 CVD-related traits in the UK Biobank. The UK Biobank data consists of 487,298 individuals and 92,693,895 imputed SNPs(*19*). We followed the same sample QC procedure in Neale lab (https://github.com/Nealelab/UK_Biobank_GWAS/tree/master/imputed-v2-gwas) to retain a total of 337,129 individuals of European ancestry for analysis. We also filtered out SNPs with an HWE *p*-value < 10^−7^, a genotype call rate < 95%, or an MAF < 0.001 to obtain a total of 13,876,958 SNPs for analysis. For the retained individuals, we obtained all lifestyle-related quantitative traits and CVD-related traits, removed those traits with a sample size less than 10,000, and focused on the remaining set of 38 lifestyle traits and 11 CVD-related traits for analysis. The 38 lifestyle traits include 8 physical activity traits, 12 alcohol intake traits, 10 diet traits (e.g. coffee and fruits) and 8 smoking related traits. The 11 CVD-related traits include four pulse wave traits, two blood pressure traits (SBP and DBP), four lipid traits (LDL, HDL, TC, and TG) and BMI. Details of these traits are listed in Table S2. Many of these lifestyle risk factors have been found to be associated with CVD-related traits in observational studies(*48-50*), though it remains unclear whether these associations represent causal relationship. For each trait in turn, we removed the effects of sex and top ten genotype principal components (PCs) to obtain the trait residuals, standardized the residuals to have a mean of zero and a standard deviation of one, and used these scaled residuals for MR analysis.

To mimic the two-sample MR design, we randomly split the 337,129 individuals into two non-overlap sets: an exposure GWAS set with 168,564 individuals and an outcome GWAS set with 168,565 individuals. The random data split strategy ensures sample homogeneity within each study and independence between studies, and was extensively used in the previous MR literature(*6, 51-53*). We examined the 38 lifestyle traits in the exposure GWAS and examined the 11 CVD-related traits in the outcome GWAS. In both GWASs, we obtained summary statistics for each trait through linear regression implemented in GEMMA. When lifestyle traits in the exposure GWAS were treated as the exposure, we selected SNPs with a *p*-value below 5 × 10^−8^ to serve as the candidate instruments for each exposure trait. Because almost all MR methods require at least two instrumental SNPs and some methods can become unstable when the number of instrumental SNPs is too large, we removed exposure traits for which the number of candidate instruments is either below two or above 10,000. This way, we removed three traits with less than two candidate instruments and four traits with more than 10,000 candidate instruments. We paired the remaining 31 exposure lifestyle traits with 11 outcome CVD-related traits into 341 trait pairs. The mean number of significantly associated SNPs among the 31 traits is 286. When CVD-related traits in the outcome GWAS were treated as the exposure, we removed three traits with less than two candidate instruments, and found that the remaining eight traits have more than 10,000 candidate instruments. Therefore, for these remaining traits, we used a more stringent *p*-value threshold of 1 × 10^−l5^ to select SNP instruments and analyzed the resulting 304 trait pairs. The mean number of associated SNPs among the eight CVD-related traits is 2,318. In total, we analyzed 645 trait pairs.

### Compared Methods

We compared the performance of MRAID with six existing methods that include the followings. (1) IVW-R, which is the random effects version of IVW. It obtains the causal effect estimate through weighting and combining the effect estimates from individual instruments. It relies on random effects to account for pleiotropy and effect estimate heterogeneity across instruments(*54*). (2) Weighted mode, which is a mode version of IVW. It obtains the causal effect estimate as the mode, instead of the mean, of the effect estimates obtained from individual instruments(*55*). (3) Robust, which is a robust version of IVW. It uses the MM-estimation procedure consisting of an initial S-estimate followed by an M-estimate(*56*) that is further combined with Tukey’s bi-weight loss function(*57*). We fitted methods (1)-(3) using R package ‘MendelianRandomization’ with default settings. (4) RAPS, which is the MR Adjusted Profile Score method. It incorporates random effects and robust loss functions into the profile score to account for systematic and idiosyncratic pleiotropy(*6*). We fitted RAPS using R package ‘mr.raps’; (5) MRMix, which relies on a mixture model to account for horizontal pleiotropic effects and their correlation with instrumental effect sizes(*7*). We fitted MRMix using R package ‘MRMix’. (6) CAUSE, which identifies instrumental effect size patterns that are consistent with causal effects while accounting for correlated pleiotropy(*8*). We fitted CAUSE using R package ‘cause’. We compared MRAID with the above six methods because CAUSE is one of the most recently developed methods; IVW-R, Robust and RAPS all have been shown to have superior performance when the InSIDE assumption is satisfied; while MRMix and Weighted mode perform well even the InSIDE assumption is violated(*8, 20*). In both simulations and real data applications, we first obtained SNPs that achieve genome-wide significance level (*p* < 5 × 10^−8^) to serve as a candidate set of instrumental SNPs. We directly use this candidate set of instrumental SNPs for MRAID. Because all other MR methods require independent instrumental SNPs, we performed LD clumping on the candidate set of instrumental SNPs to select independent ones for analysis. LD clumping is performed using PLINK, where we set the LD *r*^2^ parameter to be 0.001. CAUSE also requires estimating some nuisance parameters in the model by using a random set of SNPs across the genome, and we did so by randomly selecting 100,000 SNPs following(*8*). Finally, we explored an oracle approach in the power simulations where we knew the actual set of instrumental SNPs that affect the exposure variable. In the oracle approach, we obtained the actual set of instrumental SNPs, selected among them the independent ones via pruning, and used the selected set of SNPs to serve as instruments using the IVW-R method. The compared methods and their corresponding software are listed in Table S3.

## Data Availability

All data produced are available online at https://www.ukbiobank.ac.uk/.

## Funding

ZY was supported by the National Natural Science Foundation of China (81872712), the Natural Science Foundation of Shandong Province (ZR2019ZD02) and the Young Scholars Program of Shandong University (2016WLJH23). XZ was supported by the University of Michigan. This study has been conducted using UK Biobank resource under Application Number 51470. UK Biobank was established by the Wellcome Trust medical charity, Medical Research Council, Department of Health, Scottish Government and the Northwest Regional Development Agency. It also has funding from the Welsh Assembly Government, British Heart Foundation and Diabetes UK.

## Author contributions

XZ conceived the idea. XZ and ZY developed the methods. ZY developed the software tool with assistance from LL. ZY performed simulations and real data analysis with assistance from LL, PG, RY and FX. XZ and ZY wrote the manuscript with input from all other authors. All authors reviewed and approved the final manuscript.

## Competing interests

Authors declare that they have no competing interests.

## Data and materials availability

No data are generated in the present study. The UK Biobank data is from UK Biobank resource under application number 51470. The MRAID is implemented in the R package MRAID, freely available on GitHub (https://github.com/yuanzhongshang/MRAID).

## Notes

### Competing Interest Statement

The authors have declared no competing interest.

