## Supplementary materials for "Likelihood based Mendelian randomization analysis with automated instrument selection and horizontal pleiotropic modeling"

**This PDF file includes:**

Supplementary Text
Figs. S1 to S15
Tables S1 to S3

### Supplementary Text

#### 1. MRAID for individual level data

We follow the notations in the main text and rewrite the MRAID model in the following equivalent form:

$$\mathbf{x} = \mathbf{Z}_x\boldsymbol{\beta} + \boldsymbol{\varepsilon}_x, \quad (1)$$

$$\mathbf{y} = \mathbf{Z}_y\boldsymbol{\beta}\alpha + \mathbf{Z}_y(\boldsymbol{\beta} \circ \mathbf{v})\rho + \mathbf{Z}_y\boldsymbol{\eta}_\mathbf{u} + \boldsymbol{\varepsilon}_y, \quad (2)$$

where $\mathbf{v}$ is a $p$-vector of binary indicators that indicate whether the instrumental SNPs display correlated pleiotropy effects ($v_j = 1$) or not ($v_j = 0$); and the term $\boldsymbol{\beta} \circ \mathbf{v}$ represents the Hadamard product, also known as the element wise product, of the two vectors $\boldsymbol{\beta}$ and $\mathbf{v}$. For $\mathrm{j} = 1, \cdots, \mathrm{p}$, we further have:

$$\beta_j \sim \pi_\beta N(0, \sigma_\beta^2) + (1 - \pi_\beta)\delta_0,$$

$$\eta_{uj} \sim \pi_1 N(0, \sigma_\eta^2) + (1 - \pi_1)\delta_0 \text{ if } \beta_j \neq 0,$$

$$\eta_{uj} \sim \pi_0 N(0, \sigma_\eta^2) + (1 - \pi_0)\delta_0 \text{ if } \beta_j = 0,$$

$$p(v_j = 1) = \pi_c, \text{ if } \beta_j \neq 0.$$

Above, we assume that $\sigma_\beta^2$ follows an inverse gamma distribution $\sigma_\beta^2 \sim InvG(a_\beta, b_\beta)$. We set $a_\beta = \frac{p}{10} + 1$and $b_\beta = 0.2$ to ensure a prior mean of $2/p$. Similarly, we assume $\sigma_\eta^2 \sim InvG(a_\eta, b_\eta)$, with $a_\eta = \frac{p}{5} + 1$ and $b_\eta = 0.2$ that ensure a prior mean of $1/p$. We assume the proportion parameters to follow *beta* distributions: $\pi_\beta \sim beta(\lambda_{\beta 1}, \lambda_{\beta 2})$, with $\lambda_{\beta 1} = 0.5$ and $\lambda_{\beta 2} = 4.5$ to ensure a prior mean of approximately 0.1; $\pi_c \sim beta(\lambda_{c1}, \lambda_{c2})$ with $\lambda_{c1} = 0.5$ and $\lambda_{c2} = 9.5$ to ensure a prior mean of approximately 0.05; $\pi_1 \sim beta(\lambda_{21}, \lambda_{22})$ with $\lambda_{21} = 0.5$ and $\lambda_{22} = 1.5$ to ensure a prior mean of 0.25; and $\pi_0 \sim beta(\lambda_{31}, \lambda_{32})$ with $\lambda_{31} = 0.05$ and $\lambda_{32} = 9.95$ to ensure a prior mean of 0.005. These prior means are set to represent our prior belief that a relatively small proportion of SNPs display non-zero effects on the exposure, that a relatively small proportion of SNPs display horizontal pleiotropy, that the selected instrumental SNPs are more likely to display horizontal pleiotropy than the non-selected SNPs, and that horizontal pleiotropic SNPs are more likely to display uncorrelated pleiotropic effects than correlated pleiotropic effects. For the other hyper-parameters, we use relatively non-informative priors. Specifically, for the error variance parameters, we set $\mathrm{p}(\sigma_x^2) \propto 1$ and $\mathrm{p}(\sigma_y^2) \propto 1$. For the parameter $\rho$, we set $\mathrm{p}(\rho) \propto 1$. For the causal effect parameter $\alpha$, we set $\alpha \sim N(0, \sigma_0^2)$ with $\sigma_0^2 \to \infty$.

To facilitate computation, we introduce a $p$-vector of binary indicators $\boldsymbol{\gamma} = (\gamma_1, \ldots, \gamma_p)^T$ to indicate whether each SNP has a non-zero effect ($\beta_j$) on the exposure ($\gamma_j = 1$) or not ($\gamma_j = 0$). We also introduce another $p$-vector of binary indicators $\boldsymbol{\tau} = (\tau_1, \ldots, \tau_p)^T$ to indicate whether each SNP has the horizontal pleiotropy effect ($\tau_j = 1$) or not ($\tau_j = 0$). Thus, we have $p(\gamma_j = 1) = \pi_\beta$, $p(\tau_j = 1 | \gamma_j = 1) = \pi_1$, $p(\tau_j = 1 | \gamma_j = 0) = \pi_0$, and $p(v_j = 1 | \gamma_j = 1) = \pi_c$.

#### 2. Detailed sampling steps and efficient computation

Our goal is to first obtain the posterior samples for the causal effect $\alpha$. To do so, we rely on Gibbs sampling to obtain the conditional distribution for one parameter at a time. Our posterior likelihood is in the form of

$$f(\alpha,\boldsymbol{\beta},\boldsymbol{\gamma},\boldsymbol{\eta}_{\mathbf{u}},\boldsymbol{\tau},\mathbf{v},\rho|\mathbf{x},\mathbf{y})$$
$$\propto f(\mathbf{x},\mathbf{y}|\alpha,\boldsymbol{\beta},\boldsymbol{\gamma},\boldsymbol{\eta}_{\mathbf{u}},\boldsymbol{\tau},\mathbf{v},\rho)f(\boldsymbol{\beta}|\boldsymbol{\gamma})f(\boldsymbol{\eta}_{\mathbf{u}}|\boldsymbol{\tau})f(\mathbf{v}|\boldsymbol{\gamma})f(\boldsymbol{\tau}|\boldsymbol{\gamma})f(\boldsymbol{\gamma}|\pi_{\beta})f(\pi_{\beta})f(\pi_1)f(\pi_0)f(\pi_c)$$
$$\propto f(\mathbf{x}|\boldsymbol{\beta})f(\mathbf{y}|\alpha,\boldsymbol{\beta},\boldsymbol{\eta}_{\mathbf{u}},\mathbf{v},\rho)f(\boldsymbol{\beta}|\boldsymbol{\gamma})f(\boldsymbol{\eta}_{\mathbf{u}}|\boldsymbol{\tau})f(\mathbf{v}|\boldsymbol{\gamma})f(\boldsymbol{\tau}|\boldsymbol{\gamma})f(\boldsymbol{\gamma}|\pi_{\beta})f(\pi_{\beta})f(\pi_1)f(\pi_0)f(\pi_c) \propto$$
$$(2\pi)^{-\frac{n_2}{2}}(\sigma_y^2)^{-\frac{n_2}{2}}\exp\left(-\frac{\left(\mathbf{y}-\alpha\boldsymbol{Z}_{y,-j}\boldsymbol{\beta}_{-j}-\rho\boldsymbol{Z}_{y,-j}(\boldsymbol{\beta}_{-j}\circ\mathbf{v}_{-j})-\alpha\mathbf{Z}_{y,j}\beta_j-\rho\mathbf{Z}_{y,j}\beta_j v_j-\mathbf{Z}_{y,j}\eta_{uj}-\mathbf{Z}_{y,-j}\boldsymbol{\eta}_{u,-j}\right)^T\left(\mathbf{y}-\alpha\boldsymbol{Z}_{y,-j}\boldsymbol{\beta}_{-j}-\rho\boldsymbol{Z}_{y,-j}(\boldsymbol{\beta}_{-j}\circ\mathbf{v}_{-j})-\alpha\mathbf{Z}_{y,j}\beta_j-\rho\mathbf{Z}_{y,j}\beta_j v_j-\mathbf{Z}_{y,j}\eta_{uj}-\mathbf{Z}_{y,-j}\boldsymbol{\eta}_{u,-j}\right)}{2\sigma_y^2}\right)$$
$$(2\pi)^{-\frac{n_1}{2}}(\sigma_x^2)^{-\frac{n_1}{2}}\exp\left(-\frac{\left(\mathbf{x}-\boldsymbol{Z}_{x,-j}\boldsymbol{\beta}_{-j}-\mathbf{Z}_{x,j}\beta_j\right)^T\left(\mathbf{x}-\boldsymbol{Z}_{x,-j}\boldsymbol{\beta}_{-j}-\mathbf{Z}_{x,j}\beta_j\right)}{2\sigma_x^2}\right).$$
$$\prod_{j=1}^{p}\left[(2\pi)^{-\frac{1}{2}}(\sigma_\beta^2)^{-\frac{1}{2}}\exp\left(-\frac{\beta_j^2}{2\sigma_\beta^2}\right)\right]^{\gamma_j}\prod_{j=1}^{p}\left[\delta_0(\beta_j)\right]^{1-\gamma_j}\prod_{j=1}^{p}\left[(2\pi)^{-\frac{1}{2}}(\sigma_\eta^2)^{-\frac{1}{2}}\exp\left(-\frac{\eta_{uj}^2}{2\sigma_\eta^2}\right)\right]^{\tau_j}$$
$$\prod_{j=1}^{p}\left[\delta_0(\eta_{uj})\right]^{1-\tau_j}\pi_c^{\sum_{j=1}^{p}\gamma_j v_j}(1-\pi_c)^{\sum_{j=1}^{p}\gamma_j(1-v_j)}$$
$$\pi_\beta^{\sum_{j=1}^{p}\gamma_j}(1-\pi_\beta)^{\sum_{j=1}^{p}(1-\gamma_j)}\pi_1^{\sum_{j=1}^{p}\gamma_j\tau_j}(1-\pi_1)^{\sum_{j=1}^{p}\gamma_j(1-\tau_j)}\pi_0^{\sum_{j=1}^{p}(1-\gamma_j)\tau_j}(1-\pi_0)^{\sum_{j=1}^{p}(1-\gamma_j)(1-\tau_j)}$$
$$\cdot\, beta(\pi_\beta,\lambda_{\beta1},\lambda_{\beta2})\, beta(\pi_1,\lambda_{21},\lambda_{22}) beta(\pi_0,\lambda_{31},\lambda_{32})\, beta(\pi_c,\lambda_{c1},\lambda_{c2})$$
$$\propto C \cdot \exp\left(-\frac{\left(\mathbf{y}-\alpha\boldsymbol{Z}_{y,-j}\boldsymbol{\beta}_{-j}-\rho\boldsymbol{Z}_{y,-j}(\boldsymbol{\beta}_{-j}\circ\mathbf{v}_{-j})-\mathbf{Z}_y\boldsymbol{\eta}_u\right)^T\left(\mathbf{y}-\alpha\boldsymbol{Z}_{y,-j}\boldsymbol{\beta}_{-j}-\rho\boldsymbol{Z}_{y,-j}(\boldsymbol{\beta}_{-j}\circ\mathbf{v}_{-j})-\mathbf{Z}_y\boldsymbol{\eta}_u\right)}{2\sigma_y^2}\right)$$
$$\exp\left(-\frac{\left(\mathbf{x}-\boldsymbol{Z}_{x,-j}\boldsymbol{\beta}_{-j}\right)^T\left(\mathbf{x}-\boldsymbol{Z}_{x,-j}\boldsymbol{\beta}_{-j}\right)}{2\sigma_x^2}\right).$$
$$\prod_{i\neq j}^{p}\left[\pi_\beta(2\pi)^{-\frac{1}{2}}(\sigma_\beta^2)^{-\frac{1}{2}}\exp\left(-\frac{\beta_i^2}{2\sigma_\beta^2}\right)\right]^{\gamma_i}\prod_{i\neq j}^{p}\left[(1-\pi_\beta)\delta_0(\beta_i)\right]^{1-\gamma_i}$$
$$\prod_{j=1}^{p}\left[(2\pi)^{-\frac{1}{2}}(\sigma_\eta^2)^{-\frac{1}{2}}exp\left(-\frac{\eta_{uj}^2}{2\sigma_\eta^2}\right)\right]^{\tau_j}$$
$$\prod_{j=1}^{p}\left[\delta_0(\eta_{uj})\right]^{1-\tau_j}\pi_c^{\sum_{j=1}^{p}\gamma_j v_j}(1-\pi_c)^{\sum_{j=1}^{p}\gamma_j(1-v_j)}$$
$$\pi_1^{\sum_{j=1}^{p}\gamma_j\tau_j}(1-\pi_1)^{\sum_{j=1}^{p}\gamma_j(1-\tau_j)}\pi_0^{\sum_{j=1}^{p}(1-\gamma_j)\tau_j}(1-\pi_0)^{\sum_{j=1}^{p}(1-\gamma_j)(1-\tau_j)} beta(\pi_\beta,\lambda_{\beta1},\lambda_{\beta2})\, beta(\pi_1,\lambda_{21},\lambda_{22}) beta(\pi_0,\lambda_{31},\lambda_{32})\, beta(\pi_c,\lambda_{c1},\lambda_{c2})$$
$$\cdot\, exp\left(-\frac{\beta_j^2 K-2\left(\frac{(\alpha+\rho v_j)\left(\boldsymbol{y}-\alpha\boldsymbol{Z}_{y,-j}\boldsymbol{\beta}_{-j}-\rho\boldsymbol{Z}_{y,-j}(\boldsymbol{\beta}_{-j}\circ\mathbf{v}_{-j})-\mathbf{Z}_y\boldsymbol{\eta}_u\right)^T\mathbf{Z}_{y,j}}{\sigma_y^2}+\frac{\left(\mathbf{x}-\boldsymbol{Z}_{x,-j}\boldsymbol{\beta}_{-j}\right)^T\mathbf{Z}_{x,j}}{\sigma_x^2}\right)\beta_j}{2}\right)$$

where

$$C=(2\pi)^{-\frac{n_2}{2}}(\sigma_y^2)^{-\frac{n_2}{2}}(2\pi)^{-\frac{n_1}{2}}(\sigma_x^2)^{-\frac{n_1}{2}}(2\pi)^{-\frac{\gamma_j}{2}}\pi_\beta^{\gamma_j}(1-\pi_\beta)^{1-\gamma_j}(\sigma_\beta^2)^{-\frac{\gamma_j}{2}}\left(\delta_0(\beta_j)\right)^{1-\gamma_j};$$

$K$ is a scalar in the form of

$$K=\frac{(\alpha+\rho v_j)^2\mathbf{Z}_{\mathbf{y,j}}^{\mathbf{T}}\mathbf{Z}_{\mathbf{y,j}}}{\sigma_y^2}+\frac{\mathbf{Z}_{\mathbf{x,j}}^{\mathbf{T}}\mathbf{Z}_{\mathbf{x,j}}}{\sigma_x^2}+\frac{\gamma_j}{\sigma_\beta^2};$$

$\boldsymbol{\beta}_{-\mathbf{j}}$ is the vector $\boldsymbol{\beta}$ without the *j*th element $\beta_j$; $Z_{x,-j}$ is the $n_1\times(p-1)$ matrix $Z_x$ without the *j*th column vector $Z_{x,j}$; $Z_{y,-j}$ is the $n_2\times(p-1)$ matrix $Z_y$ without the *j*th column vector $Z_{y,j}$. Note that because $Z_x, Z_y$ are standardized to have mean zero and unite standard deviation, we have $Z_{y_j}^T Z_{y_j}=n_2-1, Z_{x_j}^T Z_{x_j}=n_1-1$ for any $j=1,\cdots,p$.

Based on the above likelihood, we conduct Gibbs sampling on all parameters that include $\theta=(\beta_j,\gamma_j,\eta_{uj},\tau_j,\alpha,v_j,\sigma_\beta^2,\sigma_\eta^2,\sigma_x^2,\sigma_y^2,\pi_\beta,\pi_1,\pi_0,\pi_c,\rho)$.

Specifically, given $\gamma_j$=1 and other parameters, the posterior conditional distribution of $\beta_j$ given all other parameters is a normal distribution with mean $U_{\beta_j}$ and variance $\sigma_{\beta_j}^2$, where

$$U_{\beta_j}=\left(\frac{(\alpha+\rho v_j)\left(\mathbf{y}-\alpha Z_{y,-j}\boldsymbol{\beta}_{-\mathbf{j}}-\rho Z_{y,-j}(\boldsymbol{\beta}_{-\mathbf{j}}\circ\mathbf{v}_{-\mathbf{j}})-\mathbf{Z}_{\mathbf{y}}\boldsymbol{\eta}_{\mathbf{u}}\right)^T\mathbf{Z}_{\mathbf{y,j}}}{\sigma_y^2}+\frac{\left(\mathbf{x}-Z_{x,-j}\boldsymbol{\beta}_{-\mathbf{j}}\right)^T\mathbf{Z}_{\mathbf{x,j}}}{\sigma_x^2}\right)/K_1$$

$$\sigma_{\beta_j}^2=1/K_1,\ K_1=\frac{(\alpha+\rho v_j)^2\mathbf{Z}_{\mathbf{y,j}}^{\mathbf{T}}\mathbf{Z}_{\mathbf{y,j}}}{\sigma_y^2}+\frac{\mathbf{Z}_{\mathbf{x,j}}^{\mathbf{T}}\mathbf{Z}_{\mathbf{x,j}}}{\sigma_x^2}+\frac{1}{\sigma_\beta^2}$$

Given $\tau_j$=1 and other parameters, the posterior conditional distribution of $\eta_{uj}$ is a normal distribution with mean $U_{\eta_j}$ and variance $\sigma_{\eta_j}^2$, where

$$U_{\eta_j}=\left(\frac{\left(\mathbf{y}-Z_{y,-j}\boldsymbol{\eta}_{\mathbf{u},-\mathbf{j}}-\alpha\mathbf{Z}_{\mathbf{y}}\boldsymbol{\beta}-\rho Z_y(\boldsymbol{\beta}\circ\mathbf{v})\right)^T\mathbf{Z}_{\mathbf{y,j}}}{\sigma_y^2}\right)/K_2$$

$$\sigma_{\eta_j}^2=1/K_2,\ K_2=\frac{\mathbf{Z}_{\mathbf{y,j}}^{\mathbf{T}}\mathbf{Z}_{\mathbf{y,j}}}{\sigma_y^2}+\frac{1}{\sigma_\eta^2}$$

After integrating out the parameter $\beta_j$, we obtain the posterior conditional distribution for $\gamma_j$ as

$$p\left(\gamma_j=1|\boldsymbol{\beta}_{-\mathbf{j}},\boldsymbol{\eta}_{\mathbf{u}},\alpha,\sigma_x^2,\sigma_y^2,\sigma_\beta^2,\pi_\beta,\pi_1,\pi_0,\pi_c,\rho\right)\propto\exp\left(\frac{\mu_{\beta_j}^2}{2\sigma_{\beta_j}^2}+0.5*\log\left(\sigma_{\beta_j}^2\right)-0.5*\log\left(\sigma_\beta^2\right)+\log(\pi_\beta)+\tau_j\log(\pi_1)+\left(1-\tau_j\right)\log(1-\pi_1)+v_j\log(\pi_c)+\left(1-v_j\right)\log(1-\pi_c)\right),$$

$$p\left(\gamma_j=0|\boldsymbol{\beta}_{-\mathbf{j}},\boldsymbol{\eta}_{\mathbf{u}},\alpha,\sigma_x^2,\sigma_y^2,\sigma_\beta^2,\pi_\beta,\pi_1,\pi_0\right)\propto\exp\left(\log(1-\pi_\beta)+\tau_j\log(\pi_0)+\left(1-\tau_j\right)\log(1-\pi_0)\right).$$

After integrating out the parameter $\eta_j$, we obtain the posterior conditional distribution for $\tau_j$ as

$$p\left(\tau_j=1|\eta_{u,-j},\beta,\alpha,\sigma_y^2,\sigma_\eta^2,\pi_1,\pi_0\right)\propto\exp\left(\frac{\mu_{\eta_j}^2}{2\sigma_{\eta_j}^2}+0.5*\log\left(\sigma_{\eta_j}^2\right)-0.5*\log\left(\sigma_\eta^2\right)+\gamma_j\log(\pi_1)+(1-\gamma_j)\log(\pi_0)\right)$$

$$p\left(\tau_j=0|\eta_{u,-j},\beta,\alpha,\sigma_y^2,\sigma_\eta^2,\pi_1,\pi_0\right)\propto\exp\left(\gamma_j\log(1-\pi_1)+(1-\gamma_j)\log(1-\pi_0)\right)$$

We also obtain the posterior conditional distribution for $v_j$ as

$$p\left(v_j=1|\eta_u,\beta,\alpha,\sigma_y^2,\sigma_\eta^2,\pi_c,\pi_1,\pi_0\right)$$
$$\propto\exp\left(-\frac{\rho^2\beta_j^2\mathbf{Z}_{\mathbf{y,j}}^{\mathbf{T}}\mathbf{Z}_{\mathbf{y,j}}-2\left(\mathbf{y}-\alpha\boldsymbol{Z}_{\boldsymbol{y}}\boldsymbol{\beta}-\rho\boldsymbol{Z}_{y,-j}(\boldsymbol{\beta}_{-\mathbf{j}}\circ\mathbf{v}_{-\mathbf{j}})-\mathbf{Z}_{\mathbf{y}}\boldsymbol{\eta}_{\mathbf{u}}\right)^T\rho\mathbf{Z}_{\mathbf{y,j}}\beta_j}{2\sigma_y^2}+\gamma_j\log(\pi_c)\right)$$

$$p\left(v_j=0|\boldsymbol{\eta}_{\mathbf{u}},\beta,\alpha,\sigma_y^2,\sigma_\eta^2,\pi_c,\pi_1,\pi_0\right)\propto\exp\left(\gamma_j\log(1-\pi_c)\right)$$

The posterior conditional distribution of $\rho$ is a normal distribution with variance $\sigma_\rho^2=$

$\left(\frac{(\mathbf{v}\circ\boldsymbol{\beta})^T Z_y^T Z_y(\mathbf{v}\circ\boldsymbol{\beta})}{\sigma_y^2}\right)^{-1}$ and mean $\mu_\rho = \frac{(\mathbf{v}\circ\boldsymbol{\beta})^T Z_y^T\mathbf{y} - (\mathbf{v}\circ\boldsymbol{\beta})^T Z_y^T Z_y \boldsymbol{\eta}_\mathbf{u} - (\mathbf{v}\circ\boldsymbol{\beta})^T Z_y^T Z_y \boldsymbol{\beta}\alpha}{\sigma_y^2}\sigma_\rho^2$.

The posterior conditional distribution of $\alpha$ is a normal distribution with variance $\sigma_\alpha^2 = \left(\frac{\boldsymbol{\beta}^\mathrm{T} Z_y^T Z_y \boldsymbol{\beta}}{\sigma_y^2}\right)^{-1}$ and mean $\mu_\alpha = \frac{\boldsymbol{\beta}^\mathrm{T} Z_y^T \mathbf{y} - \boldsymbol{\beta}^T Z_y^T Z_y \boldsymbol{\eta}_\mathbf{u} - \boldsymbol{\beta}^\mathrm{T} Z_y^T Z_y(\boldsymbol{\beta}\circ\mathbf{v})\rho}{\sigma_y^2}\sigma_\alpha^2$.

The posterior conditional distribution of $\pi_\beta$ is $beta(\sum_{j=1}^p \gamma_j + \lambda_{\beta 1}, \sum_{j=1}^p (1-\gamma_j) + \lambda_{\beta 2})$.

The posterior conditional distribution of $\pi_1$ is $beta(\sum_{j=1}^p \gamma_j \tau_j + \lambda_{21}, \sum_{j=1}^p \gamma_j (1-\tau_j) + \lambda_{22})$.

The posterior conditional distribution of $\pi_0$ is $beta(\sum_{j=1}^p (1-\gamma_j)\tau_j + \lambda_{31}, \sum_{j=1}^p (1-\gamma_j)(1-\tau_j) + \lambda_{32})$.

The posterior conditional distribution of $\pi_c$ is $beta(\sum_{j=1}^p \gamma_j v_j + \lambda_{\mathrm{c}1}, \sum_{j=1}^p \gamma_j (1-v_j) + \lambda_{\mathrm{c}2})$.

The posterior conditional distribution of $\sigma_x^2$ is an inverse gamma distribution with the shape parameter $\sigma_{x_{shape}}^2 = \frac{n_1}{2} - 1$ and the scale parameter$\sigma_{x_scale}^2 = (\mathbf{x}^T\mathbf{x} + \boldsymbol{\beta}^\mathbf{T} Z_x^T Z_x \boldsymbol{\beta} - 2\boldsymbol{\beta}^T Z_x^T \mathbf{x})/2$.

The posterior conditional distribution of $\sigma_y^2$ is an inverse gamma distribution with the shape parameter $\sigma_{y\_shape}^2 = \frac{n_2}{2} - 1$ and the scale parameter.

$$\sigma_{y\_scale}^2 = \left(\mathbf{y} - \alpha \mathrm{Z_y}\boldsymbol{\beta} - \rho \mathrm{Z_y}(\boldsymbol{\beta}\circ\mathbf{v}) - \mathrm{Z_y}\boldsymbol{\eta}_\mathbf{u}\right)^T (\mathbf{y} - \alpha \mathrm{Z_y}\boldsymbol{\beta} - \rho \mathrm{Z_y}(\boldsymbol{\beta}\circ\mathbf{v}) - \mathrm{Z_y}\boldsymbol{\eta}_\mathbf{u})/2$$

The posterior conditional distribution of $\sigma_\beta^2$ is an inverse gamma distribution with the shape parameter $\sigma_{\beta\_shape}^2 = \frac{\sum_{j=1}^p \gamma_j}{2} + a_\beta$, and the scale parameter $\sigma_{\beta\_scale}^2 = \frac{\sum_{j=1}^p \gamma_j \beta_j^2 + 2b_\beta}{2}$.

The posterior conditional distribution of $\sigma_\eta^2$ is an inverse gamma distribution with the shape parameter $\sigma_{\eta\_shape}^2 = \frac{\sum_{j=1}^p \tau_j}{2} + a_\eta$, and the scale parameter $\sigma_{\eta\_scale}^2 = \frac{\sum_{j=1}^p \tau_j \eta_{uj}^2 + 2b_\eta}{2}$.

For all analyses in the present study, we ran 1,000 Gibbs sampling iterations with the first 200 as burn-in and obtain the posterior samples of $\alpha$ for hypothesis testing, which is described in the next section.

**3. Parameter estimation and $p$ value computation**

We denote $\mu_\alpha$ and $\sigma_\alpha^2$ as the posterior mean and the posterior variance of the causal effect parameter $\alpha$. Since both the likelihood and the posterior follow normal distributions asymptotically, and because we also use a normal distribution $N(0,\sigma_0^2)$ as the prior distribution, we can easily obtain the approximate maximum likelihood estimate and its standard error by the method of moments as

$$\hat{\alpha} = \frac{\sigma_0^2 \mu_\alpha}{\sigma_0^2 - \sigma_\alpha^2},$$
$$se(\hat{\alpha}) = \frac{\sigma_0 \sigma_\alpha}{\sqrt{\sigma_0^2 - \sigma_\alpha^2}}.$$

The resulting z-score $Z = \frac{\hat{\alpha}}{se(\hat{\alpha})}$ follows standard normal distribution asymptotically, which allows us to obtain a corresponding $p$ value for testing the null hypothesis $H_0: \alpha = 0$. In present study, because we use a non-informative prior for $\alpha$ by set $\sigma_0^2 \to \infty$, we have $\hat{\alpha} = \mu_\alpha$ and $se(\hat{\alpha}) = \sigma_\alpha$. Note that the choice of the prior on $\alpha$ does not influence the inference results as we only use the likelihood, expressed as the ratio of the posterior and prior, for hypothesis test[45,46].

#### 4. MRAID model for summary statistics

The MRAID model for summary statistics can be presented in the form of marginal effect size estimates and the LD matrix. We denote the LD structure of the instrumental SNPs as $\Sigma_1$ in exposure GWAS data, and $\Sigma_2$ in outcome GWAS data, both are $p \times p$ symmetric positive definite matrices. Often, $\Sigma_1$ and $\Sigma_2$ are from the same LD reference panels (e.g. the individuals with European ancestry from the 1,000 Genomes project). The proposed MRAID model for summary statistics can be constructed as the following two equations

$$\widehat{\boldsymbol{\beta}}_{\mathrm{x}} = \Sigma_1 \boldsymbol{\beta} + \mathbf{e}_{\mathrm{x}} \qquad (3)$$

$$\widehat{\boldsymbol{\beta}}_{\mathrm{y}} = \alpha \Sigma_2 \boldsymbol{\beta} + \rho \Sigma_2 (\boldsymbol{\beta} \circ \mathbf{v}) + \Sigma_2 \boldsymbol{\eta}_{\mathbf{u}} + \mathbf{e}_{\mathrm{y}} \qquad (4)$$

where $\widehat{\boldsymbol{\beta}}_{\mathbf{x}}$ is the p-vector of estimates for the marginal SNP effect sizes on the exposure; $\widehat{\boldsymbol{\beta}}_{\mathbf{y}}$ is the $p$-vector of estimates for the marginal SNP effect sizes on the outcome; both $\mathbf{e}_{\mathrm{x}}$ and $\mathbf{e}_{\mathrm{y}}$ are $p$-vector of residual errors. $\mathbf{e}_{\mathrm{x}}$ is a $p$-vector of residual error following a multivariate normal distribution $\mathrm{N}(0, \Sigma_1 \sigma_{\mathrm{x}}^2/(n_1 - 1))$, and $\mathbf{e}_{\mathrm{y}}$ is a p-vector of residual error following a multivariate normal distribution $\mathrm{N}\left(0, \Sigma_2 \sigma_{\mathrm{y}}^2/(n_2 - 1)\right)$.

#### 5. Detailed sampling steps of MRAID model for summary statistics

For the summary statistics version of MRAID, we can obtain the posterior distribution for all parameters as

$$f\left(\alpha, \boldsymbol{\beta}, \boldsymbol{\gamma}, \boldsymbol{\eta}_{\mathbf{u}}, \boldsymbol{\tau}, \mathbf{v}, \rho | \widehat{\boldsymbol{\beta}}_{\mathrm{x}}, \widehat{\boldsymbol{\beta}}_{\mathrm{y}}\right)$$
$$\propto f\left(\widehat{\boldsymbol{\beta}}_{\mathrm{x}}, \widehat{\boldsymbol{\beta}}_{\mathrm{y}} | \alpha, \boldsymbol{\beta}, \boldsymbol{\gamma}, \boldsymbol{\eta}_{\mathbf{u}}, \boldsymbol{\tau}, \mathbf{v}, \rho\right) f(\boldsymbol{\beta}|\boldsymbol{\gamma}) f(\boldsymbol{\eta}_{\mathbf{u}}|\boldsymbol{\tau}) f(\mathbf{v}|\boldsymbol{\gamma}) f\left(\boldsymbol{\gamma}|\pi_\beta\right) f(\boldsymbol{\tau}|\boldsymbol{\gamma}) f(\pi_\beta) f(\pi_1) f(\pi_0) f(\pi_c)$$
$$\propto f\left(\widehat{\boldsymbol{\beta}}_{\mathbf{x}}|\boldsymbol{\beta}\right) f\left(\widehat{\boldsymbol{\beta}}_{\mathrm{y}} | \alpha, \boldsymbol{\beta}, \boldsymbol{\eta}_{\mathbf{u}}, \mathbf{v}, \rho\right) f(\boldsymbol{\beta}|\boldsymbol{\gamma}) f(\boldsymbol{\eta}_{\mathbf{u}}|\boldsymbol{\tau}) f(\mathbf{v}|\boldsymbol{\gamma}) f\left(\boldsymbol{\gamma}|\pi_\beta\right) f(\boldsymbol{\tau}|\boldsymbol{\gamma}) f(\pi_\beta) f(\pi_1) f(\pi_0) f(\pi_c)$$
$$\propto (2\pi)^{-\frac{p}{2}} \left(\frac{\sigma_{\mathrm{y}}^2}{n_2 - 1}\right)^{-\frac{p}{2}} |\Sigma_2|^{-\frac{1}{2}}$$

$$\exp\left( - \frac{\begin{array}{c}\left(\widehat{\boldsymbol{\beta}}_{\mathrm{y}} - \alpha \Sigma_{2,-j} \boldsymbol{\beta}_{-\mathbf{j}} - \alpha \Sigma_{2,j} \beta_j - \rho \Sigma_{2,-j}\left(\boldsymbol{\beta}_{-\mathbf{j}} \circ \mathbf{v}_{-\mathbf{j}}\right) - \rho \Sigma_{2,j}(\beta_j \circ v_j) - \Sigma_2 \boldsymbol{\eta}_{\mathbf{u}}\right)^T (n_2 - 1) \Sigma_2^{-1} \\ \left(\widehat{\boldsymbol{\beta}}_{\mathrm{y}} - \alpha \Sigma_{2,-j} \boldsymbol{\beta}_{-\mathbf{j}} - \alpha \Sigma_{2,j} \beta_j - \rho \Sigma_{2,-j}\left(\boldsymbol{\beta}_{-\mathbf{j}} \circ \mathbf{v}_{-\mathbf{j}}\right) - \rho \Sigma_{2,j}(\beta_j \circ v_j) - \Sigma_2 \boldsymbol{\eta}_{\mathbf{u}}\right)\end{array}}{2\sigma_{\mathrm{y}}^2} \right)$$

$$(2\pi)^{-\frac{p}{2}} \left(\frac{\sigma_{\mathrm{x}}^2}{n_1 - 1}\right)^{-\frac{p}{2}} |\Sigma_1|^{-\frac{1}{2}} \exp\left( - \frac{\left(\widehat{\boldsymbol{\beta}}_{\mathrm{x}} - \Sigma_{1,-j} \boldsymbol{\beta}_{-\mathbf{j}} - \Sigma_{1,j} \beta_j\right)^T (\mathrm{n}_1 - 1) \Sigma_1^{-1} \left(\widehat{\boldsymbol{\beta}}_{\mathrm{x}} - \Sigma_{1,-j} \boldsymbol{\beta}_{-\mathbf{j}} - \Sigma_{1,j} \beta_j\right)}{2\sigma_{\mathrm{x}}^2} \right)$$

$$\prod_{j=1}^{p} \left[ (2\pi)^{-\frac{1}{2}} \left(\sigma_\beta^2\right)^{-\frac{1}{2}} \exp\left(-\frac{\beta_j^2}{2\sigma_\beta^2}\right) \right]^{\gamma_j} \prod_{j=1}^{p} \left[\delta_0(\beta_j)\right]^{1-\gamma_j} \prod_{j=1}^{p} \left[ (2\pi)^{-\frac{1}{2}} \left(\sigma_\eta^2\right)^{-\frac{1}{2}} exp\left(-\frac{\eta_{uj}^2}{2\sigma_\eta^2}\right) \right]^{\tau_j}$$

$$\prod_{j=1}^{p} \left[\delta_0(\eta_{uj})\right]^{1-\tau_j} \pi_c^{\sum_{j=1}^{p} \gamma_j v_j} (1 - \pi_c)^{\sum_{j=1}^{p} \gamma_j (1 - v_j)}$$

$$\pi_\beta^{\sum_{j=1}^{p} \gamma_j} \left(1 - \pi_\beta\right)^{\sum_{j=1}^{p} (1 - \gamma_j)} \pi_1^{\sum_{j=1}^{p} \gamma_j \tau_j} (1 - \pi_1)^{\sum_{j=1}^{p} \gamma_j (1 - \tau_j)} \pi_0^{\sum_{j=1}^{p} (1 - \gamma_j) \tau_j} (1 - \pi_0)^{\sum_{j=1}^{p} (1 - \gamma_j)(1 - \tau_j)}$$

$$beta(\pi_\beta, \lambda_{\beta 1}, \lambda_{\beta 2})\ beta(\pi_1, \lambda_{21}, \lambda_{22}) beta(\pi_0, \lambda_{31}, \lambda_{32})\ beta(\pi_c, \lambda_{c1}, \lambda_{c2})$$

$$\propto C\cdot exp\left(-\frac{\left(\widehat{\boldsymbol{\beta}}_y-\alpha\Sigma_{2,-j}\boldsymbol{\beta}_{-j}-\rho\Sigma_{2,-j}(\boldsymbol{\beta}_{-j}\circ\mathbf{v}_{-j})-\Sigma_2\boldsymbol{\eta}_{\mathbf{u}}\right)^T(n_2-1)\Sigma_2^{-1}\left(\widehat{\boldsymbol{\beta}}_y-\alpha\Sigma_{2,-j}\boldsymbol{\beta}_{-j}-\rho\Sigma_{2,-j}(\boldsymbol{\beta}_{-j}\circ\mathbf{v}_{-j})-\Sigma_2\boldsymbol{\eta}_{\mathbf{u}}\right)}{2\sigma_y^2}\right)$$

$$\exp\left(-\frac{\left(\widehat{\boldsymbol{\beta}}_x-\Sigma_{1,-j}\boldsymbol{\beta}_{-j}\right)^T(n_1-1)\Sigma_1^{-1}\left(\widehat{\boldsymbol{\beta}}_x-\Sigma_{1,-j}\boldsymbol{\beta}_{-j}\right)}{2\sigma_x^2}\right)$$

$$\prod_{i\neq j}^{p}\left[\pi_\beta(2\pi)^{-\frac{1}{2}}(\sigma_\beta^2)^{-\frac{1}{2}}exp\left(-\frac{\beta_i^2}{2\sigma_\beta^2}\right)\right]^{\gamma_i}\prod_{i\neq j}^{p}\left[(1-\pi_\beta)\delta_0(\beta_i)\right]^{1-\gamma_i}$$

$$\prod_{j=1}^{p}\left[(2\pi)^{-\frac{1}{2}}(\sigma_\eta^2)^{-\frac{1}{2}}exp\left(-\frac{\eta_{uj}^2}{2\sigma_\eta^2}\right)\right]^{\tau_j}$$

$$\prod_{j=1}^{p}\left[\delta_0(\eta_{uj})\right]^{1-\tau_j}\pi_c^{\sum_{j=1}^{p}\gamma_j v_j}(1-\pi_c)^{\sum_{j=1}^{p}\gamma_j(1-v_j)}$$

$$\pi_1^{\sum_{j=1}^{p}\gamma_j\tau_j}(1-\pi_1)^{\sum_{j=1}^{p}\gamma_j(1-\tau_j)}\pi_0^{\sum_{j=1}^{p}(1-\gamma_j)\tau_j}(1-\pi_0)^{\sum_{j=1}^{p}(1-\gamma_j)(1-\tau_j)}$$

$$beta(\pi_\beta,\lambda_{\beta1},\lambda_{\beta2})\;beta(\pi_1,\lambda_{21},\lambda_{22})beta(\pi_0,\lambda_{31},\lambda_{32})\;beta(\pi_c,\lambda_{c1},\lambda_{c2})$$

$$\cdot\,exp\left(-\frac{\beta_j^2K-2\left(\frac{(\alpha+\rho v_j)\left(\widehat{\boldsymbol{\beta}}_y-\alpha\Sigma_{2,-j}\boldsymbol{\beta}_{-j}-\rho\Sigma_{2,-j}(\boldsymbol{\beta}_{-j}\circ\mathbf{v}_{-j})-\Sigma_2\boldsymbol{\eta}_{\mathbf{u}}\right)^T(n_2-1)\Sigma_2^{-1}\Sigma_{2,j}}{\sigma_y^2}+\frac{\left(\widehat{\boldsymbol{\beta}}_x-\Sigma_{1,-j}\boldsymbol{\beta}_{-j}\right)^T(n_1-1)\Sigma_1^{-1}\Sigma_{1,j}}{\sigma_x^2}\right)\beta_j}{2}\right)$$

where

$$C=(2\pi)^{-\frac{p}{2}}\left(\frac{\sigma_y^2}{n_2-1}\right)^{-\frac{p}{2}}|\Sigma_2|^{-\frac{1}{2}}(2\pi)^{-\frac{p}{2}}\left(\frac{\sigma_x^2}{n_1-1}\right)^{-\frac{p}{2}}|\Sigma_1|^{-\frac{1}{2}}(2\pi)^{-\frac{\gamma_j}{2}}\pi_\beta^{\gamma_j}(1-\pi_\beta)^{1-\gamma_j}(\sigma_\beta^2)^{-\frac{\gamma_j}{2}}\left(\delta_0(\beta_j)\right)^{1-\gamma_j};$$

$K$ is a scalar in the form of

$$K=\frac{(\alpha+\rho\boldsymbol{v_j})^2(n_2-1)}{\sigma_y^2}+\frac{(n_1-1)}{\sigma_x^2}+\frac{\gamma_j}{\sigma_\beta^2}$$

$\boldsymbol{\beta}_{-j}$ is the vector $\boldsymbol{\beta}$ without the $j$th element $\beta_j$.;

Based on the above likelihood, we conduct Gibbs sampling on all parameters that include $\theta=(\beta_j,\gamma_j,\eta_{uj},\tau_j,v_j,\alpha,\sigma_\beta^2,\sigma_\eta^2,\sigma_x^2,\sigma_y^2,\pi_\beta,\pi_1,\pi_0,\pi_c,\rho)$.

Given $\gamma_j$=1 and other parameters, the posterior distribution of $\beta_j$ is a normal distribution with mean $U_{\beta_j}$ and variance $\sigma_{\beta_j}^2$, where

$$U_{\beta_j}=\left(\frac{(\alpha+\rho v_j)\left(\widehat{\boldsymbol{\beta}}_y-\alpha\Sigma_{2,-j}\boldsymbol{\beta}_{-j}-\rho\Sigma_{2,-j}(\boldsymbol{\beta}_{-j}\circ\mathbf{v}_{-j})-\Sigma_2\boldsymbol{\eta}_{\mathbf{u}}\right)^T(n_2-1)\Sigma_2^{-1}\Sigma_{2,j}}{\sigma_y^2}+\frac{\left(\widehat{\boldsymbol{\beta}}_x-\Sigma_{1,-j}\boldsymbol{\beta}_{-j}\right)^T(n_1-1)\Sigma_1^{-1}\Sigma_{1,j}}{\sigma_x^2}\right)/K_1$$

$$\sigma_{\beta_j}^2=1/K_1,\;K_1=\frac{(\alpha+\rho v_j)^2(n_2-1)}{\sigma_y^2}+\frac{(n_1-1)}{\sigma_x^2}+\frac{1}{\sigma_\beta^2}$$

Given $\tau_j$=1 and other parameters, the posterior conditional distribution of $\eta_{uj}$ is a normal distribution with mean $U_{\eta_j}$ and variance $\sigma_{\eta_j}^2$, where

$$U_{\eta_j}=\left(\frac{\left(\widehat{\boldsymbol{\beta}}_y-\Sigma_{2,-j}\boldsymbol{\eta}_{\mathbf{u},-j}-\alpha\Sigma_2\boldsymbol{\beta}-\rho\Sigma_2(\boldsymbol{\beta}\circ\mathbf{v})\right)^T(n_2-1)\Sigma_2^{-1}\Sigma_{2,j}}{\sigma_y^2}\right)/K_2$$

$$\sigma_{\eta_j}^2=1/K_2,\;K_2=\frac{n_2-1}{\sigma_y^2}+\frac{1}{\sigma_\eta^2}$$

After integrating out the parameter $\beta_j$, we obtain the posterior conditional distribution of $\gamma_j$

as

$$p(\gamma_j = 1|\beta_{-j}, \eta_u, \alpha, \sigma_x^2, \sigma_y^2, \sigma_\beta^2, \pi_\beta, \pi_1, \pi_0, \pi_c, \rho) \propto \exp\left(\frac{\mu_{\beta_j}^2}{2\sigma_{\beta_j}^2} + 0.5 * \log\left(\sigma_{\beta_j}^2\right) - 0.5 * \log(\sigma_\beta^2) + \log(\pi_\beta) + \tau_j \log(\pi_1) + (1-\tau_j)\log(1-\pi_1) + v_j \log(\pi_c) + (1-v_j)\log(1-\pi_c)\right)$$

$$p(\gamma_j = 0|\beta_{-j}, \eta_u, \alpha, \sigma_x^2, \sigma_y^2, \sigma_\beta^2, \pi_\beta, \pi_1, \pi_0) \propto \exp(\log(1-\pi_\beta) + \tau_j \log(\pi_0) + (1-\tau_j)\log(1-\pi_0))$$

After integrating out the parameter $\eta_{uj}$, we obtain the posterior conditional distribution of $\tau_j$ as

$$p(\tau_j = 1|\eta_{u,-j}, \beta, \alpha, \sigma_y^2, \sigma_\eta^2, \pi_1, \pi_0) \propto \exp\left(\frac{\mu_{\eta_j}^2}{2\sigma_{\eta_j}^2} + 0.5 * \log\left(\sigma_{\eta_j}^2\right) - 0.5 * \log(\sigma_\eta^2) + \gamma_j \log(\pi_1) + (1-\gamma_j)\log(\pi_0)\right)$$

$$p(\tau_j = 0|\eta_{-j}, \beta, \alpha, \sigma_y^2, \sigma_\eta^2, \pi_1, \pi_0) \propto \exp(\gamma_j \log(1-\pi_1) + (1-\gamma_j)\log(1-\pi_0))$$

We also obtain the posterior conditional distribution of $v_j$ as

$$p(v_j = 1|\eta_u, \beta, \alpha, \sigma_y^2, \sigma_\eta^2, \pi_c, \pi_1, \pi_0) \propto \exp\left(-\frac{\rho^2\beta_j^2(\mathbf{n_2}-\mathbf{1}) - 2\left(\widehat{\boldsymbol{\beta}}_{\mathbf{y}} - \alpha\Sigma_2\boldsymbol{\beta} - \rho\Sigma_{\mathbf{2},-j}(\boldsymbol{\beta}_{-\mathbf{j}} \circ \mathbf{v}_{-\mathbf{j}}) - \Sigma_2\boldsymbol{\eta}_{\mathbf{u}}\right)^T (n_2-1)\Sigma_2^{-1}\rho\Sigma_{\mathbf{2},j}\beta_j}{2\sigma_y^2} + \gamma_j \log(\pi_c)\right)$$

$$p(I_j = 0|\eta_u, \beta, \alpha, \sigma_y^2, \sigma_\eta^2, \pi_c, \pi_1, \pi_0) \propto \exp\left(\gamma_j \log(1-\pi_c)\right)$$

The posterior conditional distribution of $\rho$ is a normal distribution with variance $\sigma_\rho^2 = \left(\frac{(\mathbf{v}\circ\boldsymbol{\beta})^T(n_2-1)\Sigma_2(\mathbf{v}\circ\boldsymbol{\beta})}{\sigma_y^2}\right)^{-1}$ and mean $\mu_\rho = \frac{(\mathbf{v}\circ\boldsymbol{\beta})^T(n_2-1)\widehat{\boldsymbol{\beta}}_{\mathbf{y}} - (\mathbf{v}\circ\boldsymbol{\beta})^T(n_2-1)\Sigma_2\boldsymbol{\eta}_{\mathbf{u}} - (\mathbf{v}\circ\boldsymbol{\beta})^T(n_2-1)\Sigma_2\boldsymbol{\beta}\alpha}{\sigma_y^2}\sigma_\rho^2$.

The posterior conditional distribution of $\alpha$ is a normal distribution with variance $\sigma_\alpha^2 = \left(\frac{\boldsymbol{\beta}^T(n_2-1)\Sigma_2\boldsymbol{\beta}}{\sigma_y^2}\right)^{-1}$ and mean $\mu_\alpha = \frac{\boldsymbol{\beta}^T(n_2-1)\widehat{\boldsymbol{\beta}}_{\mathbf{y}} - \boldsymbol{\beta}^T(n_2-1)\Sigma_2\boldsymbol{\eta}_{\mathbf{u}} - \boldsymbol{\beta}^T(n_2-1)\Sigma_2(\boldsymbol{\beta}\circ\mathbf{v})\rho}{\sigma_y^2}\sigma_\alpha^2$.

The posterior conditional distribution of $\pi_\beta$ is $beta(\sum_{j=1}^p \gamma_j + \lambda_{\beta 1}, \sum_{j=1}^p (1-\gamma_j) + \lambda_{\beta 2})$.

The posterior conditional distribution of $\pi_1$ is $beta(\sum_{j=1}^p \gamma_j\tau_j + \lambda_{21}, \sum_{j=1}^p \gamma_j(1-\tau_j) + \lambda_{22})$

The posterior conditional distribution of $\pi_0$ is $beta(\sum_{j=1}^p (1-\gamma_j)\tau_j + \lambda_{31}, \sum_{j=1}^p (1-\gamma_j)(1-\tau_j) + \lambda_{32})$

The posterior conditional distribution of $\pi_c$ is $beta(\sum_{j=1}^p \gamma_j v_j + \lambda_{c1}, \sum_{j=1}^p \gamma_j(1-v_j) + \lambda_{c2})$

The posterior conditional distribution of $\sigma_x^2$ is an inverse gamma distribution with the shape parameter $\sigma_{x_{shape}}^2 = \frac{n_1}{2} - 1$ and the scale parameter

$$\sigma_{x\_scale}^2 = ((n_1-1) + \boldsymbol{\beta}^T(n_1-1)\Sigma_1\boldsymbol{\beta} - 2\boldsymbol{\beta}^T(n_1-1)\widehat{\boldsymbol{\beta}}_{\mathbf{x}})/2$$

The posterior conditional distribution of $\sigma_y^2$ is an inverse gamma distribution with the shape parameter $\sigma_{y_{shape}}^2 = \frac{n_2}{2} - 1$, and the scale parameter

$$\sigma^2_{y_{scale}} = ((n_2 - 1) + \alpha^2 \boldsymbol{\beta}^T (n_2 - 1)\Sigma_2 \boldsymbol{\beta} + \rho^2 (\boldsymbol{\beta} \circ \mathbf{v})^T (n_2 - 1)\Sigma_2 (\boldsymbol{\beta} \circ \mathbf{v}) + 2\alpha \boldsymbol{\beta}^{\mathbf{T}} (n_2 - 1)\Sigma_2 \boldsymbol{\eta}_{\mathbf{u}} + \boldsymbol{\eta}_{\mathbf{u}}^{\mathbf{T}} (n_2 - 1)\Sigma_2 \boldsymbol{\eta}_{\mathbf{u}} + 2\alpha \boldsymbol{\beta}^T (n_2 - 1)\Sigma_2 (\boldsymbol{\beta} \circ \mathbf{v}) + 2\rho (\boldsymbol{\beta} \circ \mathbf{v})^T (n_2 - 1)\Sigma_2 \boldsymbol{\eta}_{\mathbf{u}} - 2\rho (\boldsymbol{\beta} \circ \mathbf{v})^T (n_2 - 1)\widehat{\boldsymbol{\beta}}_{\mathbf{y}} - 2\alpha \boldsymbol{\beta}^T (n_2 - 1)\widehat{\boldsymbol{\beta}}_{\mathbf{y}} - \mathbf{2}\boldsymbol{\eta}_{\mathbf{u}}^{\mathbf{T}} (n_2 - 1)\widehat{\boldsymbol{\beta}}_{\mathbf{y}})/2$$

The posterior conditional distribution of $\sigma^2_\beta$ is an inverse gamma distribution with the shape parameter $\sigma^2_{\beta\_shape} = \frac{\sum_{j=1}^{p} \gamma_j}{2} + a_\beta$, and the scale parameter $\sigma^2_{\beta\_scale} = \frac{\sum_{j=1}^{p} \gamma_j \beta_j^2 + 2b_\beta}{2}$

The posterior conditional distribution of $\sigma^2_\eta$ is an inverse gamma distribution with the shape parameter $\sigma^2_{\eta\_shape} = \frac{\sum_{j=1}^{p} \tau_j}{2} + a_\eta$, and the scale parameter $\sigma^2_{\eta\_scale} = \frac{\sum_{j=1}^{p} \tau_j \eta_{u,j}^2 + 2b_\eta}{2}$.

#### Supplementary Figures

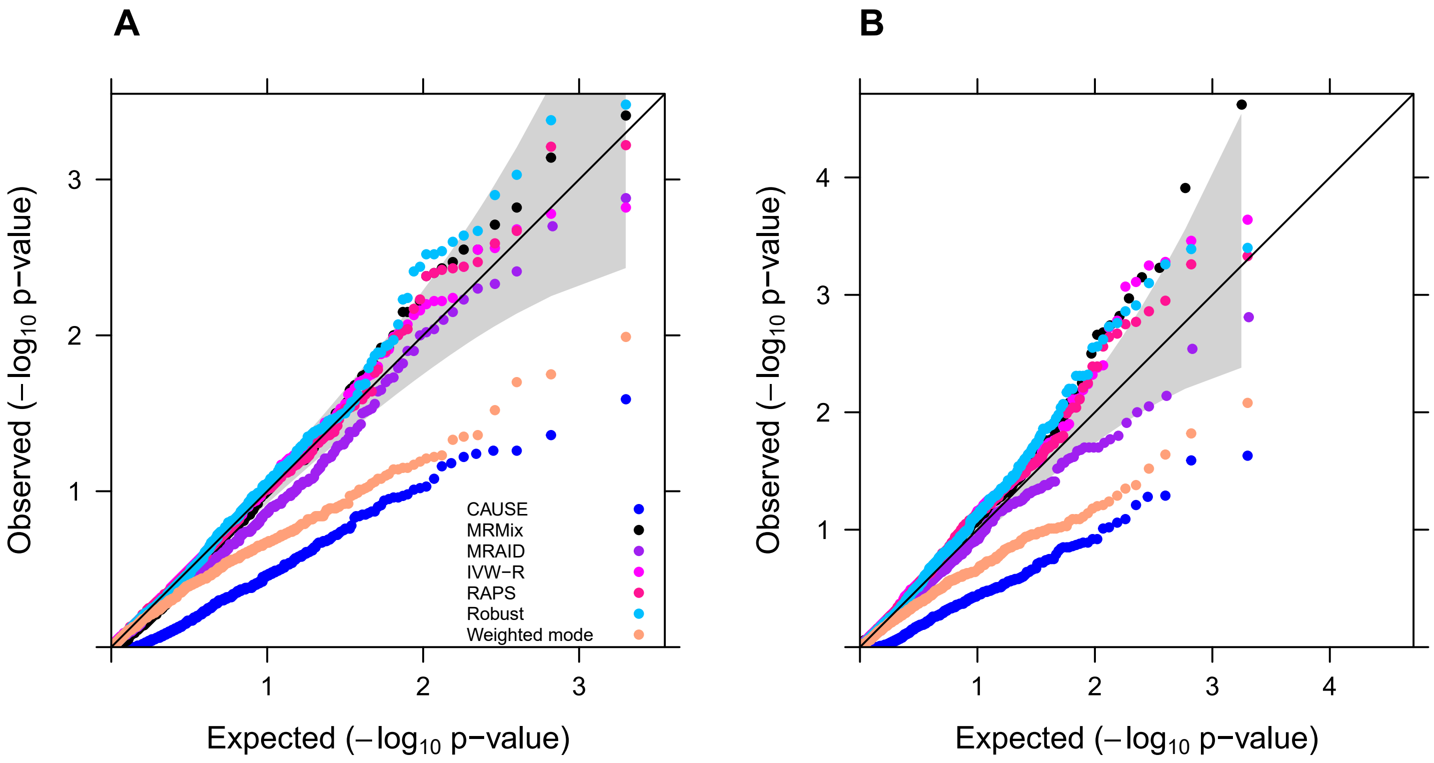


**Fig. S1 Type I error control of different MR methods in simulations.** Type I error control is evaluated by quantile-quantile plots of -log10 p-values from different MR methods on testing the causal effect in the absence of both correlated and uncorrelated horizontal pleiotropic effects under the null simulations. Compared methods include CAUSE (blue), MRMix (black), MRAID (purple), IVW-R (magenta), RAPS (deep pink), Robust (deep sky blue), Weighted mode (light salmon). (**A**) We simulated 1,000 instrumental SNPs with $PVE_{\tilde{Z}x}$ =10%; (**B**) We simulated 100 instrumental SNPs with $PVE_{\tilde{Z}x}$=5%.

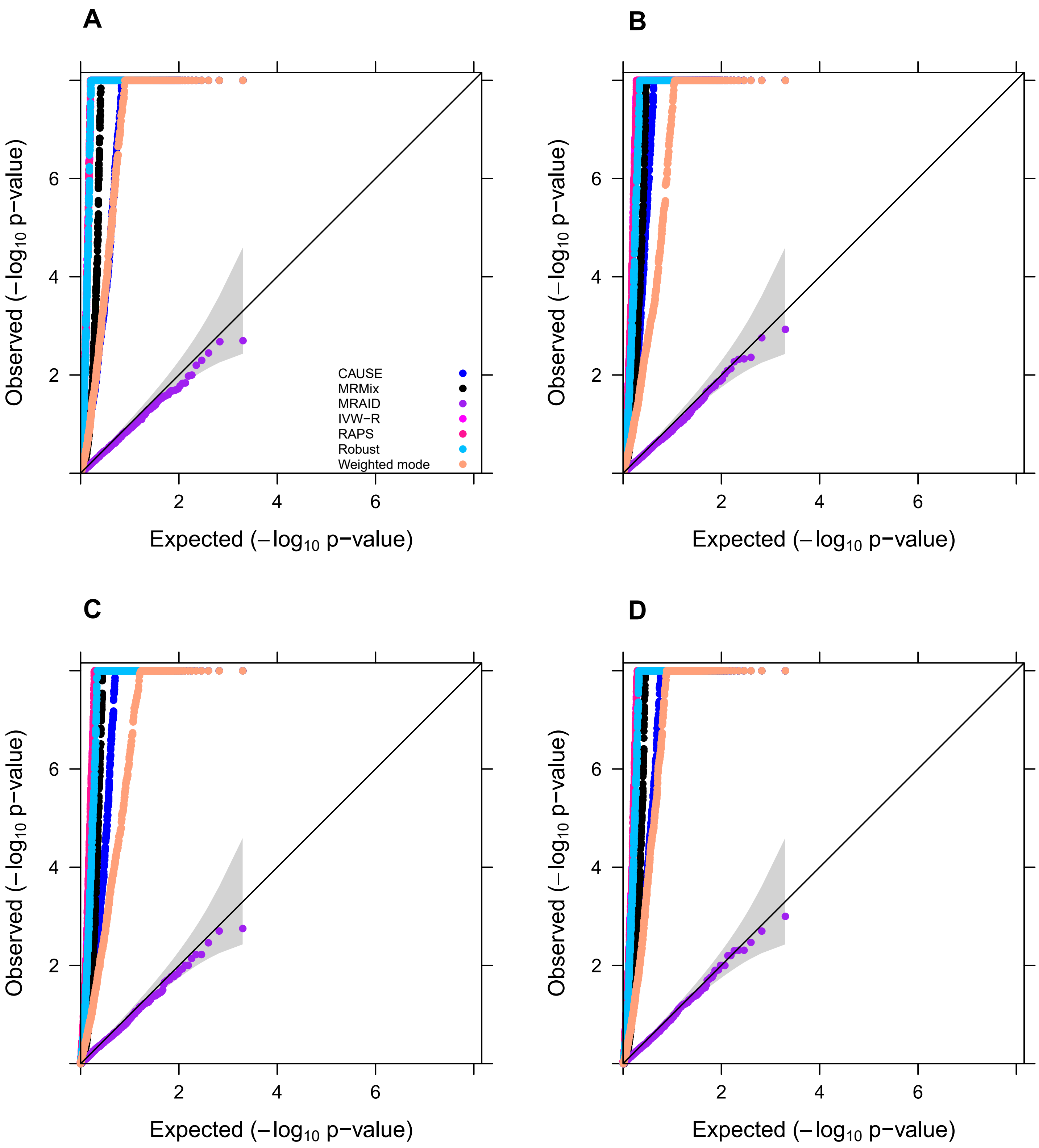


**Fig. S2 Type I error control of different MR methods in simulations.** Type I error control is evaluated by quantile-quantile plot of -log10 p-values from different MR methods on testing the causal effect in the absence of both correlated and uncorrelated horizontal pleiotropic effects under null simulations. All methods used all exposure-associated SNPs that achieve genome-wide significance level ($p < 5\times10^{-8}$) with high LD. Compared methods include CAUSE (blue), MRMix (black), MRAID (purple), IVW-R (magenta), RAPS (deep pink), Robust (deep sky blue), Weighted mode (light salmon). (**A**) We simulated 100 instrumental SNPs with their effect sizes drawing from a normal distribution; (**B**) We simulated 1,000 instrumental SNPs with their effects size drawing from a normal distribution; (**C**) We simulated 1,000 instrumental SNPs with their effects size drawing from a BSLMM distribution with 10% SNPs having large effects and 90% SNPs having small effects; (**D**) We simulated 1,000 instrumental SNPs with their effects size drawing from a BSLMM distribution with 1% SNPs having large effects and 99% SNPs having small effects.

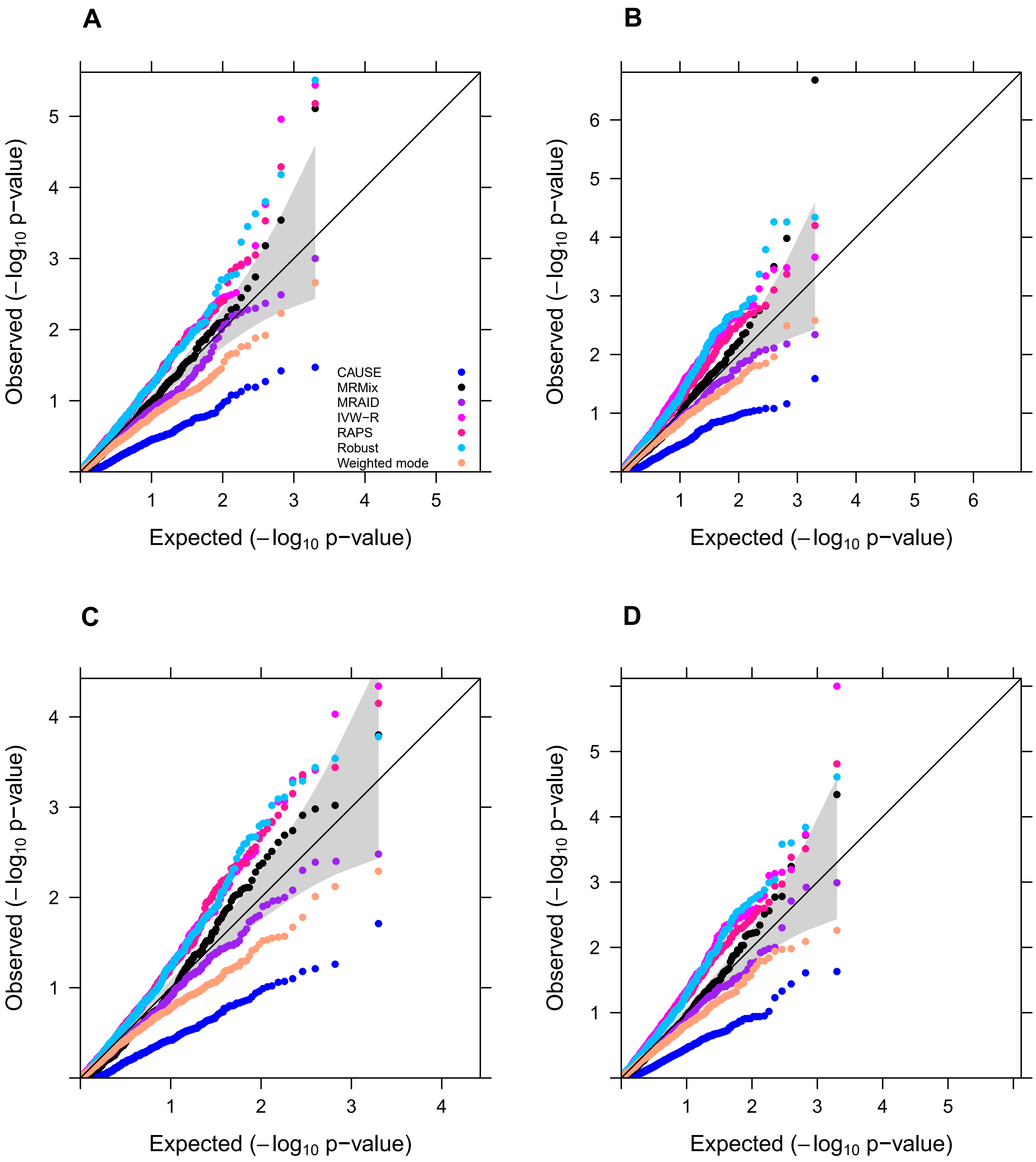


**Fig. S3 Type I error control of different MR methods in simulations.** Type I error control is evaluated by quantile-quantile plot of -log10 p-values from different MR methods on testing the causal effect under the null simulations in the absence of correlated horizontal pleiotropic effects but in the presence of uncorrelated horizontal pleiotropic effects. Compared methods include CAUSE (blue), MRMix (black), MRAID (purple), IVW-R (magenta), RAPS (deep pink), Robust (deep sky blue), Weighted mode (light salmon). We simulated 100 instrumental SNPs with different uncorrelated horizontal pleiotropy effects ($PVE_u$=5%, **A**, **B**; $PVE_u$=2.5%, **C**, **D**) and set the proportions of instrumental SNPs having the uncorrelated horizontal pleiotropy effect to be either no overlap (**A**, **C**) or 10% (**B**, **D**).

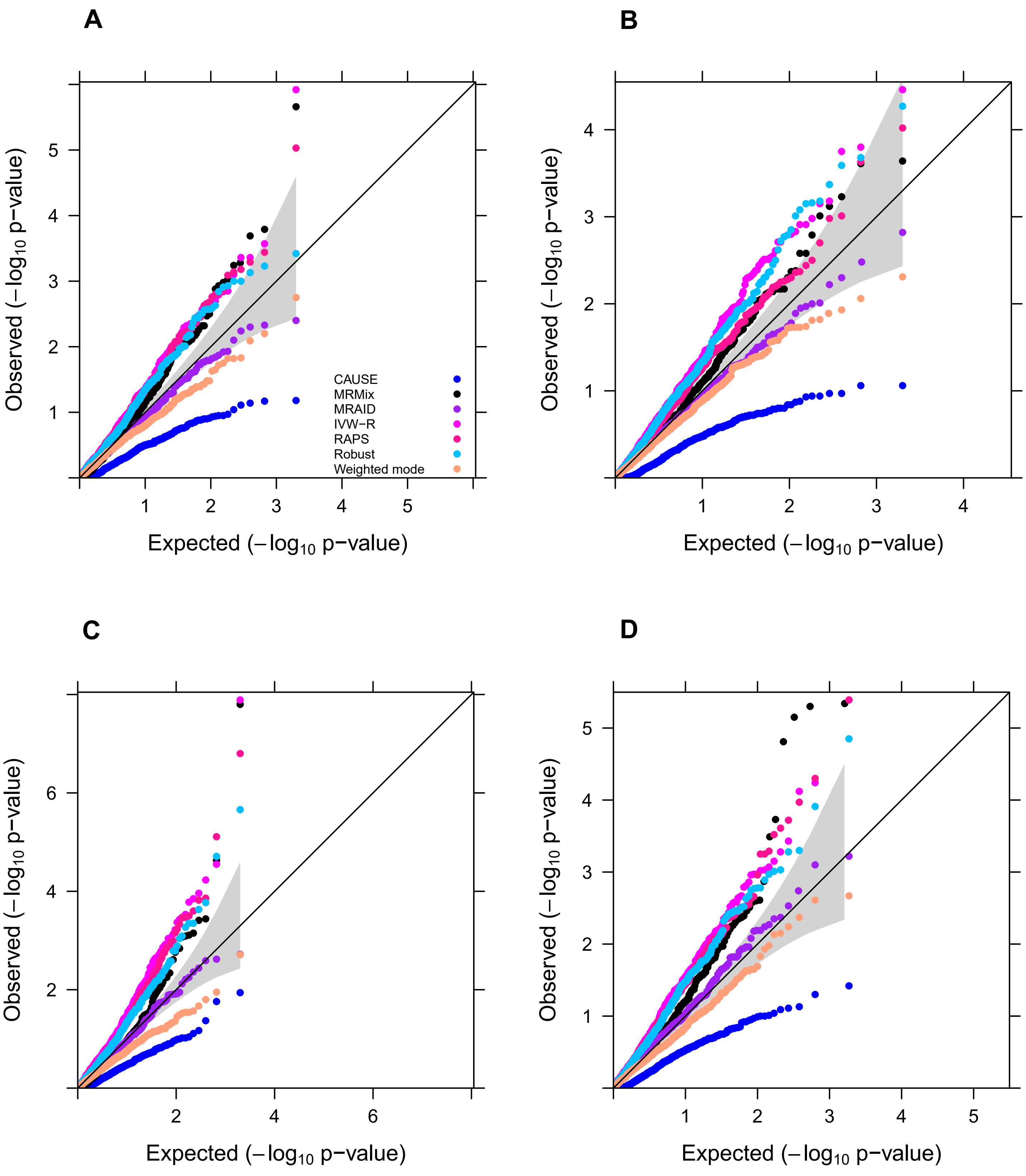


**Fig. S4 Type I error control of different MR methods in simulations.** Type I error control is evaluated by quantile-quantile plot of -log10 p-values from different MR methods on testing the causal effect under the null simulations in the presence of both correlated and uncorrelated horizontal pleiotropic effects ($PVE_u$=5%). Compared methods include CAUSE (blue), MRMix (black), MRAID (purple), IVW-R (magenta), RAPS (deep pink), Robust (deep sky blue), Weighted mode (light salmon). We simulated 100 instrumental SNPs with different proportions of instrumental SNPs having uncorrelated horizontal pleiotropy effects (no overlap; **A**, **C**, **D**; 20%; **B**), different probability of having the correlated pleiotropy effect from an unobserved confounder ($\pi_c$=5%, **A**, **B**, **C**; $\pi_c$=10%, **D**) and different correlated pleiotropy effect sizes $\rho=\sqrt{0.02}$ (**A**, **B**, **D**) or $\rho=\sqrt{0.05}$ (**C**).

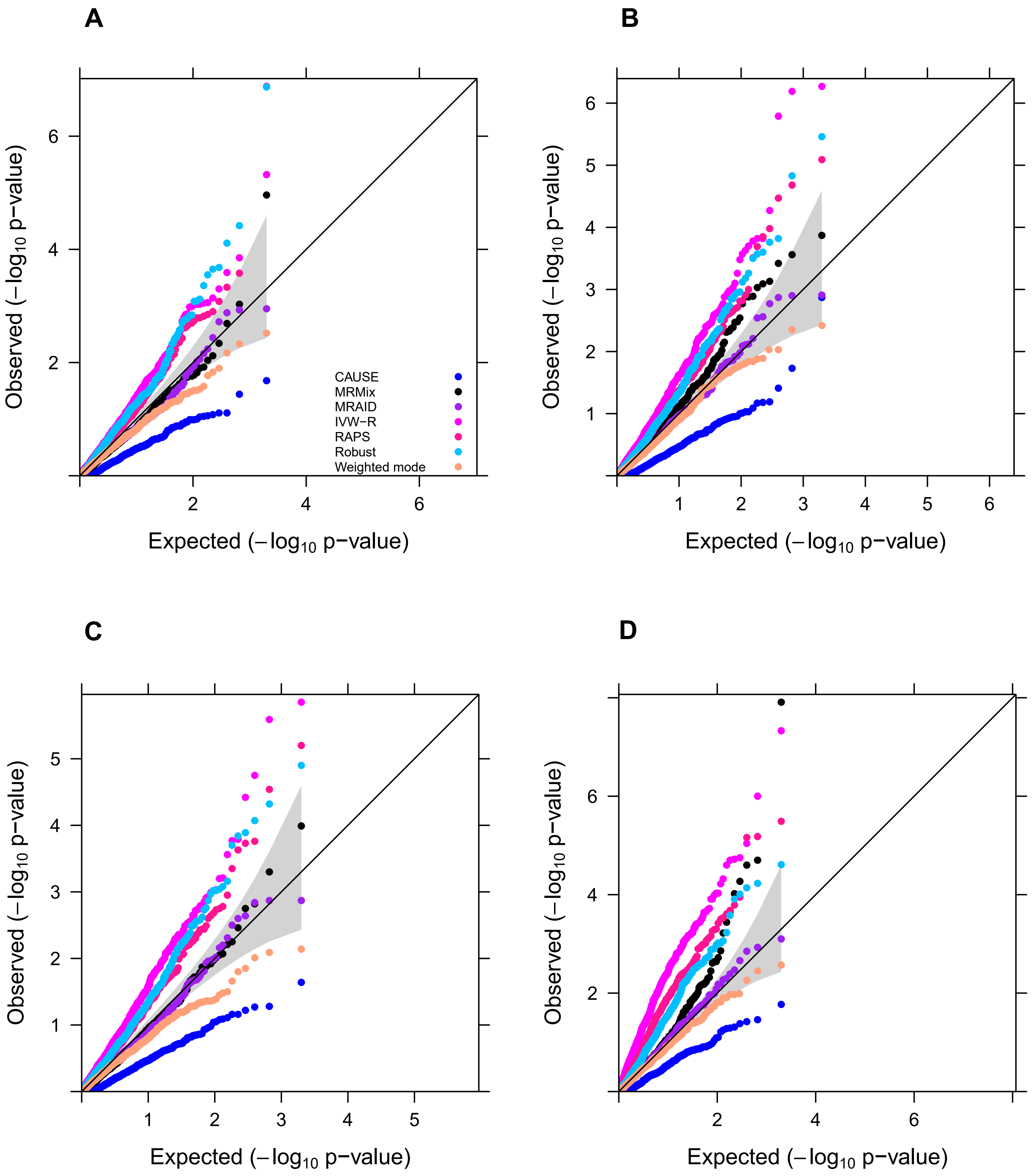


**Fig. S5 Type I error control of different MR methods in simulations.** Type I error control is evaluated by quantile-quantile plot of -log10 p-values from different MR methods on testing the causal effect under the null simulations in the presence of both correlated and uncorrelated horizontal pleiotropic effect ($PVE_u$=5%). Compared methods include CAUSE (blue), MRMix (black), MRAID (purple), IVW-R (magenta), RAPS (deep pink), Robust (deep sky blue), Weighted mode (light salmon). We simulated 100 instrumental SNPs with different overlapped proportion among instrumental SNPs and the uncorrelated pleiotropic SNPs (10%, **A**, **C**; 30%, **B**, **D**). The correlated horizontal pleiotropy is, rather than simulated from an unobserved confounder, generated through assigning correlation either 0.1 (**A**, **B**) or 0.3 (**C**, **D**) to the effect size of the overlapped SNPs from bivariate normal distribution.

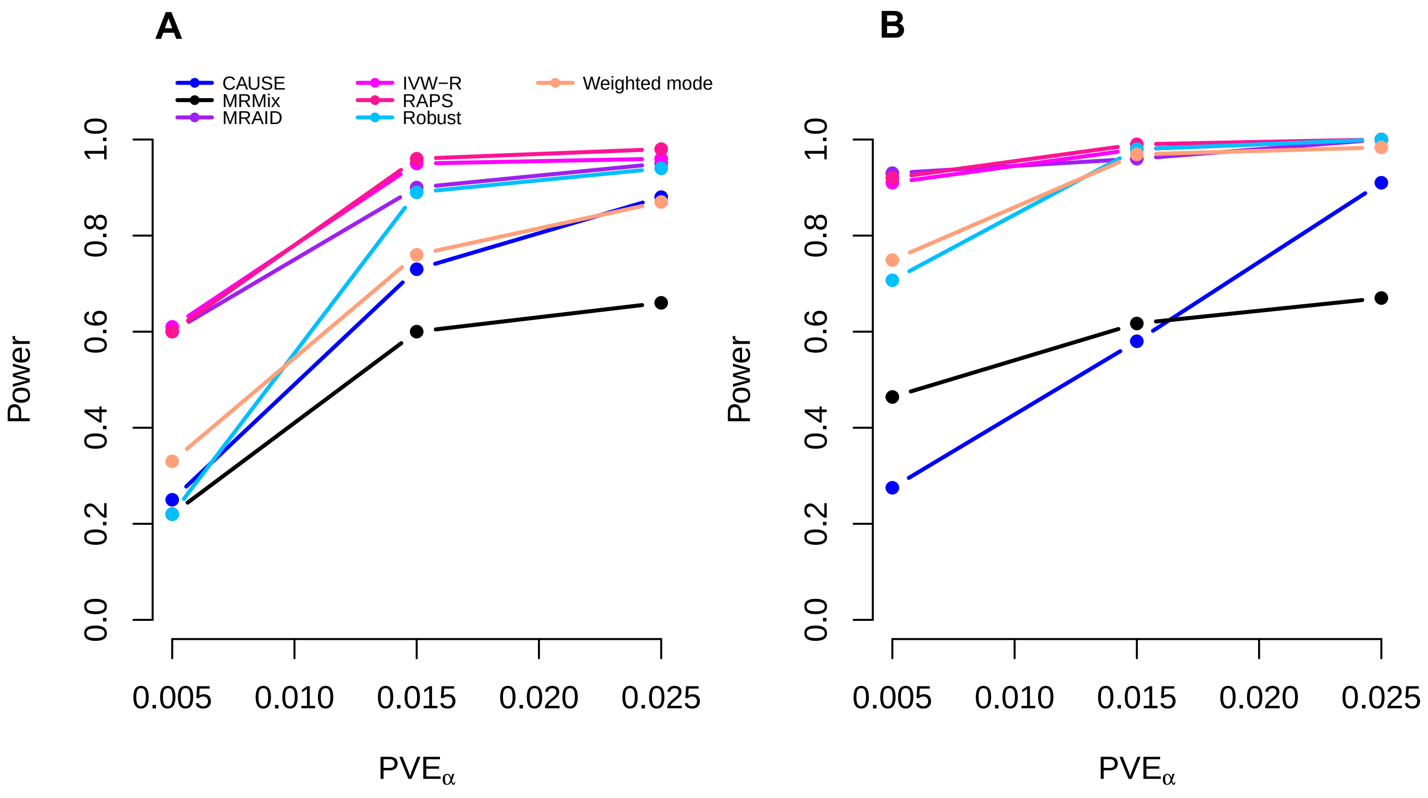


**Fig. S6 Power of different MR methods in the absence of both uncorrelated and correlated pleiotropic effects.** Power (y-axis) at a false discovery rate of 0.05 to detect the causal effect is plotted against different causal effect size characterized by $PVE_{\alpha}$ (x-axis). Compared methods include CAUSE (blue), MRMix (black), MRAID (purple), IVW-R (magenta), RAPS (deep pink), Robust (deep sky blue), Weighted mode (light salmon). (**A**) We simulated 1,000 instrumental SNPs with their effect size drawing from a normal distribution; (**B**) We simulated 1,000 instrumental SNPs with their effect size drawing from a BSLMM distribution with 10% SNPs having large effects and 90% SNPs having small effects.

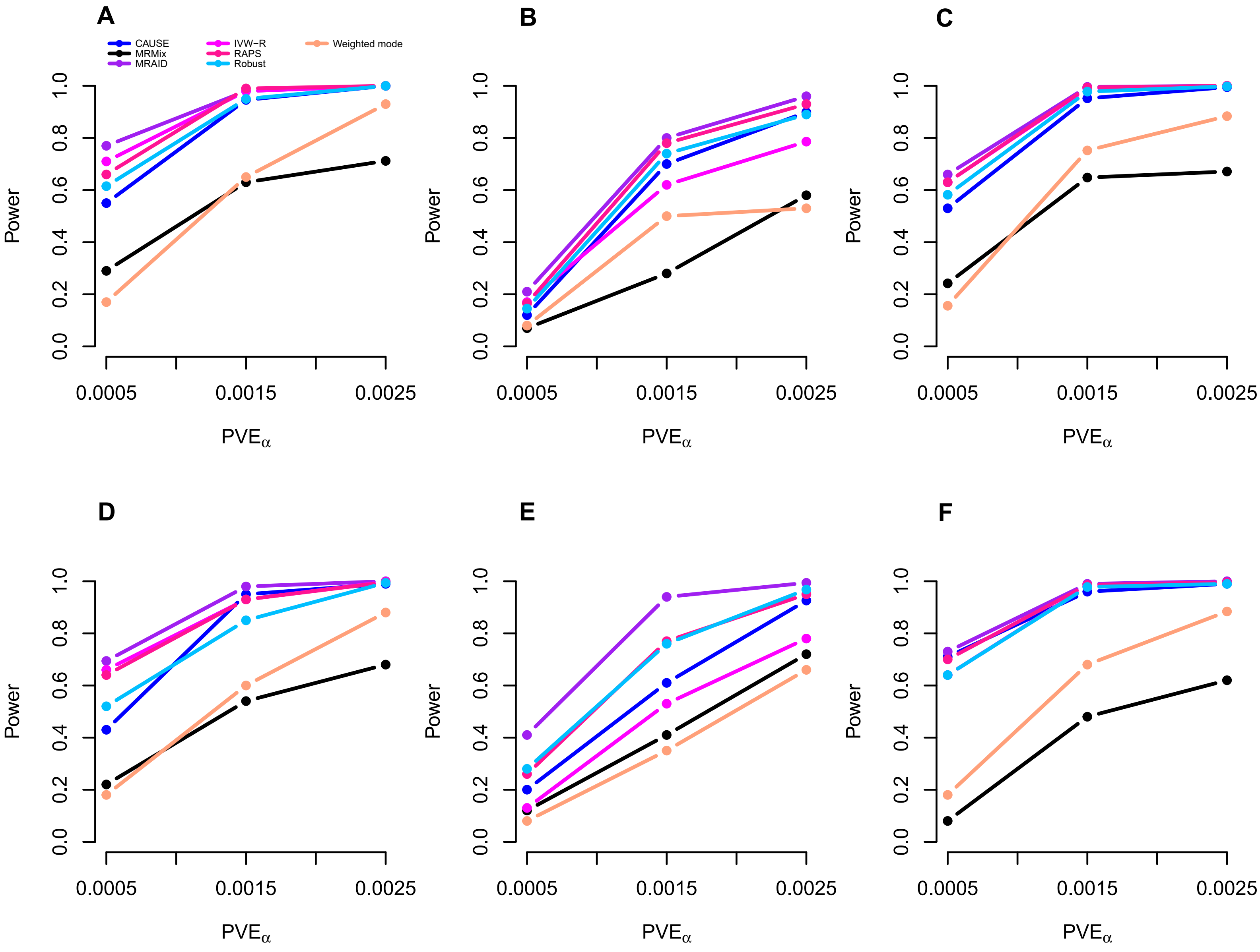


**Fig. S7 Power of different MR methods in simulations.** Power (y-axis) at a false discovery rate of 0.05 to detect the causal effect is plotted against different causal effect size characterized by $PVE_\alpha$ (x-axis). Compared methods include CAUSE (blue), MRMix (black), MRAID (purple), IVW-R (magenta), RAPS (deep pink), Robust (deep sky blue), Weighted mode (light salmon). We simulated 100 instrumental SNPs with different uncorrelated horizontal pleiotropy effects ($PVE_u$=2.5%; **A**, **B**; $PVE_u$=5%; **C**, **D**, **E**, **F**), different proportions of instrumental SNPs having the uncorrelated horizontal pleiotropy effects (no overlap, **A**, **C**, **D**, **F**; 20%, **E**; 30%, **B**), and different correlated horizontal pleiotropy effect sizes ($\pi_c$=0, $\rho$=0, **A**, **B**, **C**; $\pi_c$=5%, $\rho=\sqrt{0.02}$, **D**, **E**; $\pi_c$=5%, $\rho=\sqrt{0.05}$, **F**).

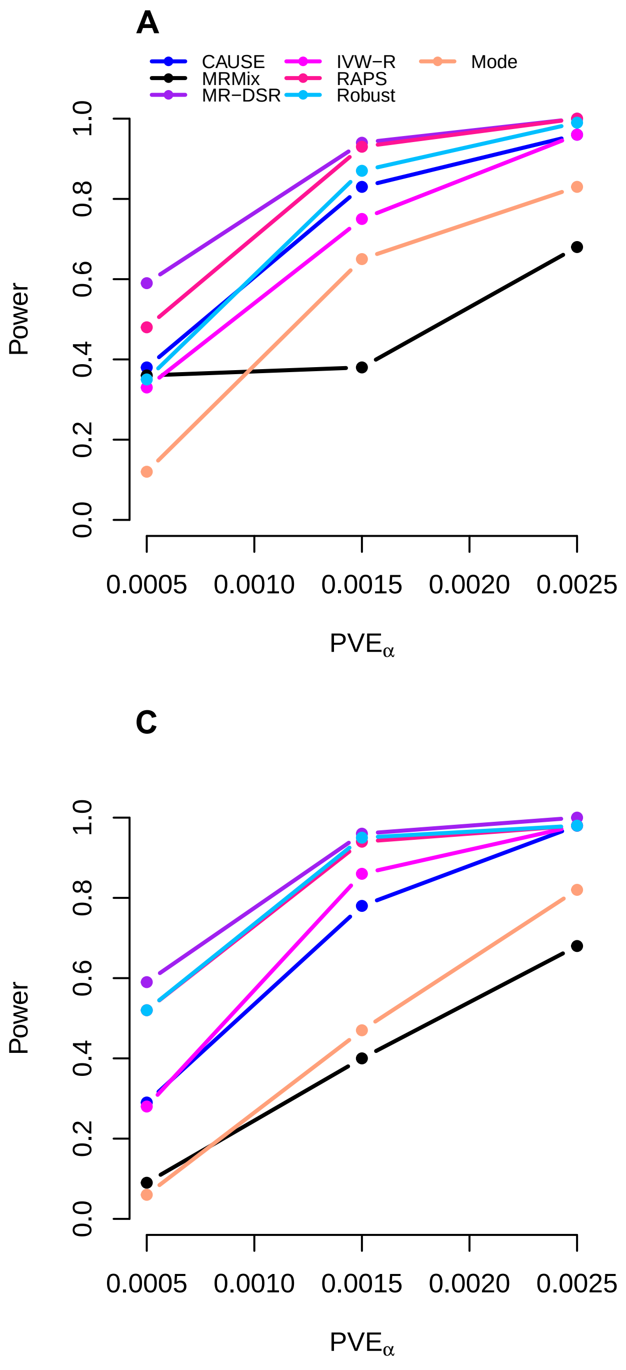


**Fig. S8 Power of different methods in the presence of both correlated and uncorrelated horizontal pleiotropic effect ($PVE_u$=5%).** Power (y-axis) at a false discovery rate of 0.05 to detect the causal effect is plotted against different causal effect size characterized by $PVE_\alpha$ (x-axis). Compared methods include CAUSE (blue), MRMix (black), MRAID (purple), IVW-R (magenta), RAPS (deep pink), Robust (deep sky blue), Weighted mode (light salmon). We simulated 100 instrumental SNPs with different overlapped proportion among instrumental SNPs and the pleiotropic SNPs (10%, **A**, **C**; 30%, **B**, **D**). The correlated horizontal pleiotropy is, rather than simulated from an unobserved confounder, generated through assigning correlation either 0.1 (**A**, **B**) or 0.3 (**C**, **D**) to the effect size of the overlapped SNPs from bivariate normal distribution.

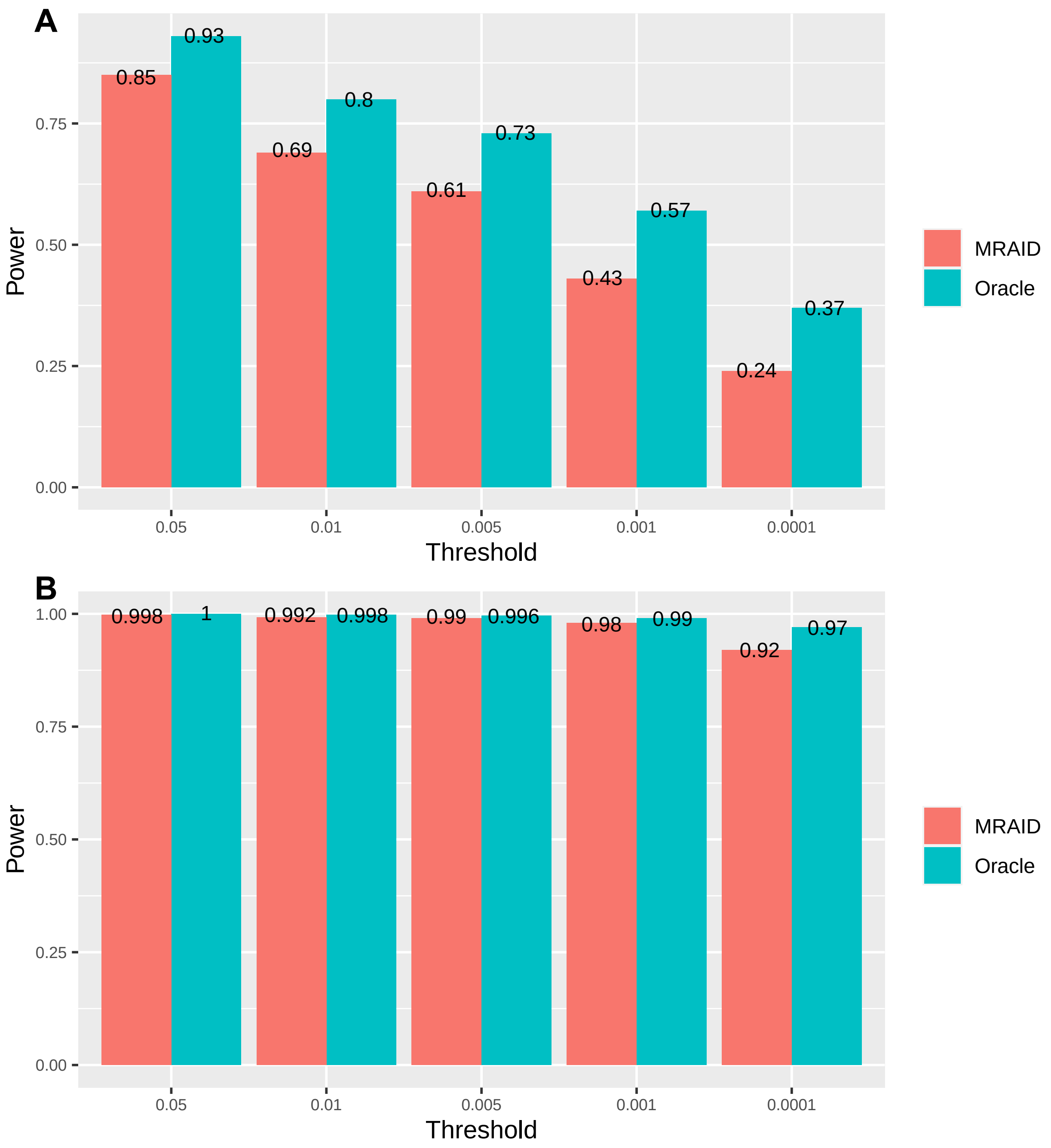


**Fig. S9 Power of MRAID and Oracle method in the absence of both uncorrelated and correlated pleiotropic effects.** Power (y-axis) to detect the causal effect is plotted against different p value threshold (x-axis). Compared methods include MRAID (red) and Oracle (blue). We simulated 100 instrumental SNPs with 500 replicates. The power of the oracle method is calculated from the IVW-R method using the independent SNPs screened from the 100 instruments, which is the gold standard method in the absence of any type of horizontal pleiotropy effect. (**A**) simulations with $PVE_{\alpha}$=0.05%; (**B**) simulations with $PVE_{\alpha}$=0.15%.

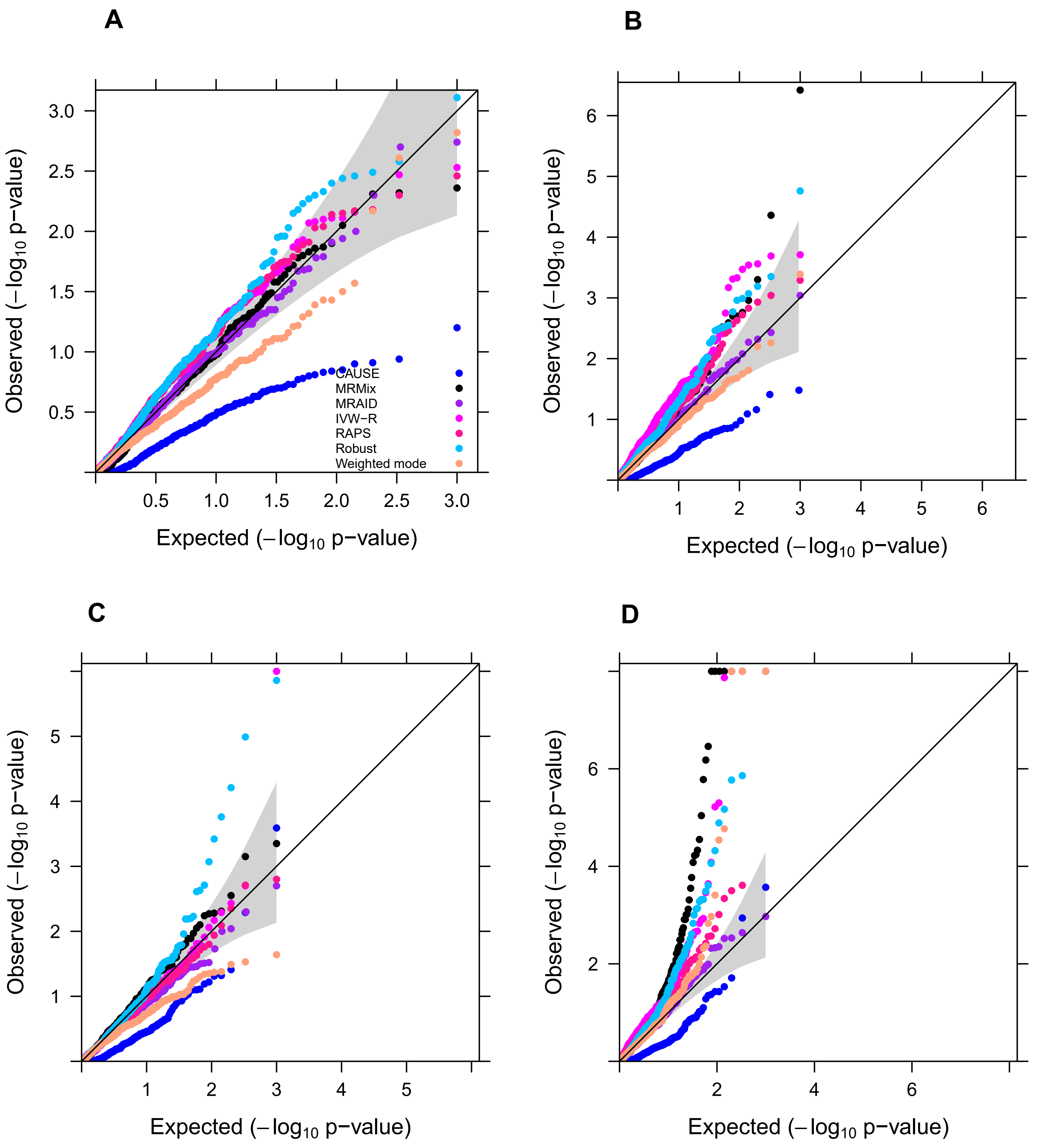


**Fig. S10 Quantile-quantile plot of -log10 p-values from different MR methods in distinguishing the causal effect direction through reverse causality analysis in the absence of correlated horizontal pleiotropic effects but in the presence of uncorrelated horizontal pleiotropic effects.** We tested the causal effect of the outcome on the exposure in the alternative simulations where the exposure has causal effect on the outcome ($PVE_{\alpha}$=0.15%) but not vice versa. Compared methods include CAUSE (blue), MRMix (black), MRAID (purple), IVW-R (magenta), RAPS (deep pink), Robust (deep sky blue), Weighted mode (light salmon). We simulated 100 instrumental SNPs with different uncorrelated horizontal pleiotropy effects ($PVE_{u}$=5%, **A**, **B**; $PVE_{u}$=2.5%, **C**, **D**) and set the proportions of instrumental SNPs having the uncorrelated horizontal pleiotropy effect to be either no overlap (**A**, **C**) or 30% (**B**, **D**). MRAID is the only method showing calibrated p values across a range of simulation scenarios.

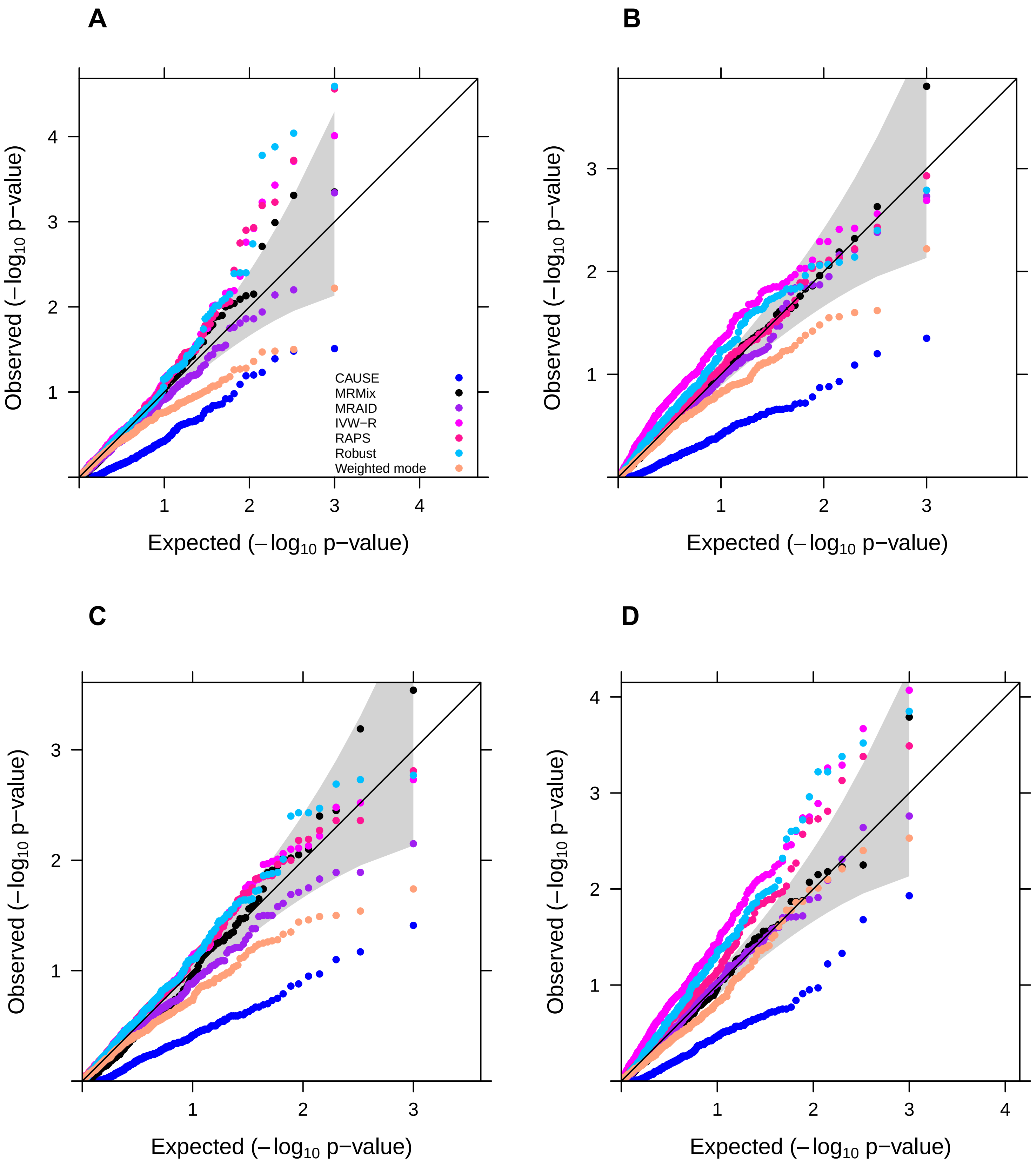


**Fig. S11 Quantile-quantile plot of -log10 p-values from different MR methods in distinguishing the causal effect direction through reverse causality analysis in the presence of both correlated ($\pi_c$=5%) and uncorrelated horizontal pleiotropic effect (*$PVE_u$*=5%).** We tested the causal effect of the outcome on the exposure in the alternative simulations where the exposure has causal effect on the outcome (*$PVE_\alpha$*=0.15%) but not vice versa. Compared methods include CAUSE (blue), MRMix (black), MRAID (purple), IVW-R (magenta), RAPS (deep pink), Robust (deep sky blue), Weighted mode (light salmon). We simulated 100 instrumental SNPs with different proportions of instrumental SNPs having the uncorrelated horizontal pleiotropy effect being either no overlap (**A**, **C**) or 20% (**B**, **D**) and different correlated horizontal pleiotropy effect sizes ($\rho=\sqrt{0.05}$, **A**, **B**; $\rho=\sqrt{0.02}$, **C**, **D**). MRAID is the only method showing calibrated p values across a range of simulation scenarios.

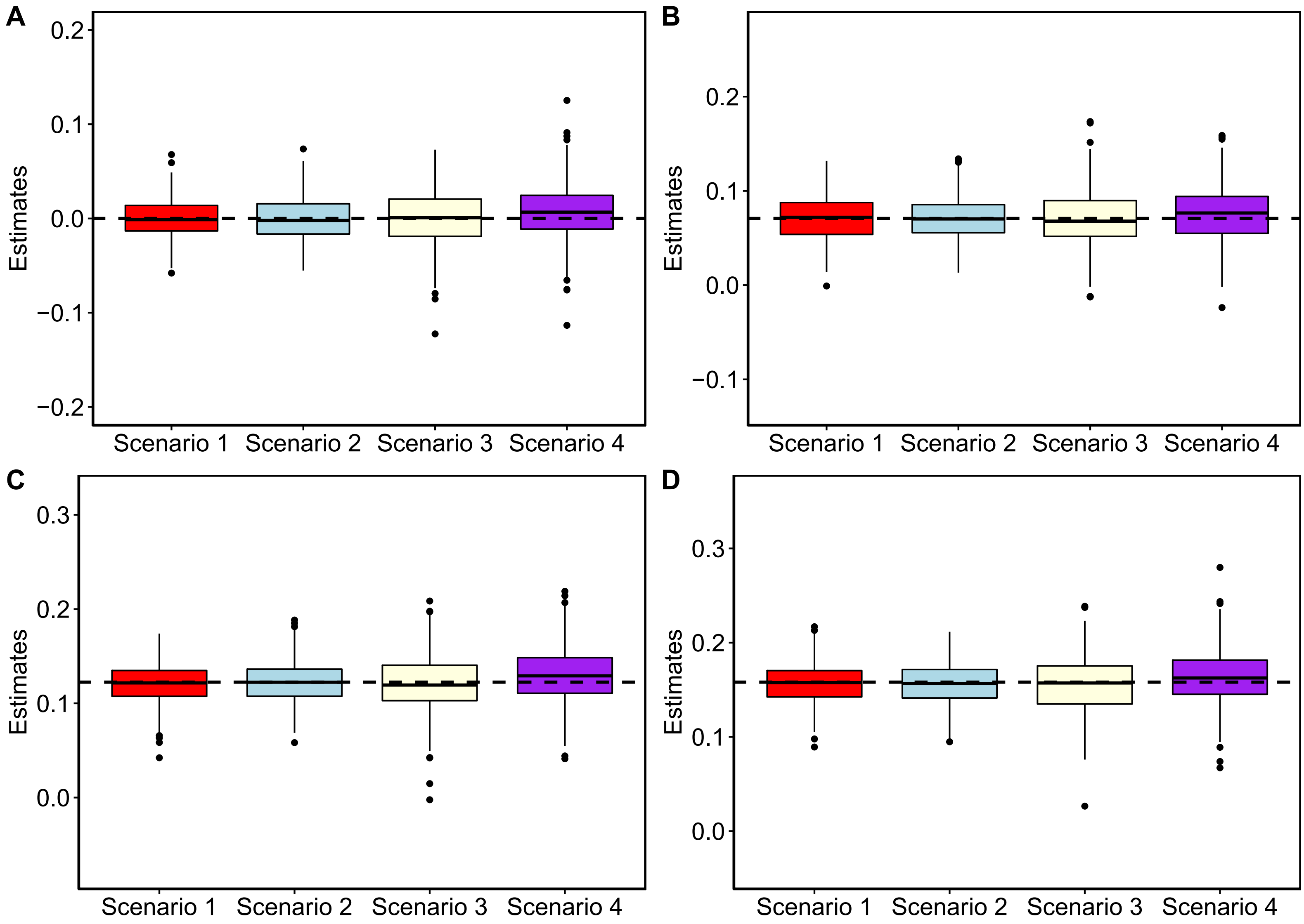


**Fig. S12 Quantile-quantile plot of -log10 p-values from different MR methods in distinguishing the causal effect direction through reverse causality analysis in the presence of both correlated ($\pi_c$=5%) and uncorrelated horizontal pleiotropic effect ($PVE_u$=5%).** We tested the causal effect of the outcome on the exposure in the alternative simulations where the exposure has causal effect on the outcome ($PVE_\alpha$=0.15%) but not vice versa. Compared methods include CAUSE (blue), MRMix (black), MRAID (purple), IVW-R (magenta), RAPS (deep pink), Robust (deep sky blue), Weighted mode (light salmon). We simulated 100 instrumental SNPs with different proportions of instrumental SNPs having the uncorrelated horizontal pleiotropy effect being either no overlap (**A**, **C**) or 20% (**B**, **D**) and different correlated horizontal pleiotropy effect sizes ($\rho=\sqrt{0.05}$, **A**, **B**; $\rho=\sqrt{0.02}$, **C**, **D**). MRAID is the only method showing calibrated p values across a range of simulation scenarios.

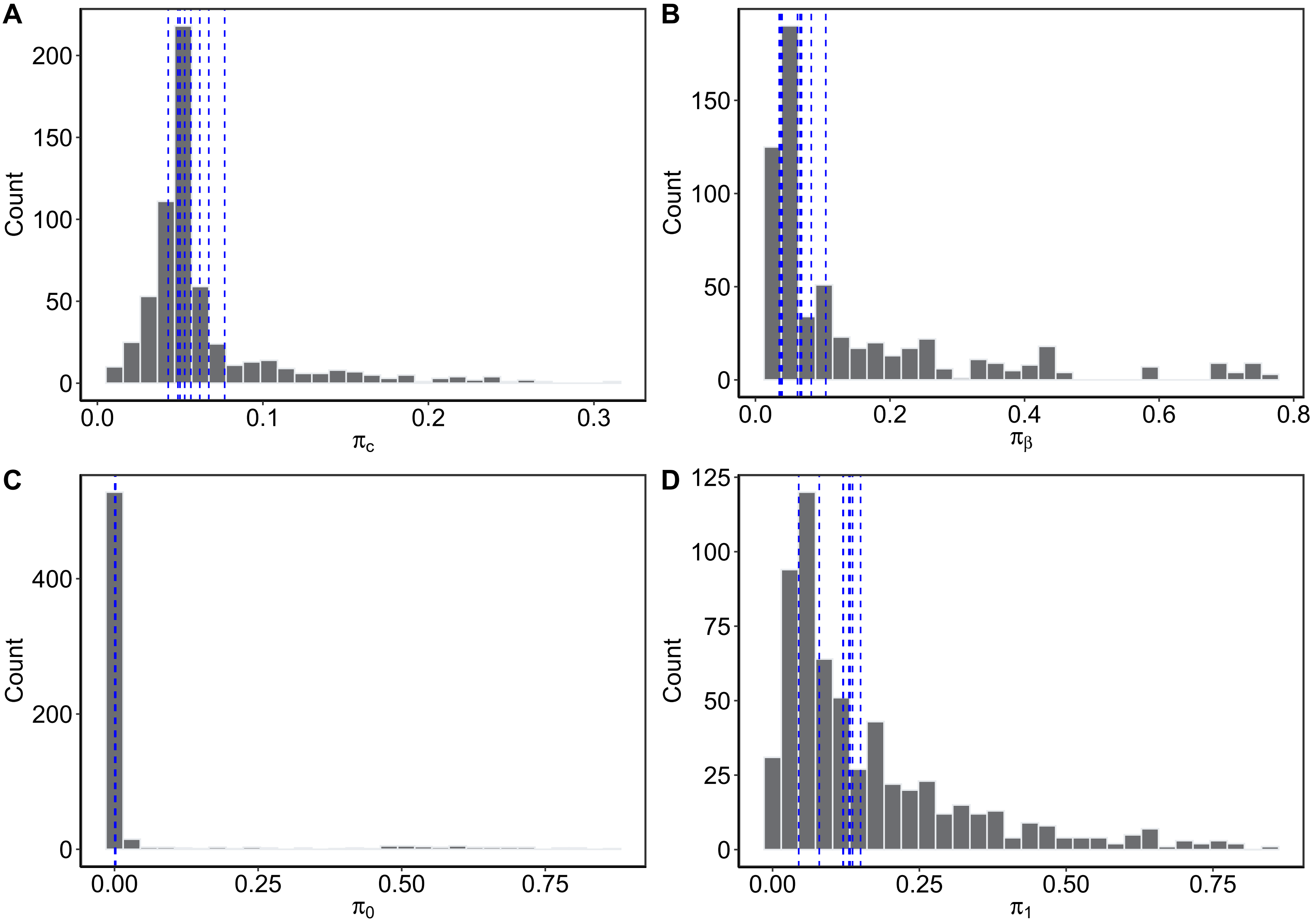


**Fig. S13 Histograms of the four proportion parameters in MARID across the 645 analyzed trait pairs from UK Biobank, with blue dashed line in each panel representing the value from the 8 significant trait pairs detected by MRAID.** Histograms are for $\pi_c$ (**A**), $\pi_\beta$ (**B**), $\pi_0$ (**C**) and $\pi_1$ (**D**) respectively.

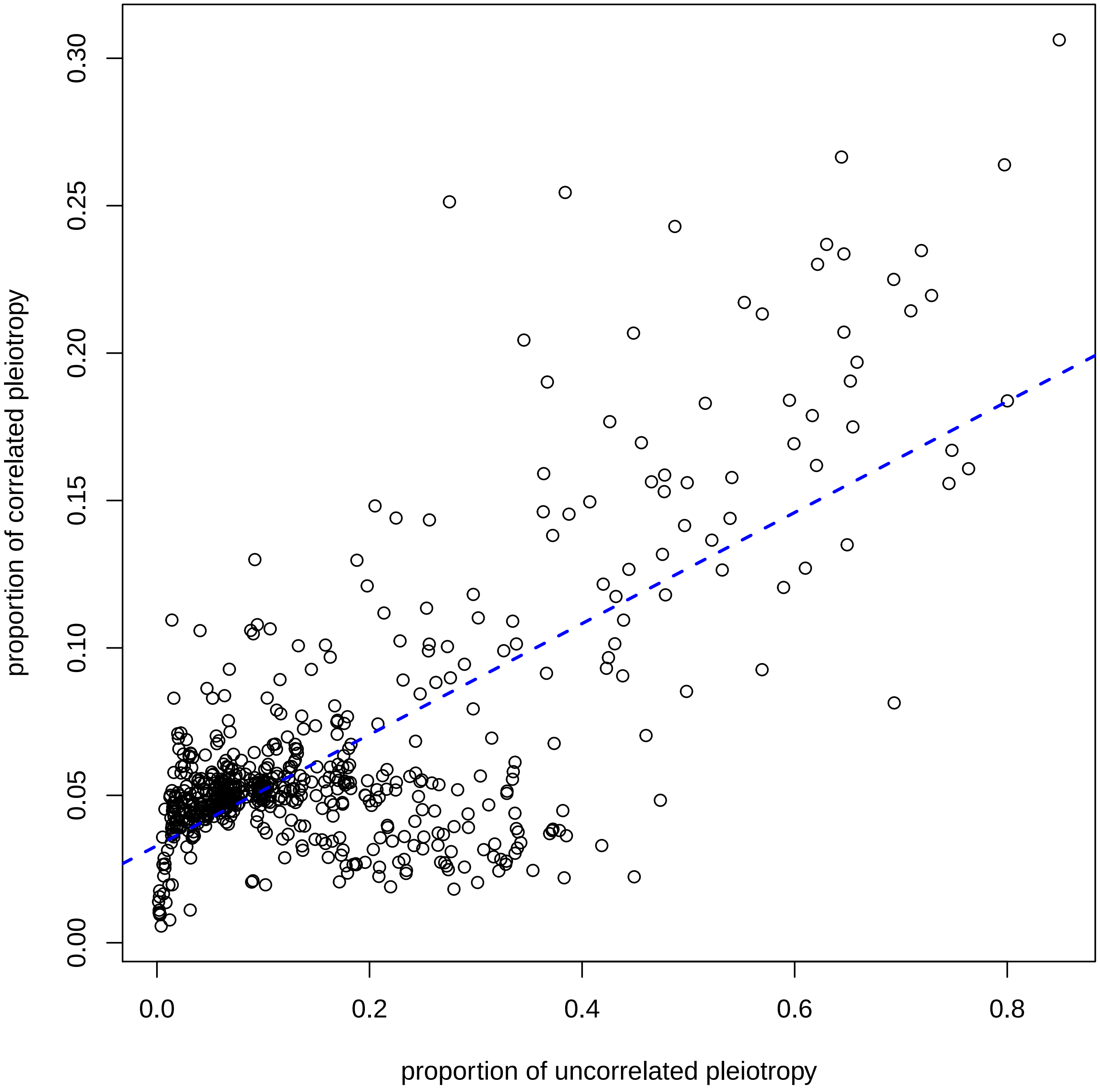


**Fig. S14 Scatterplot displays the relationship between the proportion of SNPs exhibiting uncorrelated horizontal pleiotropy among the selected instruments ($\pi_1$) and the proportions of SNPs displaying correlated horizontal pleiotropy among the selected instruments ($\pi_c$) across the 645 analyzed trait pairs from UK Biobank.** The proportion of SNPs displaying correlated pleiotropy is correlated with the proportions of SNPs displaying uncorrelated pleiotropy, with the latter generally being larger than the former.

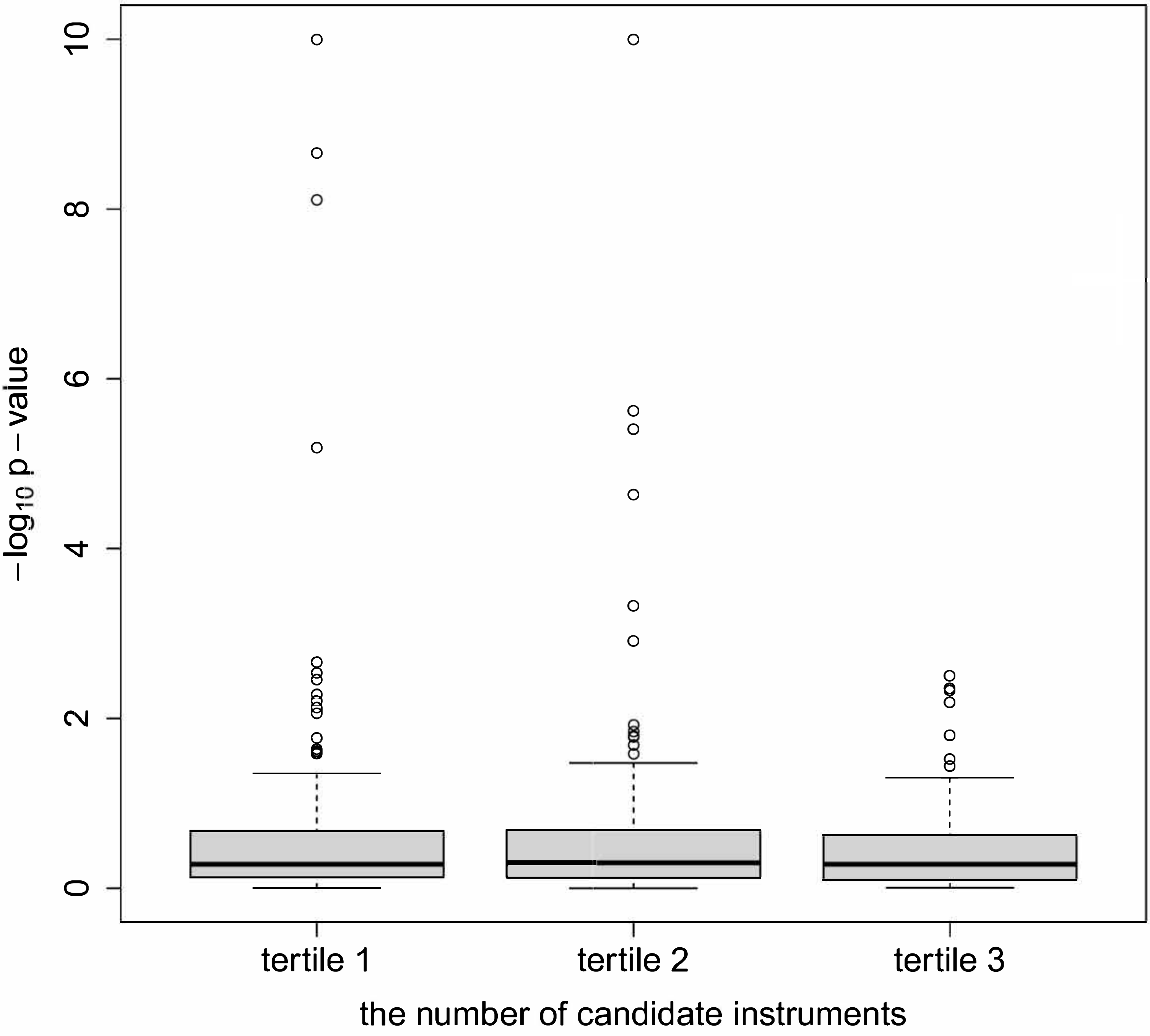


**Fig. S15 Boxplot displays the -log10 p-values of MRAID stratified by the tertiles of the number of candidate instruments across the 645 analyzed trait pairs from UK Biobank.**

### Supplementary Tables

#### Table S1. All causal trait associations detected by different MR methods.

| Traits | MRAID | IVW-R | RAPS | MRMix | CAUSE | Robust | Weighted mode |
|---|---|---|---|---|---|---|---|
| Age started smoking in former smokers | | | | | | | |
| PR | 0.67 (0.65) | -0.40 ($2.2\times10^{-4}$) | -0.40 ($5.28\times10^{-4}$) | -0.40 ($3.0\times10^{-4}$) | **-0.30 ($1.3\times10^{-11}$)** | **-0.40 ($3.1\times10^{-15}$)** | -0.39 ($1.3\times10^{-3}$) |
| PWRIx | 0.46 (0.62) | 0.35 ($1.1\times10^{-3}$) | 0.35 ($2.0\times10^{-3}$) | 0.36 ($1.2\times10^{-3}$) | 0.28 ($1.5\times10^{-3}$) | **0.35 ($2.7\times10^{-12}$)** | 0.37 ($2.4\times10^{-3}$) |
| PWP2PT | 0.24 (0.41) | 0.14 (0.19) | 0.14 (0.21) | 0.14 (0.19) | **0.03 ($7.2\times10^{-57}$)** | 0.14 ($5.5\times10^{-3}$) | 0.14 (0.25) |
| BMI | **2.80 (0)** | **2.64 (0)** | **2.65 (0)** | 0.00 (1) | **4.57 (0)** | **2.64 ($8.9\times10^{-54}$)** | **2.68 ($2.9\times10^{-48}$)** |
| SBP | -0.42 (0.06) | **-0.34 ($7.8\times10^{-8}$)** | **-0.34 ($8.9\times10^{-7}$)** | **-0.35 ($4.1\times10^{-7}$)** | -0.33 ($1.3\times10^{-3}$) | **-0.34 ($1.5\times10^{-30}$)** | **-0.35 ($1.7\times10^{-6}$)** |
| DBP | -0.14 (0.14) | -0.14 (0.03) | -0.14 (0.04) | -0.14 (0.03) | -0.04 (0.02) | **-0.14 ($3.3\times10^{-6}$)** | -0.15 (0.03) |
| TC | -0.19 (0.75) | 0.18 ($3.7\times10^{-3}$) | 0.18 ($5.9\times10^{-3}$) | 0.19 ($4.0\times10^{-3}$) | 0.11 ($4.0\times10^{-3}$) | **0.18 ($5.6\times10^{-10}$)** | 0.19 ($5.6\times10^{-3}$) |
| HDL-C | **0.70 ($2.2\times10^{-9}$)** | **0.70 ($8.0\times10^{-26}$)** | **0.70 (0)** | **0.70 ($7.1\times10^{-18}$)** | 0.13 (0.11) | **0.70 ($1.1\times10^{-111}$)** | **0.70 ($5.4\times10^{-16}$)** |
| LDL-C | 0.06 (0.81) | 0.13 (0.04) | 0.13 (0.05) | 0.14 (0.04) | 0.05 ($3.6\times10^{-3}$) | **0.13 ($1.0\times10^{-5}$)** | 0.14 (0.05) |
| TG | **-0.64 ($7.8\times10^{-9}$)** | **-0.65 ($6.0\times10^{-25}$)** | **-0.65 (0)** | **-0.66 ($3.9\times10^{-17}$)** | **-0.92 ($4.9\times10^{-267}$)** | **-0.65 ($1.0\times10^{-107}$)** | **-0.65 ($1.4\times10^{-15}$)** |
| Number of unsuccessful stop-smoking attempts | | | | | | | |
| PR | 0.20 (0.31) | 0.23 ($3.8\times10^{-3}$) | 0.23 ($5.5\times10^{-3}$) | 0.23 ($4.2\times10^{-3}$) | 0.20 (0.03) | **0.23 ($6.0\times10^{-14}$)** | 0.23 (0.01) |
| PWRIx | -0.56 (0.59) | -0.20 (0.01) | -0.20 (0.02) | -0.21 (0.01) | -0.14 (0.06) | **-0.20 ($4.7\times10^{-5}$)** | -0.23 (0.02) |
| BMI | **-1.94 (0)** | **-1.82 ($1.8\times10^{-38}$)** | **-1.84 (0)** | -1.00 ($1.9\times10^{-4}$) | -1.93 ($1.8\times10^{-3}$) | **-1.82 ($7.0\times10^{-59}$)** | **-1.91 ($2.7\times10^{-62}$)** |
| SBP | 0.15 (0.44) | **0.23 ($1.4\times10^{-6}$)** | **0.23 ($5.5\times10^{-6}$)** | **0.23 ($2.8\times10^{-6}$)** | 0.19 (0.01) | **0.23 ($4.1\times10^{-34}$)** | 0.22 ($1.3\times10^{-4}$) |
| TC | -0.15 (0.07) | -0.13 ($5.3\times10^{-3}$) | -0.13 ($7.6\times10^{-3}$) | -0.13 ($5.5\times10^{-3}$) | -0.11 (0.08) | **-0.13 ($6.7\times10^{-6}$)** | -0.14 (0.02) |
| HDL-C | **-0.46 ($2.4\times10^{-6}$)** | **-0.44 ($1.1\times10^{-19}$)** | **-0.45 ($4.4\times10^{-16}$)** | **-0.45 ($2.2\times10^{-16}$)** | -0.24 ($1.38\times10^{-3}$) | **-0.44 ($1.3\times10^{-38}$)** | **-0.48 ($4.8\times10^{-14}$)** |
| LDL-C | -0.07 (0.66) | -0.11 (0.02) | -0.11 (0.03) | -0.11 (0.02) | -0.08 (0.07) | **-0.11 ($2.6\times10^{-5}$)** | -0.11 (0.06) |
| TG | **0.48 ($3.9\times10^{-6}$)** | **0.48 ($3.0\times10^{-12}$)** | **0.49 ($2.1\times10^{-14}$)** | 0.00 (1) | 0.40 ($1.9\times10^{-3}$) | **0.48 ($9.6\times10^{-27}$)** | **0.48 ($5.0\times10^{-14}$)** |
| Number of cigarettes previously smoked daily | | | | | | | |
| PWRIx | -0.07 (0.08) | -0.09 ($3.2\times10^{-3}$) | -0.09 ($3.4\times10^{-3}$) | -0.09 ($6.8\times10^{-3}$) | -0.16 (0.05) | **-0.09 ($1.2\times10^{-11}$)** | -0.08 (0.01) |
| BMI | -0.05 (0.31) | **-0.26 ($5.4\times10^{-17}$)** | **-0.27 (0)** | **-0.29 ($5.6\times10^{-45}$)** | -0.48 (0.60) | **-0.27 ($3.3\times10^{-21}$)** | **-0.29 ($7.6\times10^{-39}$)** |
| TC | -0.03 (0.32) | -0.07 (0.01) | -0.07 ($3.8\times10^{-3}$) | -0.07 (0.08) | -0.05 (0.93) | **-0.07 ($3.8\times10^{-12}$)** | -0.07 ($1.4\times10^{-3}$) |
| HDL-C | -0.07 ($3\times10^{-3}$) | **-0.08 ($2.8\times10^{-5}$)** | **-0.08 ($2.7\times10^{-5}$)** | -0.08 ($5.6\times10^{-3}$) | -0.12 (0.09) | **-0.08 ($6.8\times10^{-18}$)** | **-0.08 ($7.4\times10^{-5}$)** |
| LDL-C | -0.02 (0.46) | -0.06 (0.03) | -0.06 (0.01) | -0.06 (0.11) | -0.06 (0.32) | **-0.06 ($6.9\times10^{-7}$)** | -0.06 ($4.4\times10^{-3}$) |
| Age stopped smoking | | | | | | | |
| BMI | -0.16 ($8.1\times10^{-3}$) | **-0.41 ($3.4\times10^{-56}$)** | **-0.42 (0)** | **-0.34 ($4.7\times10^{-33}$)** | -1.08 (0.01) | **-0.42 ($1.1\times10^{-94}$)** | **-0.43 ($3.8\times10^{-82}$)** |
| SBP | 0.03 (0.55) | 0.04 (0.01) | 0.04 (0.01) | 0.05 (0.01) | 0.06 (0.14) | **0.05 ($9.5\times10^{-9}$)** | 0.05 (0.01) |
| HDL-C | **-0.10 ($2.3\times10^{-5}$)** | -0.09 ($9.2\times10^{-4}$) | -0.09 ($6.9\times10^{-4}$) | 0.07 (0.06) | -0.10 (0.61) | **-0.09 ($4.0\times10^{-8}$)** | **-0.11 ($7.0\times10^{-8}$)** |
| TG | 0.09 ($6.7\times10^{-4}$) | 0.08 ($6.1\times10^{-4}$) | **0.09 ($6.4\times10^{-7}$)** | **0.09 ($3.1\times10^{-6}$)** | 0.20 (0.12) | **0.09 ($1.1\times10^{-96}$)** | **0.09 ($8.7\times10^{-7}$)** |
| Age started smoking in current smokers | | | | | | | |
| SBP | 0.23 (0.14) | -0.26 (0.18) | -0.27 (0.18) | 0.24 (0.07) | -0.23 (0.45) | -0.26 (0.15) | **-0.49 ($7.9\times10^{-5}$)** |
| Number of cigarettes currently smoked daily (current cigarette smokers) | | | | | | | |
| PWRIx | 0.07 (0.36) | 0.10 (0.05) | 0.00 (0.06) | 0.11 (0.07) | 0.11 (0.14) | **0.13 ($4.1\times10^{-9}$)** | 0.13 (0.06) |
| BMI | -0.06 (0.38) | **-0.57 ($2.5\times10^{-9}$)** | **-0.58 ($1.6\times10^{-9}$)** | **-0.70 ($6.2\times10^{-50}$)** | -0.58 (0.39) | **-0.57 ($5.8\times10^{-7}$)** | **-0.69 ($6.4\times10^{-41}$)** |
| TC | -0.08 (0.05) | -0.13 ($1.9\times10^{-3}$) | -0.13 ($3.2\times10^{-3}$) | **-0.28 ($8.0\times10^{-10}$)** | -0.18 (0.08) | -0.13 ($2.0\times10^{-3}$) | -0.10 (0.04) |
| HDL-C | -0.13 ($1.5\times10^{-3}$) | -0.11 ($1.1\times10^{-3}$) | -0.11 ($1.5\times10^{-3}$) | -0.11 ($1.4\times10^{-3}$) | -0.02 (0.98) | **-0.11 ($6.0\times10^{-6}$)** | -0.12 ($3.9\times10^{-3}$) |
| Average weekly red wine intake | | | | | | | |
| TG | **-0.50 ($6.5\times10^{-6}$)** | **-0.52 ($4.0\times10^{-6}$)** | **-0.51 ($2.8\times10^{-6}$)** | **-0.33 ($3.2\times10^{-5}$)** | -0.73 (0.02) | -0.50 ($4.2\times10^{-3}$) | -0.40 (0.06) |
| Average monthly intake of other alcoholic drinks | | | | | | | |
| TG | 0.15 (0.58) | 0.18 (0.25) | 0.23 (0.09) | 0.37 ($4.4\times10^{-3}$) | 0.12 (0.47) | **0.34 ($2.7\times10^{-16}$)** | 0.36 (0.01) |
| Duration of walks | | | | | | | |
| PWAIx | -0.23 (0.35) | -0.29 (0.17) | -0.29 (0.21) | -0.29 (0.16) | **-0.05 ($2.1\times10^{-5}$)** | -0.29 ($4.4\times10^{-3}$) | -0.30 (0.18) |
| Duration of moderate activity | | | | | | | |
| PWP2PT | -0.21 (0.37) | -0.23 (0.21) | -0.24 (0.23) | -0.23 (0.23) | -0.10 (0.46) | **-0.41 (0)** | -0.41 (0.18) |
| Number of days/week of vigorous physical activity 10+ minutes | | | | | | | |
| PWP2PT | 0.40 (0.03) | 0.42 (0.01) | 0.43 (0.01) | **0.71 ($2.9\times10^{-5}$)** | 0.41 (0.16) | 0.43 (0.01) | 0.65 (0.01) |
| SBP | -0.33 (0.03) | -0.31 (0.35) | -0.36 ($5.6\times10^{-3}$) | **-0.46 ($4.8\times10^{-7}$)** | -0.35 (0.21) | -0.35 (0.01) | -0.35 ($6.6\times10^{-3}$) |
| Time spent watching television (TV) | | | | | | | |
| HDL-C | -0.06 (0.16) | **-0.13 ($3.4\times10^{-5}$)** | -0.12 ($1.0\times10^{-4}$) | -0.07 (0.67) | -0.15 (0.24) | -0.09 (0.04) | -0.08 (0.53) |
| BMI | | | | | | | |
| TSDriv | 0.65 (0.01) | **0.82 ($2.5\times10^{-6}$)** | **0.71 ($3.1\times10^{-5}$)** | 0.77 (0.12) | 0.71 (0.02) | 0.67 ($1.5\times10^{-4}$) | 0.52 (0.04) |
| AveWRWI | 0.01 (0.99) | -0.10 (0.63) | -0.06 (0.78) | **-0.96 ($4.9\times10^{-5}$)** | -0.08 (0.88) | -0.08 (0.75) | 0.37 (0.21) |
| TC | | | | | | | |
| AveWFWI | -1.08 (0.32) | **-1.47 ($2.2\times10^{-5}$)** | **-1.43 ($6.1\times10^{-5}$)** | -0.56 (0.42) | -1.46 (0.01) | **-1.42 ($2.9\times10^{-7}$)** | -1.35 (0.03) |

Values are bold if $p< 8\times10^{-5}$. PR, pulse rate; PWRIx, pulse wave reflection index; PWP2PT, pulse wave peak to peak time; PWAIx, pulse wave Arterial Stiffness index; BMI, body mass index; SBP, systolic blood pressure, automated reading; DBP, diastolic blood pressure, automated reading; TC, cholesterol; HDL-C, high density lipoprotein cholesterol; LDL-C, low density lipoprotein cholesterol; TG, triglycerides; TSDriv, time spent driving; AveWRWI, Average weekly red wine intake; AveWFWI, Average weekly fortified wine intake.

**Table S2. The UK Biobank traits analyzed in this study.**

| | Category | Trait | Field ID |
|---|---|---|---|
| lifestyle risk factors | Physical activity | Number of days/week walked 10+ minutes | 864 |
| | | Duration of walks | 874 |
| | | Number of days/week of moderate physical activity 10+ minutes | 884 |
| | | Duration of moderate activity | 894 |
| | | Number of days/week of vigorous physical activity 10+ minutes | 904 |
| | | Time spent watching television (TV) | 1070 |
| | | Time spent using computer | 1080 |
| | | Time spent driving | 1090 |
| | Alcohol | Average weekly red wine intake | 1568 |
| | | Average weekly champagne plus white wine intake | 1578 |
| | | Average weekly beer plus cider intake | 1588 |
| | | Average weekly spirits intake | 1598 |
| | | Average weekly fortified wine intake | 1608 |
| | | Average monthly red wine intake | 4407 |
| | | Average monthly champagne plus white wine intake | 4418 |
| | | Average monthly beer plus cider intake | 4429 |
| | | Average monthly spirits intake | 4440 |
| | | Average monthly fortified wine intake | 4451 |
| | | Average monthly intake of other alcoholic drinks | 4462 |
| | | Average weekly intake of other alcoholic drinks | 5364 |
| | Diet | Cooked vegetable intake | 1289 |
| | | Salad / raw vegetable intake | 1299 |
| | | Fresh fruit intake | 1309 |
| | | Dried fruit intake | 1319 |
| | | Bread intake | 1438 |
| | | Cereal intake | 1458 |
| | | Tea intake | 1488 |
| | | Coffee intake | 1498 |
| | | Water intake | 1528 |
| | | Age when last ate meat | 3680 |
| | Smoking | Exposure to tobacco smoke at home | 1269 |
| | | Exposure to tobacco smoke outside home | 1279 |
| | | Age started smoking in former smokers | 2867 |
| | | Number of cigarettes previously smoked daily | 2887 |
| | | Age stopped smoking | 2897 |
| | | Number of unsuccessful stop-smoking attempts | 2926 |
| | | Age started smoking in current smokers | 3436 |
| | | Number of cigarettes currently smoked daily (current cigarette smokers) | 3456 |
| CVD-related traits | Arterial stiffness | Pulse rate | 4194 |
| | | Pulse wave reflection index | 4195 |
| | | Pulse wave peak to peak time | 4196 |
| | | Pulse wave Arterial Stiffness index | 21021 |
| | Body size measures | Body mass index (BMI) | 21001 |
| | Blood pressure | Diastolic blood pressure, automated reading | 4079 |
| | | Systolic blood pressure, automated reading | 4080 |
| | Blood biochemistry | Cholesterol | 30690 |
| | | HDL cholesterol | 30760 |
| | | LDL direct | 30780 |
| | | Triglycerides | 30870 |

**Table S3. Seven MR methods in this study and the corresponding software.**

| Method | R package | Version | Function | Tuning parameters |
|---|---|---|---|---|
| IVW-R[54] | MendelianRandomization | 0.4.0 | mr_ivw | default settings |
| Weighted mode[55] | MendelianRandomization | 0.4.0 | mr_mbe | default settings |
| Robust[57] | MendelianRandomization | 0.4.0 | mr_ivw | (model="random", robust=TRUE), other settings are default |
| RAPS[6] | mr.raps | NA | mr.raps.overdispersed.robust | (b_exp,b_out,se_exp,se_out,loss.function="huber",k=1.345,initialization=c("l2"),suppress.warning=FALSE,diagnostics=FALSE,niter=20,tol=.Machine$double.eps^0.5) |
| MRMix[7] | MRMix | NA | MRMix | default settings |
| CAUSE[8] | cause | NA | cause | default settings |
| **MRAID** | **MRAID** | **NA** | **MRAID** | **default settings** |
